# Brain microstructural changes and fatigue after COVID-19

**DOI:** 10.1101/2022.08.20.22279023

**Authors:** Diógenes Diego de Carvalho Bispo, Pedro Renato de Paula Brandão, Danilo Assis Pereira, Fernando Bisinoto Maluf, Bruna Arrais Dias, Hugo Rafael Paranhos, Felipe von Glehn, Augusto César Penalva de Oliveira, Neysa Aparecida Tinoco Regattieri, Lucas Scardua Silva, Clarissa Lin Yasuda, Alexandre Anderson de Sousa Munhoz Soares, Maxime Descoteaux

## Abstract

**Background:** Fatigue and cognitive complaints are the most frequent persistent symptoms in patients after severe acute respiratory syndrome coronavirus 2 (SARS-CoV-2) infection. This study aimed to assess fatigue and neuropsychological performance and investigate changes in the thickness and volume of gray matter (GM) and microstructural abnormalities in the white matter (WM) in a group of patients with mild-to-moderate coronavirus disease 2019 (COVID-19).

**Methods:** We studied 56 COVID-19 patients and 37 matched controls using magnetic resonance imaging (MRI). Cognition was assessed using Montreal Cognitive Assessment and Cambridge Neuropsychological Test Automated Battery, and fatigue was assessed using Chalder Fatigue Scale (CFQ-11). T1-weighted MRI was used to assess GM thickness and volume. Fiber-specific apparent fiber density (FD), free water index, and diffusion tensor imaging data were extracted using diffusion-weighted MRI (d-MRI). d-MRI data were correlated with clinical and cognitive measures using partial correlations and general linear modeling.

**Results:** COVID-19 patients had mild-to-moderate acute illness (95% non-hospitalized). The average period between real-time quantitative reverse transcription polymerase chain reaction-based diagnosis and clinical/MRI assessments was 93.3 (±26.4) days. The COVID-19 group had higher CFQ-11 scores than the control group (*p* < 0.001). There were no differences in neuropsychological performance between groups. The COVID-19 group had lower FD in the association, projection, and commissural tracts, but no change in GM. The corona radiata, corticospinal tract, corpus callosum, arcuate fasciculus, cingulate, fornix, inferior fronto-occipital fasciculus, inferior longitudinal fasciculus, superior longitudinal fasciculus, and uncinate fasciculus were involved. CFQ-11 scores correlated with microstructural changes in patients with COVID-19.

**Conclusions:** Quantitative d-MRI detected changes in the WM microstructure of patients recovering from COVID-19. This study suggests a possible brain substrate underlying the symptoms caused by SARS-CoV-2 during medium- to long-term recovery.

**Key points:**

- Patients with COVID-19 had microstructural changes in the WM at a mean follow-up of 3 months.
- There is a possible brain substrate underlying the symptoms caused by SARS-CoV-2 during medium- to long-term recovery.
- A serial d-MRI study following up on a non-hospitalized sample of patients with milder COVID-19 forms is warranted.

## 1 INTRODUCTION

The sequelae of coronavirus disease 2019 (COVID-19) beyond the acute phase of infection are being increasingly understood as scientific research and clinical experience accumulate, and, in this sense, studies that include the identification and characterization of clinical, serological, and imaging of COVID-19 in the acute, subacute, and chronic phases of the disease are needed.^1^ People with post-COVID conditions can have a wide range of symptoms, lasting for more than 4 weeks, but commonly for months after infection. These symptoms must not be explained by an alternative diagnosis.^2^ Several symptoms, such as fatigue, myalgia, anosmia, dysgeusia, and cognitive impairment (difficulty concentrating and memory complaints) have been reported in post-COVID.^3^ Symptoms may appear following recovery from acute COVID-19, persist for an extended period, fluctuate, or relapse over time.^1,4^

Perceived fatigue following severe acute respiratory syndrome coronavirus 2 (SARS-CoV-2) infection is more pronounced than in the general population and does not depend on initial COVID-19 severity.^5^ Cognitive deficits seem to occur even in non-hospitalized individuals with mild acute symptoms.^6^ Decreased performance in attention and working memory has been reported,^7^ as well as in reasoning, problem-solving, spatial planning, processing speed,^8^ verbal fluency, and visuospatial construction.^9^ The nature and causes of fatigue and cognitive dysfunction across the COVID-19 severity spectrum remain, however, disputed.

Numerous hypotheses have been proposed to explain the mechanisms underlying post-COVID symptoms. Direct viral infection effects, systemic inflammation, neuroinflammation (due to cytokine-induced microglial activation), microvascular thrombosis, blood-brain barrier disruption, and even viral-induced neurodegeneration may play a role.^10^ In critical cases, hypoxic-ischemic changes are associated with infarcts, microhemorrhage, microglial activation, microglial nodules, and neuronophagia.^11^ However, hypoxic-ischemic changes and microglial-induced damage may not occur in mild-to-moderately symptomatic patients with no hypoxia, a fact that encourages alternative biological explanations.

Post-COVID brain imaging characteristics were also examined. Tractometry and volume-based magnetic resonance imaging (MRI) measurements in patients 3 months after COVID-19 have shown changes in white matter (WM) microstructure metrics, especially in the frontal and limbic systems, in both mild and severe cases.^12^ In a large sample derived from the UK Biobank study, SARS-CoV-2 infection was associated with changes in brain structure.^13^ Significant longitudinal effects were identified: a more substantial reduction in the cortical thickness of the orbitofrontal and parahippocampal gyrus, as well as prominent changes in tissue damage markers in brain regions functionally linked to the primary olfactory cortex. Furthermore, stronger overall brain atrophy was observed in those infected with SARS-CoV-2 than in the control cohort examined at similar time intervals.^13^ With regard to nuclear medicine techniques, frontoparietal hypometabolism was identified in fluorodeoxyglucose-positron emission tomography examinations studying post-COVID, correlating with the Montreal Cognitive Assessment (MoCA) performance.^14^ Neuroimaging techniques, thus, seem to serve as surrogate biomarkers of post-COVID neurological abnormalities.

Diffusion-weighted MRI (d-MRI) generates three families of potentially useful metrics to investigate post-COVID structural brain damage. The first, voxel-wise diffusion tensor imaging (DTI) measures, relate to the main eigenvector and eigenvalue of the elliptical unidirectional tensor.^15,16^ The second, free water (FW) imaging, investigates tissue changes by separating the contribution of freely diffusing extracellular water from the tissue component.^17^ In this two-compartment model, extracellular FW represents changes caused by neuroinflammation, atrophy, or edema. The third, apparent fiber density (AFD), derived from constrained spherical deconvolution,^18^ represents an indirect measure of axon degeneration, reflecting an apparent number of axons.^19^ AFD is computed in two distinct ways. AFDtotal represents the total number of axons in a voxel, integrating all the diffusion orientations. On the other hand, FD stands in for a fiber population within a single voxel, overcoming the “crossing-fibers” interpretation issue.^20^

The current study assessed fatigue and general cognitive performance, examined changes in GM thickness and volume, and investigated WM microstructural abnormalities after COVID-19 compared to a control group using FW imaging, voxel-based analysis, and fixel-based analysis. Our secondary objective was to determine whether microstructural changes were associated with clinical and cognitive data.

## 2 METHODS

### 2.1 Participants

This cross-sectional prospective analytical study was conducted as part of the NeuroCOVID-19 Brazilian Registry.^21^ Participants were recruited between October 2020 and May 2021 in Brasilia, Brazil, from a population of health professionals and patients assisted at the Brasilia University Hospital, before the implementation of mass vaccination campaigns, with a non-probabilistic sampling strategy. During the recruitment period, a timeframe that corresponded approximately to alpha and gamma (P1) variants predominance in Brazil, we consecutively reached out by phone to a list of 364 patients who were diagnosed with COVID-19 by real-time quantitative reverse transcription polymerase chain reaction (qRT-PCR) to invite them to participate in the study.

The inclusion criteria for the COVID-19 group (COV+) were (a) diagnosis of SARS-CoV-2 infection confirmed by detection of viral RNA by qRT-PCR testing of a nasopharyngeal swab, (b) at least one COVID-19-related symptom during the acute phase of infection, and (c) 18–60 years of age. Patients were evaluated at least 4 weeks after diagnosis of COVID-19. The control group (COV-) was recruited from the same population (patients or health professionals from Brasilia University Hospital) through convenience sampling, matching for age, sex, and education level. Subjects in the COV- group were not previously infected with SARS-CoV-2 and had a negative SARS-CoV IgG/IgM test.

The exclusion criteria for both groups were (a) pre-existing brain structural disorders (stroke, epilepsy, multiple sclerosis, neoplasia, hydrocephalus, traumatic brain injury, Parkinson’s disease, and dementia), (b) severe psychiatric diseases, (c) previous hospital admission with treatment in an intensive care unit who required mechanical ventilation, and (c) illiteracy.

Each participant signed a consent form and underwent clinical, cognitive, and MRI examinations. All the procedures were performed on the same day. This study was approved by the local ethics committee of the University of Brasilia. All procedures adhered to current regulations, such as the Helsinki Declaration.

### 2.2 Clinical assessment

Demographic and clinical data were collected using an electronic form. Age, education, sex, and a comorbidity checklist were collected during anamnesis with the aim of identifying potential confusion variables. Current neurological, chemosensory, respiratory, and constitutional symptoms were evaluated. The participants reported symptoms that occurred during the acute and post-acute phases of COVID-19.

The Chalder Fatigue Scale (CFQ-11) was used to evaluate fatigue severity and extent.^22,23^ Using a prespecified CFQ-11 cut-off greater than or equal to 16, we dichotomized participants into no fatigue vs. increased fatigue.^24,25^

### 2.3 Cognitive assessment

All participants underwent a cognitive screening examination, MoCA,^26^ followed by a comprehensive cognitive assessment using the Cambridge Neuropsychological Test Automated Battery (CANTAB).^27,28^ This battery assesses executive functions (One Touch Stockings of Cambridge), verbal memory (Verbal Recognition Memory), visual memory (Paired Associates Learning, Pattern Recognition Memory), working memory (Spatial Working Memory), and reaction time (simple and five-choice Reaction Time). **Supplementary Table 1** summarizes the key cognitive variables.

### 2.4 MRI data acquisition

MRI was performed using a Philips Achieva 3T scanner (Best, Netherlands) equipped with an 8-channel SENSE coil. The following sequences were obtained: (a) Three dimensional (3D) T1-weighted sequence, turbo field echo, sagittal, with field of view (FOV) = 208 × 240 × 256 mm, reconstructed resolution of 1 × 1 × 1 mm, echo time (TE) = min full echo, repetition time (TR) = 2300 ms, TI = 900 ms, two times accelerated acquisition; (b) Diffusion-weighted sequence, axial, with FOV 232 × 232 × 160 mm, reconstructed resolution of 2 × 2 × 2 mm, TE = 71 ms; TR = 3300 ms, 32 directions (b = 800 s/mm2); (c) Diffusion-weighted sequence, axial, with FOV 232 × 232 × 160 mm, reconstructed resolution of 2 × 2 × 2 mm, TE = 71 ms; TR = 3300 ms (reversed phase encoded b0); (d) 3D-fluid attenuated inversion recovery (FLAIR) sequence, sagittal, with FOV 256 × 256 × 160 mm, reconstructed resolution of 1.2 × 1 × 1 mm, TE = 119 ms, TR = 4800 ms, TI = 1650 ms.

### 2.5 Automated cortical and subcortical segmentation

MRI data were processed using the FreeSurfer suite (version 7.1)^29^ to estimate cortical thickness and deep GM nuclei volume. Cortical thickness was extracted by measuring the distance between the WM and GM boundary and the pial surface. Cortical parcellation maps capable of detecting submillimeter differences between the groups were created using spatial intensity gradients. To smoothen the cortical maps, a circularly symmetric Gaussian kernel with a full width at half maximum of 10 mm was applied.

The volumes of subcortical and limbic structures were measured using automated procedures that assigned a neuroanatomical label to each voxel in the MRI volume. This procedure is based on probabilistic information estimated from a manually labeled training set. The caudate, putamen, globus pallidum, hippocampus, nucleus accumbens, and amygdala were bilaterally segmented. To avoid biases related to unequal head size, the volumes were normalized to the intracranial volume.

### 2.6 Diffusion-weighted MRI processing

TractoFlow^30^ was used to analyze d-MRI and T1-weighted images (**Figure 1**). As an automated tool for processing d-MRI, it extracts DTI and high angular resolution diffusion-weighted imaging (HARDI) measures. The fractional anisotropy (FA), mean diffusivity (MD), radial diffusivity (RD), and axial diffusivity (AD) were calculated. In addition, voxel-wise AFD (AFDtotal) values were extracted from the fiber orientation distribution function. Probabilistic whole-brain tractography was used. The standardized processing steps have been detailed elsewhere.^30^

**Figure 1.**
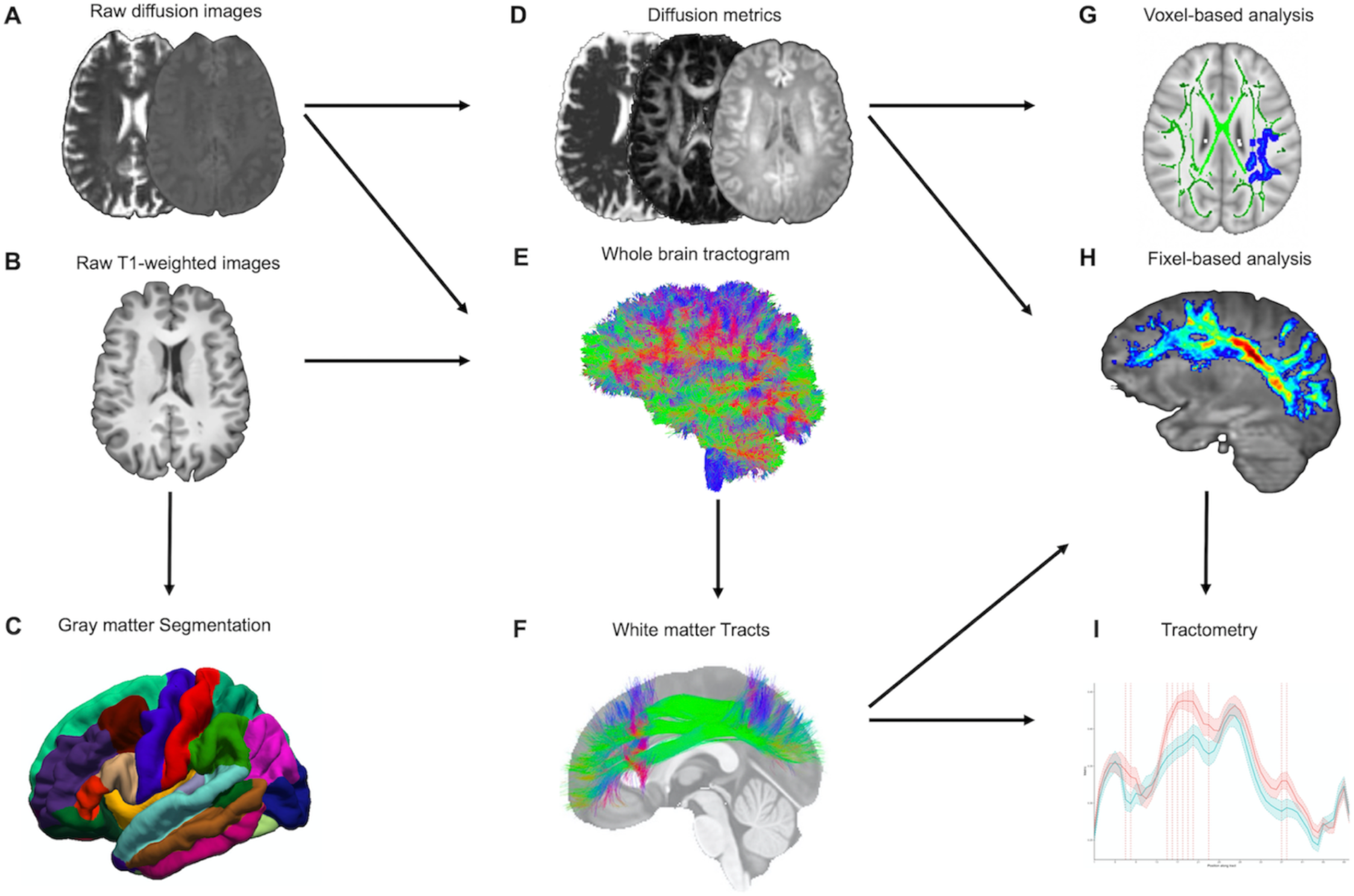
MRI processing pipeline. (A and B) Raw diffusion-weighted and T1-weighted images are processed by the TractoFlow pipeline. (C) Raw T1-weighted images also are processed by the FreeSurfer suite for gray matter segmentation. (D) Diffusion MRI-derived measures and free-water fraction are computed. (E) Whole brain probabilistic tractography is performed using an anatomically constrained particle filter algorithm. (F) Extraction of the white matter tracts by RecoBundlesX (e.g., SLF). (G) Voxel-based analysis was used to investigate the metrics FW, FAt, MDt, RDt, ADt, and AFDtotal. (H) Fiber-specific apparent fiber density (FD) is extracted at each fixel (e.g., SLF). (I) Tractometry of each bundle using the FD. Abbreviations: FW, free water; FAt, tissue fractional anisotropy; MDt, tissue mean diffusivity; RDt, tissue radial diffusivity; ADt, tissue axial diffusivity; AFDtotal, voxel-wise apparent fiber density; FD, fiber-specific apparent fiber density; SLF, superior longitudinal fasciculus.

The FW imaging analysis followed the methods described elsewhere.^17^ The FW maps at each voxel were reconstructed using a two-tensor model, with values ranging from 0 to 1. Values close to 0 indicate negligible FW diffusion in the extracellular space, whereas 1 indicates unrestricted FW diffusion (i.e., water in a voxel diffuses completely freely). While the FW parameter quantifies the fractional volume of free water found in the extracellular space, the tissue compartment is fitted to a diffusion tensor that accounts for the remaining signal after the removal of free water. As a result, it generates FW-corrected measures, which are expected to be more sensitive and specific to tissue changes than single tensor model-derived measures. Tissue fractional anisotropy (FAt), mean diffusivity (MDt), radial diffusivity (RDt), and axial diffusivity (ADt) denote the FW-corrected DTI maps.^17^

### 2.7 Voxel-based diffusion imaging analysis (VBA)

The tract-based spatial statistics (TBSS) pipeline in FSL (version 6.0)^31^ permitted the investigation of d-MRI metric contrasts between the COV+ and COV- groups. The FA maps were nonlinearly aligned to the FMRIB-58 map from the Montreal Neuroimaging Institute template space. The mean FA skeleton was computed following the deformable registration. The deformation fields from the FA maps were used for FAt, MDt, RDt, ADt, FW, and AFDtotal. The registered maps were projected onto the FA skeleton.

### 2.8 Segmentation of WM tracts

A multi-atlas and multi-parameter version of RecoBundles extracted preselected WM bundles from whole-brain tractography.^32,33^ RecoBundles recognizes bundles based on the similarities between a subject’s streamline and a template or atlas. In RecobundlesX, the algorithm was repeated with different parameters, followed by label fusion. This tool is based on shape similarity to a template constructed from anatomical prior-inspired delineation rules. For both groups, a bundle-specific tractography approach was used to reconstruct the “hard-to-track” fornix pathway.^34^ The overall approach, entirely performed in native space, has the advantage of generating unique bundles for each individual (**Figure 1**).

### 2.9 Tract-wise analysis

The fiber-specific AFD (FD) values were computed for each fixel, representing a particular fiber orientation. The AFD signal in a fixel is proportional to the volume of axons aligned in that direction. The mean FAt, MDt, RDt, ADt, FW, and FD values were calculated by averaging the template-normalized metric image values across voxels/fixels contained within the mask.

Subsequently, each bundle was subjected to tractometry^35^ using previously computed diffusion measures and FW index. This method was chosen because, depending on the underlying WM fiber organization, d-MRI measures may vary throughout the studied bundles.^36^ Tractometry provides higher sensitivity to the pathway’s microstructure by mapping a set of measures over the WM bundles. Finally, each bundle was divided into 50 segments along its length to provide additional topological insight.

### 2.10 MRI Quality control

Every raw and processed MRI dataset was inspected for gross geometric distortion, bulk motion, or signal dropout artifacts. T1-weighted and d-MRI datasets were examined using Dmriqc-flow^37^ for d-MRI quality control. The cortical and subcortical segmentations and WM tracts were visually reviewed by a board-certified neuroradiologist to ensure accuracy.

### 2.11 Statistical analysis

#### 2.11.1 Demographic, clinical, and cognitive assessments

The clinical characteristics were compared between the groups using independent-sample t-tests for normally distributed continuous variables, the Mann-Whitney test for non-normally distributed data, and χ^2^ for categorical inputs. Normality was assessed by visual inspection of histograms and the Shapiro-Wilk test. Statistical significance was set at *p* < 0.05. Statistical analyses were performed using R, v4.1.0 (*R Foundation for Statistical Computing*, Vienna, Austria).

#### 2.11.2 FreeSurfer

Each hemisphere’s vertex-wise cortical thickness was computed using generalized linear models (GLM). Patients were compared to controls employing FreeSurfer’s “*mri glmfit*”.^29^ Monte Carlo simulations with a p-value set to 0.001 corrected for multiple comparisons. Age and sex were used as nuisance covariates. A GLM was used to analyze differences in the volume of GM subcortical nuclei between the two groups, using age, sex, and intracranial volume as covariates. All results were corrected using the *false discovery rate* (FDR) method.

#### 2.11.3 VBA

For VBA, GLM with contrast was performed to test for group differences and correlations. The TBSS framework^31^ includes nonparametric permutation testing (5,000 permutations) to correct for multiple comparisons and *threshold-free cluster enhancement* (TFCE). Age and sex were used as nuisance covariates. Results were considered significant at *p* < 0.05, TFCE corrected for multiple comparisons. WM regions were named according to the Johns Hopkins University white-matter tractography atlas.^38^

#### 2.11.4 Tract-wise analysis

Comparisons of tract-average FW, FAt, MDt, RDt, ADt, and FD values between the groups were performed using GLM, adjusting for age and sex. FDR correction was performed for the 35 tracts tested using the Benjamini-Hochberg procedure.

Each tract was divided into 50 sections for further examination. Contrasts between groups were calculated with t-tests for each bundle subsection.^35,36^ The procedure aimed to explore bundle segments that were contrasted between the COV+ and COV- groups. To increase the statistical robustness and account for multiple comparisons, each t-test was repeated with 10,000 permutations to generate a corrected significance threshold.^39^ A t-test was considered statistically significant if the p-value was <0.05, and its t-absolute values exceeded the computed threshold. The purpose of this analysis was to ensure that the observed changes were distributed uniformly along the bundle, as fanning of the fibers at the extremities of a bundle could bias the diffusion measurements.

In the COV+ group, we performed a partial correlation analysis between tract-average measures and CFQ-11 scores, adjusting for age, sex, and education. Data underwent a non-paranormal transformation and were analyzed using Pearson’s coefficient. Statistical significance was defined as a two-tailed p-value <0.05, with FDR correction for multiple comparisons.

## 3 RESULTS

### 3.1 Demographic and clinical characteristics

Initially, we recruited 97 participants (**Figure 2**). In the COV+ group, two participants were excluded because of MRI contraindications. Two participants from the COV- group were excluded: one due to a positive SARS-CoV-2 IgG test result and another because of a brain structural change on MRI.

**Figure 2.**
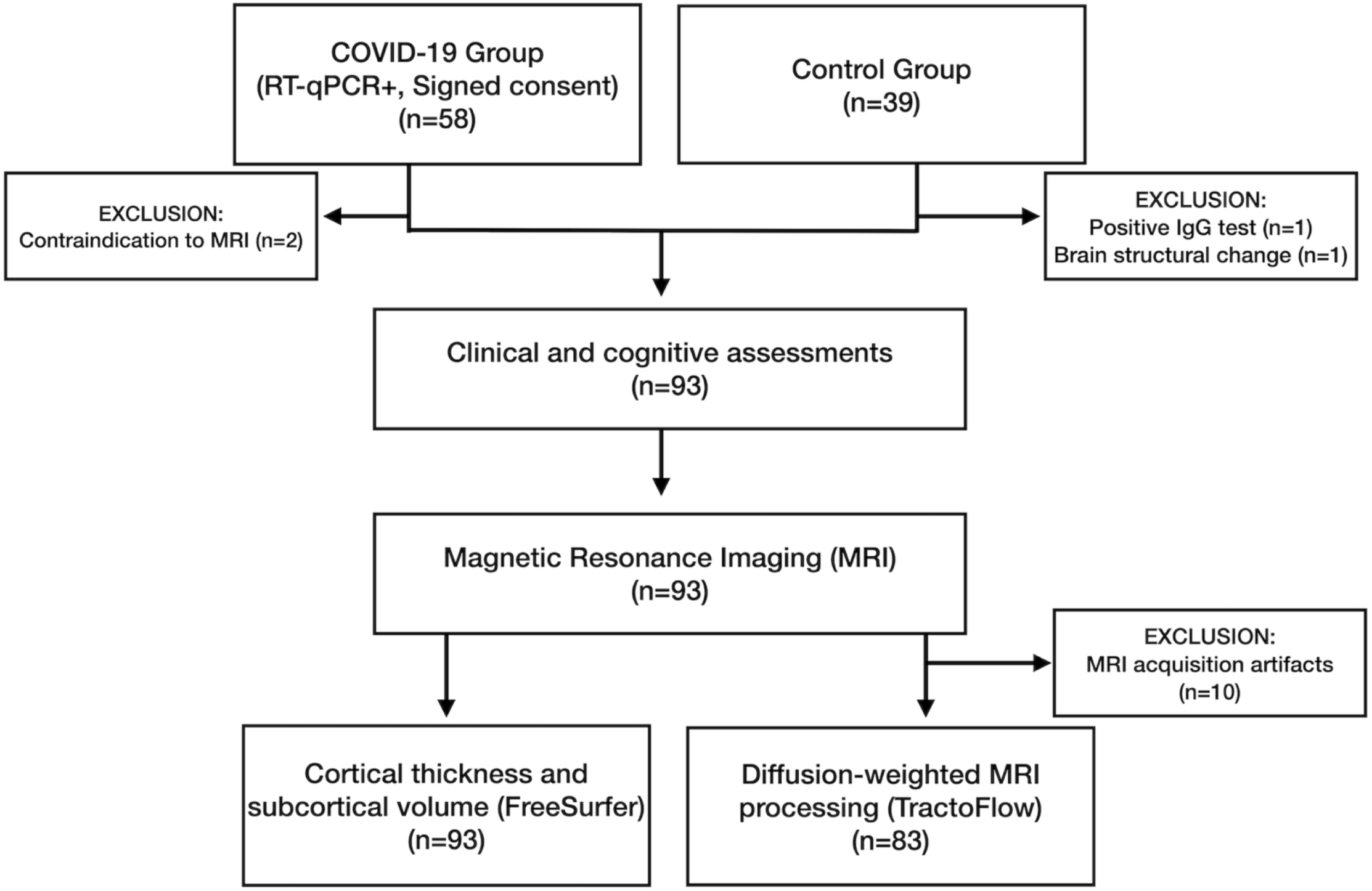
Flowchart with the enrollment of participants in the COVID-19 (COV+) and control (COV-) groups and the investigations that were carried out.

Ninety-three participants underwent clinical examinations, cognitive tests, and MRI: 56 in the COV+ group and 37 in the COV- group. The groups did not differ in age (*p* = 0.237), sex (*p* = 0.638), education (*p* = 0.840), or comorbidity profiles (**Table 1**). The average time between COVID-19 diagnosis and study clinical/imaging procedures was 93.3 (±26.4) days, ranging from 31 to 167 days. Most patients (95%) did not require hospitalization. None of the patients required mechanical ventilation.

**Table 1.** Demographic and clinical features (COV+ and COV- groups).

| **Demographic and Clinical characteristics** | **COVID-19 (COV+)**<br>**(n=56, 60%)** | **Control (COV-)**<br>**(n= 37, 40%)** | **Statistic** |
| --- | --- | --- | --- |
| **Age** | 37.2 ± 9.4 (20, 57) | 40.2 ± 11.8 (22, 60) | U = 885; p = 0.237<sup>1</sup> |
| **Gender** | | | $\chi^2 = 0.22$; p = 0.638<sup>2</sup> |
| Male, n (%) | 20 (36.0%) | 15 (41.0%) |  |
| Female, n (%) | 36 (64.0%) | 22 (59.0%) |  |
| **Years of formal education** | 15.3 ± 3.3 (11, 24) | 15.0 ± 3.3 (8, 21) | U = 1010; p = 0.840<sup>1</sup> |
| **Self-reported comorbidities n (%)** |  |  |  |
| Hypertension | 5 (8.9%) | 3 (8.1%) | $\chi^2 = 0.02$; p = 0.890<sup>2</sup> |
| Diabetes mellitus | 5 (8.9%) | 3 (8.1%) | $\chi^2 = 0.02$; p = 0.890<sup>2</sup> |
| Obesity | 1 (1.8%) | 3 (8.1%) | $\chi^2 = 2.16$; p = 0.141<sup>2</sup> |
| Asthma/COPD | 2 (3.6%) | 2 (5.4%) | $\chi^2 = 0.18$; p = 0.670<sup>2</sup> |
| Allergic rhinosinusitis | 15 (27.0%) | 10 (27.0%) | $\chi^2 = 0.00$; p = 0.980<sup>2</sup> |
| Thyroid disorder | 3 (5.4%) | 1 (2.7%) | $\chi^2 = 0.38$; p = 0.537<sup>2</sup> |
| Mood disorder | 4 (7.1%) | 2 (5.4%) | $\chi^2 = 0.11$; p = 0.739<sup>2</sup> |
| Migraine | 14 (25.0%) | 7 (19.0%) | $\chi^2 = 0.47$; p = 0.492<sup>2</sup> |
| **Chalder fatigue scale (CFQ-11)** |  |  |  |
| Total score CFQ-11 | 16.3 ± 7.5 (0, 29) | 9.2 ± 7.4 (0, 26) | t = 4.502; p = <0.001<sup>3</sup> |
| Cut-off ≥ 16 n (%) | 33 (58.9%) | 8 (21.6%) | $\chi^2 = 12.58$; p = <0.001<sup>2</sup> |
| **Average time between positive qRT-PCR and clinical** | 93.3 ± 26.4 (31, 167) | - | - |
| **Acute phase treatment scenario n (%)** |  |  |  |
| Inpatient | 3 (5.2%) |  |  |
| Outpatient | 53 (94.6%) |  |  |
| **Oxygen supplementation n (%)** |  |  |  |
| Non-invasive ventilation or high flow mask | 2 (3.6%) |  |  |
| Low flow nasal catheter | 3 (5.4%) |  |  |
| None | 51 (91.0%) |  |  |

Abbreviations: CFQ-11, Chalder Fatigue Scale; COPD, chronic obstructive pulmonary disease; PD-CRS, Parkinson's Disease-Cognitive Rating Scale; qRT-PCR, real-time quantitative reverse transcription polymerase chain reaction; MRI, magnetic resonance imaging.
Data are shown as mean ± standard deviation (minimum, maximum) or n (%).
Legend:1 Mann-Whitney U test, 2 Chi-square test, 3 independent-sample t-test

All COV+ patients had at least two COVID-19-related symptoms during the acute phase of infection. The main acute-phase symptoms were headache (80.4%), hyposmia (80.4%), myalgia (73.2%), dysgeusia (67.9%), fatigue (60.7%), hyporexia (53.6%), fever (50.0%), dry cough (46.4%), sore throat (44.6%), nasal discharge (44.6%), and dyspnea (39.3%).

The prevalence of post-acute COVID-19 symptoms was also estimated. Of 56 COVID-19 patients, 52 (92.8%) had at least one post-COVID symptom. Hyposmia occurred in 71.4%, fatigue in 51.8%, headache in 44.6%, sustained attention complaints in 39.3%, memory complaints in 37.6%, dysgeusia in 33.9%, daytime sleepiness in 28.6%, dyspnea in 17.9%, and difficulty in daily activities in 14.3%. The COV+ group scored higher on the CFQ-11 scale (*p* < 0.001) (**Table 1**).

All participants underwent cognitive assessments and MRI. Ten participants were excluded from the d-MRI analysis because of head motion artifacts (**Figure 2**).

### 3.2 Cognitive assessment

The COV+ and COV- groups did not differ with respect to the MoCA global score. There were no differences in CANTAB neurocognitive performance between the groups (**Table 2**).

**Table 2.** Cognitive function comparison between COV+ and COV- groups.

| Cognitive Measure | COVID-19 (COV+)<br>(n=56, 60%) | Control (COV-)<br>(n= 37, 40%) | Statistic |
| --- | --- | --- | --- |
| **Spatial Working Memory** |  |  |  |
| SWMBE (between-errors) | 15 (5, 22) | 12 (4, 17) | U = 909; p = 0.320<sup>1</sup> |
| SWMS (strategy use) | 8.5 (7, 10) | 8 (7, 9) | U = 984; p = 0.683<sup>1</sup> |
| **One Touch Stockings of Cambridge** |  |  |  |
| OTSPSFC (number of attempts) | 10 (8.8, 12) | 11 (9, 12) | U = 1035; p = 0.997<sup>1</sup> |
| OTSMDLFC (average latency, ms) | 12,770 (8,819, 15,913) | 12,247 (9,347, 15,530) | U = 1021; 0.909<sup>1</sup> |
| OTSMCC (average of choices) | 1.50 (1.20, 1.80) | 1.47 (1.27, 1.87) | U = 1019; 0.897<sup>1</sup> |
| OTSMCLC (average latency, ms) | 27,908 (18,372, 35,436) | 23,745 (20,138, 29,896) | U = 941; p = 0.458<sup>1</sup> |
| **Paired Associates Learning** |  |  |  |
| PALTEA (total error adjusted) | 8 (5, 17) | 12 (7, 21) | U = 892; 0.258<sup>1</sup> |
| PALFAMS (first attempt memory score) | 13.12 (4.23) | 12.27 (3.88) | t = 0,985; p = 0.327<sup>2</sup> |
| PALMETS (number of attempts) | 2.0 (0.4, 2.0) | 2.0 (1.0, 3.0) | U = 823; 0.085<sup>1</sup> |
| **Pattern Recognition Memory** |  |  |  |
| PRMPCI (% correct, immediate) | 95.8 (83.3, 100.0) | 100.0 (91.7, 100.0) | U = 946; p = 0.446<sup>1</sup> |
| PRMPCD (% correct, delayed) | 91.7 (75.0, 91.7) | 91.7 (83.3, 100.0) | U = 997; p = 0.756<sup>1</sup> |
| **Verbal Recognition Memory** |  |  |  |
| VRMIRTC (immediate, total correct) | 31 (28, 33) | 30 (28, 32) | U = 961; p = 0.557<sup>1</sup> |
| VRMDRTC (delayed, total correct) | 32 (30, 34) | 31 (30, 32) | U = 887; p = 0.241<sup>1</sup> |
| VRMFRDS (free recall distinct stimuli) | 6.5 (5.0, 8.0) | 6.0 (4.0, 8.0) | U = 900; p = 0.284<sup>1</sup> |
| **Reaction Time** |  |  |  |
| RTISMDRT (single choice reaction time, ms) | 328 (311, 360) | 328 (307, 344) | U = 914; p =0.338<sup>1</sup> |
| RTISMDMT (single choice mov. time, ms) | 219.73 (64.61) | 219.35 (55.28) | t = 0.029; p = 0.977<sup>2</sup> |
| RTIFMDRT (five choice reaction time, ms) | 383 (360, 429) | 380 (354, 418) | U = 924; 0.379<sup>1</sup> |
| RTIFMDMT (five choice mov. time, ms) | 261.80 (75.10) | 249.58 (29.29) | t = 0.833; p = 0.407<sup>2</sup> |
| **MoCA** |  |  |  |
| Global Score | 25 (22, 27) | 25 (22, 28) | U = 1031; p = 0.969<sup>1</sup> |

Abbreviations: MoCA, Montreal Cognitive Assessment; OTS, One Touch Stockings of Cambridge; PAL, Paired Associates Learning; PD-CR, Parkinson's Disease-Cognitive Rating Scale; PRM, Pattern Recognition Memory; RTI, Reaction time; SD, standard deviation; SWM, Spatial Working Memory; VRM, Verbal Recognition Memory; ms, milliseconds; mov., movement.
PALFAMS, RTISMDMT, and RTIFMDMT are shown as mean (standard deviation).
The other data are shown as median (interquartile range).
Legend: <sup>1</sup> Mann-Whitney U test, <sup>2</sup> independent-sample t-test

### 3.3 Cortical thickness and subcortical structures volume

The vertex-wise cortical thickness did not differ between the groups. The caudate, putamen, pallidum, thalamus, accumbens, hippocampus, and amygdala volumes did not differ (all *p* > 0.120).

### 3.4 VBA

#### 3.4.1 COV+ vs. COV- group comparison

To explore AFD total between-group contrasts, whole-brain TBSS analysis was employed, adjusting for age and sex effects. The COV+ group had lower AFDtotal values than the COV- group across 4,515 voxels (*p* < 0.05, TFCE-corrected; **Figure 3A**; **Supplementary Table 2**). The affected tracts included the left anterior thalamic radiation, corticospinal tract, cingulum (cingulate gyrus), inferior fronto-occipital fasciculus, inferior longitudinal fasciculus, superior longitudinal fasciculus, and superior longitudinal fasciculus (temporal part). No between-group differences were observed for FAt, MDt, RDt, ADt, and FW using TBSS.

**Figure 3.**
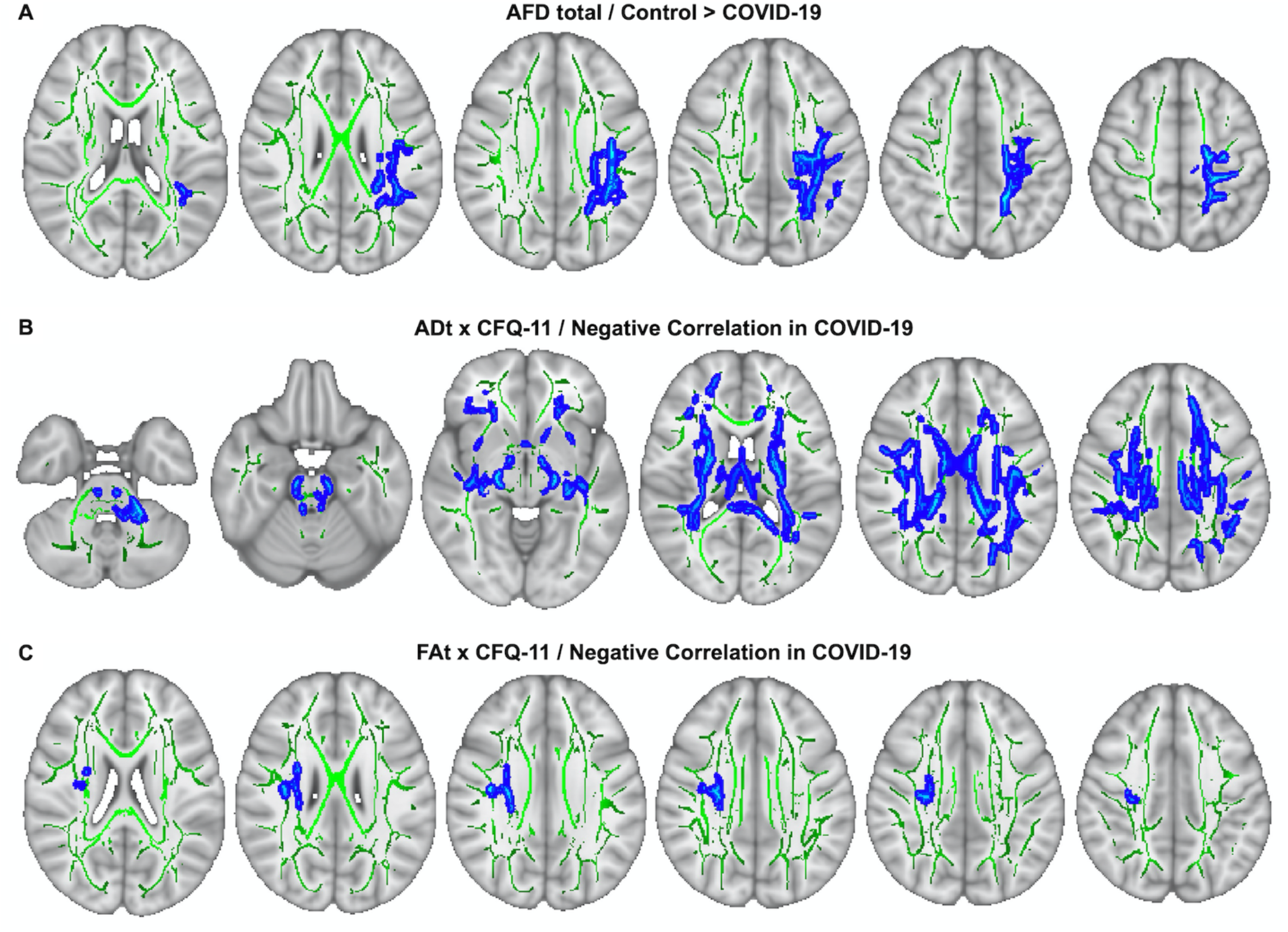
TBSS analysis. (A) AFDtotal voxel-wise analysis compares patients with COV+ and COV- groups. Areas where AFDtotal values in the COV+ group are significantly lower than in COV- within WM skeleton (green) are reported on a blue scale (p-values ranging from 0.05 to <0.01). (B) ADt voxel-wise analysis, correlating this DTI metric with CFQ-11 total score, in COV+ group. The blue scale depicts WM regions where negative correlations between ADt and CFQ-11 are found (p-values ranging from 0.05 to <0.01). (C) FAt voxel-wise analysis, correlating this DTI metric with CFQ-11 total score, in COV+ group. The blue scale depicts WM regions where negative correlations between FAt and CFQ-11 are found (p-values ranging from 0.05 to <0.01). PERMUTATIONS = 5000.

#### 3.4.2 ADt and FAt relationship with fatigue

We investigated the association between the ADt values and CFQ-11 scores in the COV+ group. ADt was significantly negatively correlated with CFQ-11 in 25,425 voxels, including the following regions: forceps major, forceps minor, left cingulum (hippocampus), bilateral cingulum (cingulate gyrus), anterior thalamic radiation, corticospinal tract, inferior fronto-occipital fasciculus, inferior longitudinal fasciculus, superior longitudinal fasciculus, superior longitudinal fasciculus (temporal part), and uncinate fasciculus (*p* < 0.05, TFCE-corrected; **Figure 3B**; **Supplementary Table 3**). FAt was significantly negatively correlated with CFQ-11 in 535 voxels, including the right superior longitudinal fasciculus, superior longitudinal fasciculus (temporal part), and the corticospinal tract (*p* < 0.05, TFCE-corrected; **Figure 3C**; **Supplementary Table 4**). We did not find any significant positive associations between ADt, FAt, and CFQ-11 scores.

### 3.5 Tract-wise analysis

#### 3.5.1 COV+ vs. COV- group comparison

In the tract-average analysis, the COV+ group had reduced FD in the left arcuate fasciculus and superior longitudinal fasciculus compared with the COV- group after adjusting for multiple comparisons (**Supplementary Table 5**). Reduced ADt in the right arcuate fasciculus and increased RDt in the left superior longitudinal fasciculus were observed in the COV+ group (**Supplementary Table 6**).

In along-tract statistics (tractometry), decreased FD was found in bundle sections within the arcuate fasciculus, cingulum, fornix, inferior fronto-occipital fasciculus, inferior longitudinal fasciculus, superior longitudinal fasciculus, uncinate fasciculus, corona radiata, corticospinal tract, and corpus callosum (posterior genu and rostral body) in the COV+ group as compared with the controls (**Figure 4, Supplementary Figure 1, Supplementary Table 7**). Only results with a p-value less than 0.05 and a t-value greater than the significance threshold were reported. Contrasting tract segments lay within clusters, that is, shared at least two or three significant neighboring bundle sections.

**Figure 4.**
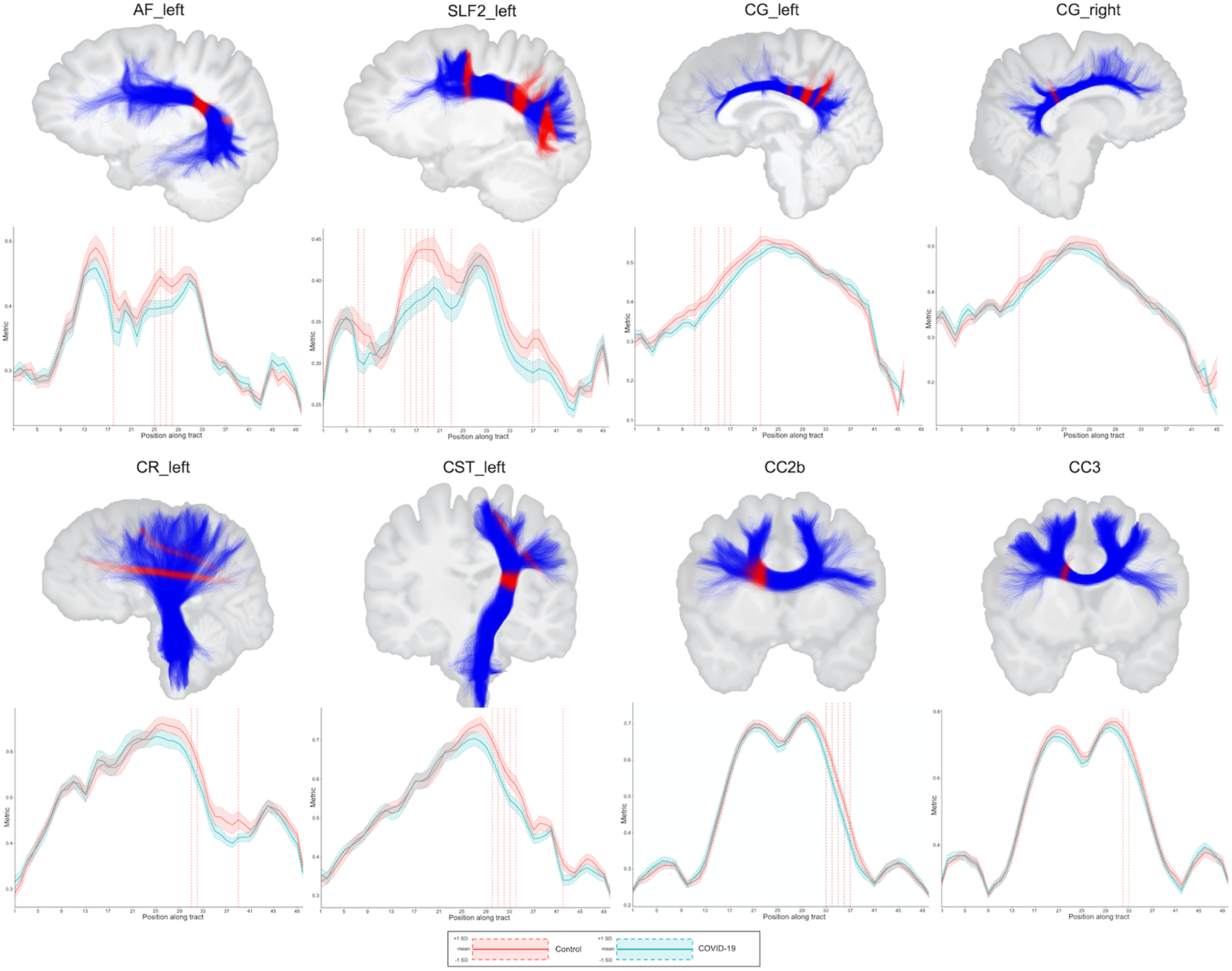
Results of the between-group comparisons on tractometry analysis (association, projection, and commissural tracts): COV- (red line) and COV+ (green line) groups. Only results with a *p* < 0.05 and a t-value greater than the significance threshold are reported. The dashed red line indicates whether the FD values of the COV+ group were significantly lower than those of the COV- group. The figures illustrate the tracts in blue and the regions with significance in red. Abbreviations: AF, arcuate fasciculus; CC, corpus callosum; CG, cingulum; CR, corona radiata; CST, corticospinal tract; SLF, superior longitudinal fasciculus; L, left; R, right; FD, fiber-specific apparent fiber density.

#### 3.5.2 Fiber density relationship with fatigue

In the COV+ group, tract-average FD values were negatively associated with CFQ-11 in the right corona radiata (*r* = -0.47, *p* = 0.007), left corona radiata (*r* = -0.63, *p* < 0.001), right corticospinal tract (*r* = -0.57, *p* < 0.001), left corticospinal tract (*r* = -0.52, *p* = 0.002), posterior mid-body of the corpus callosum (*r* = -0.42, *p* = 0.019), and the middle cerebellar peduncle (*r* = -0.43, *p* = 0.016). The tract-average FAt and ADt measurements in the corona radiata and corticospinal tract were negatively correlated with the total CFQ-11 score (**Figure 5, Supplementary Figure 2**). In the COV- group, the CFQ-11 and d-MRI metrics were not correlated.

**Figure 5.**
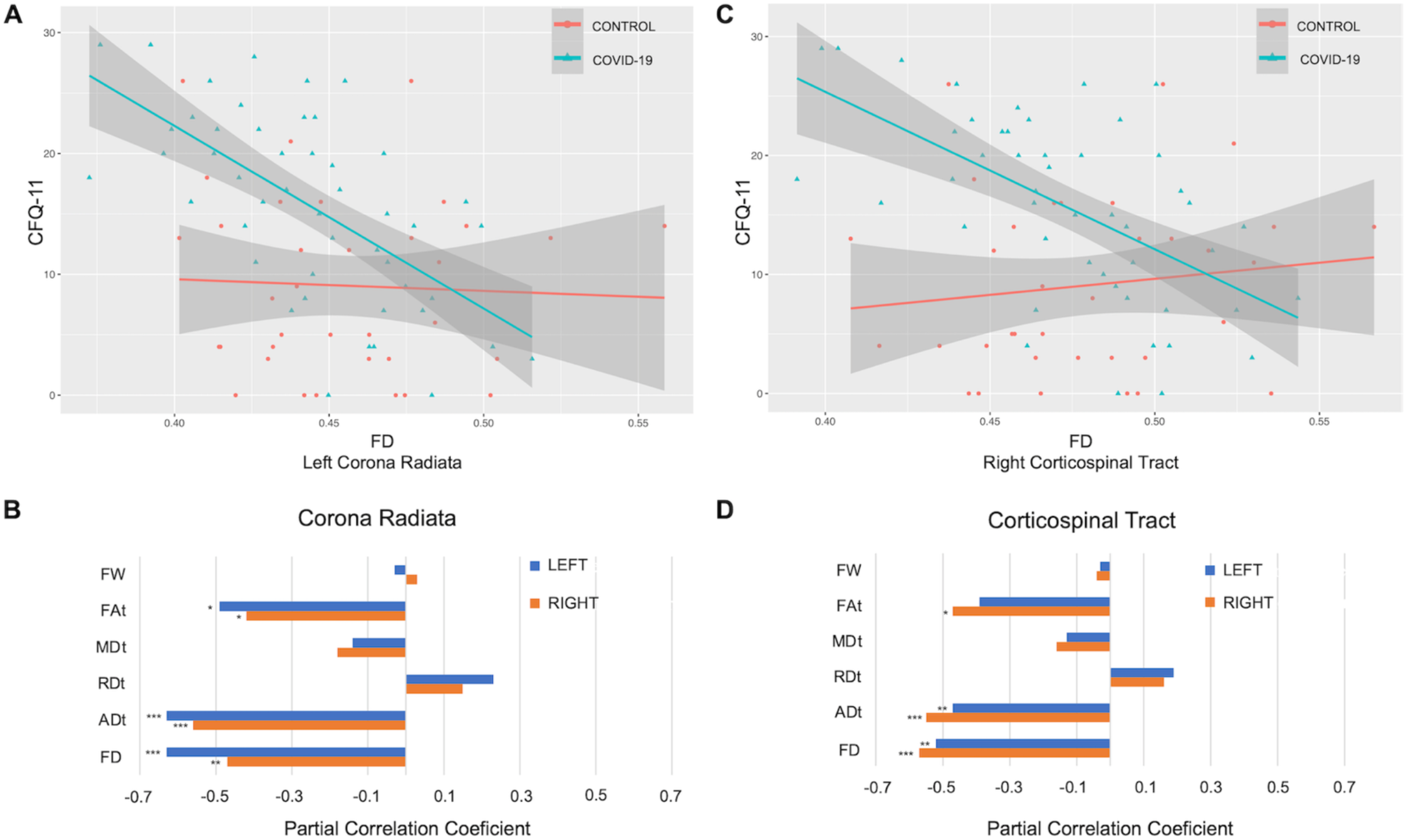
Associations between diffusion measures and total CFQ-11 score. (A–C) Associations between FD and total CFQ-11 in left corona radiata and right corticospinal tract. (B–D) Partial correlations between diffusion measures (average diffusion measure in the bundle) and total CFQ-11 score controlling for age, sex, and education were performed in the COV+ group. Partial correlation coefficient for each diffusion measure in the right and left bundles is reported as bar graphs (\**p* < 0.05; ** *p* < 0.01, \*\*\**p* < 0.001), with adjustment for multiple comparisons (FDR). Abbreviations: CFQ-11, Chalder Fatigue Scale; ADt, tissue axial diffusivity; FD, fiber-specific apparent fiber density; FAt, tissue fractional anisotropy; MDt, tissue mean diffusivity; RDt, tissue radial diffusivity; FW, free-water index; FDR, false discovery rate. Legend: * *p* < 0.05, ** *p* < 0.01, *** *p* < 0.001

## 4 DISCUSSION

Our study showed that patients with COVID-19 had microstructural changes in the WM at a mean follow-up of 3 months. Compared to the control group, the COV+ subjects had decreased fiber density in the association, projection, and commissural WM tracts, but no significant change in GM (cortical thickness or subcortical and limbic volumes). In the COV+ group, brain microstructural changes correlated with fatigue severity. Thus, the study provides evidence for possible brain substrates underlying symptoms caused by SARS-CoV-2 during medium-to long-term recovery in a predominantly non-hospitalized sample.

While DTI is the most frequently used d-MRI model for assessing WM integrity, it cannot resolve complex fiber geometries within the brain, which affects the quantification of related tissues. AFD, on the other hand, is a proxy for axonal degeneration because it reflects the apparent number of axons and is robust to crossing fibers.^20^ The FD for the fiber population within a single voxel was calculated using a fixel-based approach.^40^ We identified WM microstructural changes in the COV+ group: a reduction in FD in several bundles, such as the arcuate fasciculus, cingulum, fornix, inferior fronto-occipital fasciculus, inferior longitudinal fasciculus, superior longitudinal fasciculus, uncinate fasciculus, corona radiata, corticospinal tract, and corpus callosum, in comparison to the COV- group. Reduced FD suggests that intra-axonal volume reduction of specific fiber populations (e.g., axonal loss) might be a contributing factor to the pathological substrate for post-COVID symptoms and deserves further exploration. One caveat is that our MRI protocol is in the clinical range (single-shell, with low b-values of 800 m/s^2^). Thus, the interpretation of these findings must be cautious because the correlation between axon volumes and FD might not be as straightforward as if the MRI had a multi-shell DTI acquisition and high b-values (e.g., 3,000 m/s^2^).

There are limited publications on post-COVID brain microstructural changes. In studies performing DTI, increased FA was found in corona radiata, external capsule, and superior fronto-occipital fasciculus 3 months after SARS-CoV-2 infection,^41^ and decreased volume, length, and FA were found in association, projection, commissural, and limbic bundles in patients with mild-to-severe symptoms after COVID-19 convalescence.^12^ Our study identified relevant changes in FD but did not replicate some of these previously described DTI abnormalities. Multicompartment diffusion microstructure imaging in inpatients with subacute COVID-19 with neurological symptoms revealed widespread volume shifts compatible with vasogenic edema, affecting various white matter tracts. Redistribution with decreasing intra-axonal and extra-axonal volumes and increasing free water/CSF fraction was observed. These changes were associated with cognitive impairment and COVID-19-related changes in 18F-FDG PET imaging.^42^ Reduced axonal densities have been detected in patients after recovery from COVID-19, 1 year after infection in a sample consisting primarily of hospitalized patients.^43^ To our knowledge, there is no serial d-MRI study following up on a non-hospitalized sample of patients with milder COVID-19 forms. Such a study is essential to validate and assess the persistence of WM changes in post-COVID conditions.

In our study, the COV+ group had higher fatigue intensity than the control group. Fatigue is well documented in the post-COVID condition, even in non-hospitalized cases.^44^ There was a negative correlation between fatigue intensity and axonal integrity measures (FD, FAt, and ADt) in the projection bundles, cerebellar tracts, and corpus callosum. These results are comparable to those of patients with chronic fatigue syndrome (CFS).^45^ Patients with CFS have WM microstructural changes in the ascending and descending tracts of the brainstem as well as in the superior longitudinal fasciculus. Likewise, fatigue induced by multiple sclerosis associates with MD values across several WM tracts bilaterally (corona radiata, corticospinal tracts, and cerebellar peduncles).^46^ Patients with fatigue and cognitive difficulties following mild COVID-19 have altered excitability and neurotransmission within the motor cortex and deficits in executive functions and attention.^47^ Taken together, these data give insights into the mechanisms of post-infectious fatigue, which remains a poorly understood topic.^10^

A study including >80,000 participants (>12,000 patients with suspected COVID-19) identified a small but significant impairment in the global cognitive composite score for those infected with COVID-19.^8^ Negative effects on cognitive performance were stronger for those with respiratory difficulties, hospitalized, and placed on a ventilator. Our study found no difference in the MoCA global score and CANTAB cognitive performance between the COV+ and COV- groups. This lack of effect on sensitive electronic cognitive tests might be explained by a milder COVID severity in our cohort or due to decreased statistical power due to sample size.

The present study had some limitations. This was a cross-sectional study using non-probabilistic sampling, thus limiting the generalizability of the results. The patients were evaluated only once during the post-acute phase. The subjects were not serially evaluated at two distinct time points, a caveat that precludes inferences about the temporal dynamics of WM abnormalities. The diffusion parameters chosen (e.g., low b-values) may limit the analysis of HARDI metrics, but, on the other hand, may better reflect the context of a clinical protocol.

In summary, WM microstructure changes were detected by d-MRI in patients in the COVID-19 post-acute phase, providing new insights into the neurological damage directly or indirectly caused by SARS-CoV-2 infection. Further follow-up of these patients throughout the recovery process will contribute to the understanding of the pathophysiology of neurological damage and the possible sequelae generated by COVID-19.

## Data Availability

The anonymized dataset that supports these study findings is available upon reasonable request from the corresponding author from a qualified investigator if the intent is to increase reproducibility. The data were not publicly available because of privacy or ethical restrictions.

## Notes

### Competing Interest Statement

The authors have declared no competing interest.

### Funding Statement

L.S. is supported by the CAPES Foundation (Coordination for the Improvement of Higher Education Personnel).

### Author Declarations

This study was approved by the Research Ethics Committee of the University of Brasilia (Certificate of Ethical Appreciation Presentation - CAAE 31378820.1.2006.0030), and all subjects signed a consent form to participate.

## REFERENCES

1. 1. Nalbandian A, Sehgal K, Gupta A, et al. Post-acute COVID-19 syndrome. Nat Med. 2021;27(4):601–615. doi:10.1038/s41591-021-01283-z

2. CDC. Centers for Disease Control and Prevention. COVID-19. Long COVID or Post-COVID Conditions. https://www.cdc.gov/coronavirus/2019-ncov/long-term-effects/index.html. Accessed June 27, 2022.

3. Groff D, Sun A, Ssentongo AE, et al. Short-term and Long-term Rates of Postacute Sequelae of SARS-CoV-2 Infection: A Systematic Review. JAMA Netw Open. 2021;4(10):1–17. doi:10.1001/jamanetworkopen.2021.28568

4. Soriano JB, Murthy S, Marshall JC, Relan P, Diaz J V. A clinical case definition of post-COVID-19 condition by a Delphi consensus. Lancet Infect Dis. 2021;(October). doi:10.1016/s1473-3099(21)00703-9

5. Townsend L, Dyer AH, Jones K, et al. Persistent fatigue following SARS-CoV-2 infection is common and independent of severity of initial infection. PLoS One. 2020;15(11):e0240784. doi:10.1371/journal.pone.0240784

6. Becker JH, Lin JJ, Doernberg M, et al. Assessment of Cognitive Function in Patients after COVID-19 Infection. JAMA Netw Open. 2021;4(10):8–11. doi:10.1001/jamanetworkopen.2021.30645

7. Graham EL, Clark JR, Orban ZS, et al. Persistent neurologic symptoms and cognitive dysfunction in non-hospitalized Covid-19 “long haulers.” Ann Clin Transl Neurol. 2021;8(5):1073–1085. doi:10.1002/acn3.51350

8. Hampshire A, Trender W, Chamberlain SR, et al. Cognitive deficits in people who have recovered from COVID-19. EClinicalMedicine. 2021;39:101044. doi:10.1016/j.eclinm.2021.101044

9. Matos A De Mb, Dahy FE, de Moura Jvl, et al. Subacute Cognitive Impairment in Individuals With Mild and Moderate COVID-19: A Case Series. Front Neurol. 2021;12(August):1–8. doi:10.3389/fneur.2021.678924

10. Spudich S, Nath A. Nervous system consequences of COVID-19. Science (80-). 2022;375(6578):267–269. https://www.science.org/doi/10.1126/science.abm2052.

11. Thakur KT, Miller EH, Glendinning MD, et al. COVID-19 neuropathology at Columbia University Irving Medical Center/New York Presbyterian Hospital. Brain. 2021;144(9):2696–2708. doi:10.1093/brain/awab148

12. Qin Y, Wu J, Chen T, et al. Long-term microstructure and cerebral blood flow changes in patients recovered from COVID-19 without neurological manifestations. J Clin Invest. 2021;131(8). doi:10.1172/JCI147329

13. Douaud G, Lee S, Alfaro-Almagro F, et al. SARS-CoV-2 is associated with changes in brain structure in UK Biobank. Nature. 2022;604(7907):697–707. doi:10.1038/s41586-022-04569-5

14. Hosp JA, Dressing A, Blazhenets G, et al. Cognitive impairment and altered cerebral glucose metabolism in the subacute stage of COVID-19. Brain. 2021;144(4):1263–1276. doi:10.1093/brain/awab009

15. Basser PJ, Mattiello J, LeBihan D. MR diffusion tensor spectroscopy and imaging. Biophys J. 1994;66(1):259–267. doi:10.1016/S0006-3495(94)80775-1

16. Pierpaoli C, Jezzard P, Basser PJ, Barnett A, Di Chiro G. Diffusion tensor MR imaging of the human brain. Radiology. 1996;201(3):637–648. doi:10.1148/radiology.201.3.8939209

17. Pasternak O, Sochen N, Gur Y, Intrator N, Assaf Y. Free water elimination and mapping from diffusion MRI. Magn Reson Med. 2009;62(3):717–730. doi:10.1002/mrm.22055

18. Tournier J-D, Calamante F, Connelly A. Robust determination of the fibre orientation distribution in diffusion MRI: non-negativity constrained super-resolved spherical deconvolution. Neuroimage. 2007;35(4):1459–1472. doi:10.1016/j.neuroimage.2007.02.016

19. Raffelt DA, Tournier J-D, Smith RE, et al. Investigating white matter fibre density and morphology using fixel-based analysis. Neuroimage. 2017;144(Pt A):58-73. doi:10.1016/j.neuroimage.2016.09.029

20. Raffelt D, Tournier J-D, Rose S, et al. Apparent Fibre Density: a novel measure for the analysis of diffusion-weighted magnetic resonance images. Neuroimage. 2012;59(4):3976–3994. doi:10.1016/j.neuroimage.2011.10.045

21. NeuroCOVID-19. Brazilian Registry NeuroCovBr. https://www.neurocovbr.com/. Accessed February 1, 2022.

22. Jackson C. The Chalder Fatigue Scale (CFQ 11). Occup Med (Lond). 2015;65(1):86. doi:10.1093/occmed/kqu168

23. Chalder T, Berelowitz G, Pawlikowska T, et al. Development of a fatigue scale. J Psychosom Res. 1993;37(2):147–153. doi:10.1016/0022-3999(93)90081-p

24. Nøstdahl T, Bernklev T, Fredheim OM, Paddison JS, Raeder J. Defining the cut-off point of clinically significant postoperative fatigue in three common fatigue scales. Qual life Res an Int J Qual life Asp Treat care Rehabil. 2019;28(4):991–1003. doi:10.1007/s11136-018-2068-0

25. Cella M, Chalder T. Measuring fatigue in clinical and community settings. J Psychosom Res. 2010;69(1):17–22. doi:10.1016/j.jpsychores.2009.10.007

26. Nasreddine ZS, Phillips NA, Bédirian V, et al. The Montreal Cognitive Assessment, MoCA: a brief screening tool for mild cognitive impairment. J Am Geriatr Soc. 2005;53(4):695–699. doi:10.1111/j.1532-5415.2005.53221.x

27. Robbins TW, James M, Owen AM, Sahakian BJ, McInnes L, Rabbitt P. Cambridge Neuropsychological Test Automated Battery (CANTAB): a factor analytic study of a large sample of normal elderly volunteers. Dementia. 1994;5(5):266–281. doi:10.1159/000106735

28. Robbins TW, James M, Owen AM, et al. A study of performance on tests from the CANTAB battery sensitive to frontal lobe dysfunction in a large sample of normal volunteers: implications for theories of executive functioning and cognitive aging. Cambridge Neuropsychological Test Automated Batt. J Int Neuropsychol Soc. 1998;4(5):474–490. doi:10.1017/s1355617798455073

29. FreeSurfer Software Suite for Brain MRI Analysis. Athinoula A. Martinos Center for Biomedical Imaging. http://surfer.nmr.mgh.harvard.edu. Accessed February 1, 2022.

30. Theaud G, Houde J-C, Boré A, Rheault F, Morency F, Descoteaux M. TractoFlow: A robust, efficient and reproducible diffusion MRI pipeline leveraging Nextflow & Singularity. Neuroimage. 2020;218:116889. doi:10.1016/j.neuroimage.2020.116889

31. Smith SM, Jenkinson M, Johansen-Berg H, et al. Tract-based spatial statistics: voxelwise analysis of multi-subject diffusion data. Neuroimage. 2006;31(4):1487–1505. doi:10.1016/j.neuroimage.2006.02.024

32. Garyfallidis E, Côté M-A, Rheault F, et al. Recognition of white matter bundles using local and global streamline-based registration and clustering. Neuroimage. 2018;170:283–295. doi:10.1016/j.neuroimage.2017.07.015

33. Rheault F. Analyse et reconstruction de faisceaux de la matière blanche. 2020. https://savoirs.usherbrooke.ca/handle/11143/17255.

34. Rheault F, St-Onge E, Sidhu J, et al. Bundle-specific tractography with incorporated anatomical and orientational priors. Neuroimage. 2019;186:382–398. doi:10.1016/j.neuroimage.2018.11.018

35. Cousineau M, Jodoin P-M, Morency FC, et al. A test-retest study on Parkinson’s PPMI dataset yields statistically significant white matter fascicles. NeuroImage Clin. 2017;16:222–233. doi:10.1016/j.nicl.2017.07.020

36. Yeatman JD, Dougherty RF, Myall NJ, Wandell BA, Feldman HM. Tract profiles of white matter properties: automating fiber-tract quantification. PLoS One. 2012;7(11):e49790. doi:10.1371/journal.pone.0049790

37. A Nextflow pipeline for diffusion MRI quality check (dmriqc_flow). Sherbrooke Connectivity Imaging Lab. https://github.com/scilus/dmriqc_flow. Accessed February 1, 2022.

38. Hua K, Zhang J, Wakana S, et al. Tract probability maps in stereotaxic spaces: analyses of white matter anatomy and tract-specific quantification. Neuroimage. 2008;39(1):336–347. doi:10.1016/j.neuroimage.2007.07.053

39. Nichols TE, Holmes AP. Nonparametric permutation tests for functional neuroimaging: a primer with examples. Hum Brain Mapp. 2002;15(1):1–25. doi:10.1002/hbm.1058

40. Raffelt DA, Smith RE, Ridgway GR, et al. Connectivity-based fixel enhancement: Whole-brain statistical analysis of diffusion MRI measures in the presence of crossing fibres. Neuroimage. 2015;117:40–55. doi:10.1016/j.neuroimage.2015.05.039

41. Lu Y, Li X, Geng D, et al. Cerebral Micro-Structural Changes in COVID-19 Patients – An MRI-based 3-month Follow-up Study: A brief title: Cerebral Changes in COVID-19. EClinicalMedicine. 2020;25(2):100484. doi:10.1016/j.eclinm.2020.100484

42. Rau A, Schroeter N, Blazhenets G, et al. Widespread white matter oedema in subacute COVID-19 patients with neurological symptoms. Brain. June 2022. doi:10.1093/brain/awac045

43. Huang S, Zhou Z, Yang D, et al. Persistent white matter changes in recovered COVID-19 patients at the 1-year follow-up. Brain. December 2021. doi:10.1093/brain/awab435

44. El Sayed S, Shokry D, Gomaa SM. Post-COVID-19 fatigue and anhedonia: A cross-sectional study and their correlation to post-recovery period. Neuropsychopharmacol reports. 2021;41(1):50–55. doi:10.1002/npr2.12154

45. Thapaliya K, Marshall-Gradisnik S, Staines D, Barnden L. Diffusion tensor imaging reveals neuronal microstructural changes in myalgic encephalomyelitis/chronic fatigue syndrome. Eur J Neurosci. 2021;54(6):6214–6228. doi:10.1111/ejn.15413

46. Novo AM, Batista S, Alves C, et al. The neural basis of fatigue in multiple sclerosis: A multimodal MRI approach. Neurol Clin Pract. 2018;8(6):492–500. doi:10.1212/CPJ.0000000000000545

47. Ortelli P, Ferrazzoli D, Sebastianelli L, et al. Altered motor cortex physiology and dysexecutive syndrome in patients with fatigue and cognitive difficulties after mild COVID-19. Eur J Neurol. February 2022. doi:10.1111/ene.15278

