## Supplementary material 1 for "Brain microstructural changes and fatigue after COVID-19"

### **Supplementary Table 1.** Key cognitive outcome measures

| Cognitive measure name | Abbreviation | Brief description |
|---|---|---|
| Spatial working memory, between-errors total | SWMBE | The number of times a subject incorrectly revisits a box in which a token had already been found. |
| Spatial working memory, strategy | SWMS | The number of times a subject starts a new search pattern from the same box they started with earlier. |
| One-Touch Stockings of Cambridge, Problems solved on the first choice | OTSPSFC | The total number of assessed trials where the subject chose the correct answer on their first attempt. |
| One-Touch Stockings of Cambridge, The average latency for the first choice | OTSMDLFC | Average latency, measured from the appearance of the stocking balls to the first box choice, was taken by the subject. |
| One-Touch Stockings of Cambridge, Average of choices to correct | OTSMCC | The average number of box selections that the subject took before choosing the correct box. |
| One-Touch Stockings of Cambridge, Average latency to correct | OTSMLC | Average latency, measured from the appearance of the stocking balls to the choice of the correct box, was taken by the subject. |
| Paired Associates Learning, Adjusted total errors | PALTEA | The number of times the subject chose the wrong box for a stimulus in assessment problems plus an adjustment to the estimated number of errors they would have made in any problems, trials, and recalls. |
| Paired Associates Learning, Memory score on the first attempt | PALFAMS | The number of times the subject chooses the correct box on the first attempt to remember its location. |
| Paired Associates Learning, Mean errors to success | PALMETS | The average number of attempts to complete a test stage |
| Pattern Recognition Memory, the percentage of correct answers, immediate | PRMPCI | Number of patterns correctly selected by the subject in the immediate "forced-choice" condition, expressed as a percentage |
| Pattern Recognition Memory, the percentage of correct answers, delayed | PRMPCD | The number of patterns correctly selected by the subject in the delayed "forced-choice" condition expressed as a percentage. |
| Verbal Recognition Memory, Immediate verbal recognition, total correct answers | VRMIRTC | Total target words correctly recognized in the immediate recognition phase, plus the total number of distracting words that the subject correctly rejects. |
| Verbal Recognition Memory, Delayed verbal recognition, total correct answers | VRMDRTC | The total number of target words correctly recognized in the delayed recognition phase, plus the total number of distracting words that the subject correctly rejects. |
| Verbal Recognition Memory, Free verbal recall, different stimuli | VRMFRDS | Total number of different stimuli that are correctly remembered by the subject in the phase of free word list recall |
| Single-choice median reaction time | RTISMDRT | Median time taken to release the button after presenting a target stimulus. Measured in milliseconds |
| Single-choice median movement time | RTISMDMT | Median time taken to release the button and select the target stimulus after it flashes yellow on the screen. It is measured in milliseconds. |
| Five-choice median reaction time | RTIFMDRT | Median time taken to release the button after the target stimulus flashes yellow on the screen. Measured in milliseconds |
| Five-choice median movement time | RTIFMDMT | Median time taken to release the button and select the target stimulus after it flashes yellow on the screen. It is measured in milliseconds. |

**Supplementary Table 2.** TBSS results significantly reduced AFDtotal in the COV+ group compared to the COV- group. The anatomical locations were determined by referring to the JHU white-matter tractography atlas. The value after each region indicates the percentage probability of the cluster belonging to the given atlas label.

| Cluster | White matter tract | p-value | Voxels | MNI (x y z) (mm) | | |
|---|---|---|---|---|---|---|
| corrected p-value < 0.05 | | | | | | |
| AFDtotal / COV- > COV+ | | | | | | |
| 1 | Anterior thalamic radiation L:0.3928<br>Corticospinal tract L:4.5856<br>Cingulum (cingulate gyrus) L:0.0829<br>Inferior fronto-occipital fasciculus L:0.0108<br>Inferior longitudinal fasciculus L:0.0775<br>Superior longitudinal fasciculus L:12.4739<br>Superior longitudinal fasciculus (temporal part) L:4.3892 | 0.026 | 4,515 | -35 | -19 | 34 |

Abbreviations: TBSS, tract-based spatial statistics; MNI, Montreal Neurological Institute; L, an abbreviation for the left hemisphere, R, abbreviation for the right hemisphere; AFDtotal, voxel-wise apparent fiber density.

**Supplementary Table 3.** TBSS results show a negative correlation between ADt and CFQ-11 in the COV+ group. The anatomical locations were determined by referring to the JHU white-matter tractography atlas. The value after each region indicates the percentage probability of the cluster belonging to the given atlas label.

| Cluster | White matter tract | p-value | Voxels | MNI (x y z) (mm) | | |
|---|---|---|---|---|---|---|
| corrected p-value < 0.05 | | | | | | |
| Negative correlation between ADt and CFQ-11 | | | | | | |
| 1 | Anterior thalamic radiation L:7.3333<br>Cingulum (cingulate gyrus) L:1.8889<br>Forceps minor:47.8889<br>Inferior fronto-occipital fasciculus L:1.6667<br>Uncinate fasciculus L:0.3333 | 0.044 | 53 | -19 | 48 | 10 |
| 2 | Anterior thalamic radiation L:6.2500<br>Cingulum (cingulate gyrus) L:2.6250<br>Forceps minor:32.8750<br>Inferior fronto-occipital fasciculus L:1.1250 | 0.041 | 60 | -17 | 45 | 20 |
| 3 | Cingulum (cingulate gyrus) L:6.9444<br>Forceps minor:32.7778 | 0.036 | 144 | -12 | 23 | 19 |
| 4 | Anterior thalamic radiation R:10.4706<br>Cingulum (cingulate gyrus) R:0.0294<br>Forceps minor:42.7647<br>Inferior fronto-occipital fasciculus R:1.4118<br>Uncinate fasciculus R:0.0882 | 0.034 | 328 | 18 | 52 | 15 |
| 5 | Anterior thalamic radiation L:1.6008<br>Anterior thalamic radiation R:1.3787<br>Corticospinal tract L:2.3020<br>Corticospinal tract R:2.2288<br>Cingulum (cingulate gyrus) L:0.2892<br>Cingulum (cingulate gyrus) R:0.0662<br>Cingulum (hippocampus) L:0.0006<br>Forceps major:0.2496<br>Forceps minor:0.0256<br>Inferior fronto-occipital fasciculus L:1.2630<br>Inferior fronto-occipital fasciculus R:1.2924<br>Inferior longitudinal fasciculus L:0.3944<br>Inferior longitudinal fasciculus R:0.1521<br>Superior longitudinal fasciculus L:2.1668<br>Superior longitudinal fasciculus R:1.7983<br>Uncinate fasciculus L:0.4263<br>Uncinate fasciculus R:0.2793<br>Superior longitudinal fasciculus (temporal part) L:0.8453<br>Superior longitudinal fasciculus (temporal part) R:0.5638 | <0.001 | 24,840 | 14 | -18 | -14 |

Abbreviations: TBSS, tract-based spatial statistics; MNI, Montreal Neurological Institute; L, an abbreviation for the left hemisphere, R, abbreviation for the right hemisphere; ADt, tissue axial diffusivity.

**Supplementary Table 4.** TBSS results show a negative correlation between FAt and CFQ-11 in the COV+ group. The anatomical locations were determined by referring to the JHU white-matter tractography atlas. The value after each region indicates the percentage probability of the cluster belonging to the given atlas label.

| Cluster | White matter tract | p-value | Voxels | MNI (x y z) (mm) | | |
|---|---|---|---|---|---|---|
| corrected p-value < 0.05 | | | | | | |
| Negative correlation between FAt and CFQ-11 | | | | | | |
| 1 | Corticospinal tract R:27.0000 | 0.049 | 13 | 21 | -33 | 51 |
| 2 | Corticospinal tract R:10.0806<br>Superior longitudinal fasciculus R:16.4677<br>Superior longitudinal fasciculus (temporal part) R:5.1290 | 0.036 | 522 | 27 | -16 | 30 |

Abbreviations: TBSS, tract-based spatial statistics; MNI, Montreal Neurological Institute; R, abbreviation for the right hemisphere; FAt, tissue fractional anisotropy.

**Supplementary Table 5.** Comparison of tract-average FW, FAt, and FD measures between control and COVID-19 groups, adjusting for multiple comparisons (FDR). Age and sex were included as covariates.

| | | FW | | | FAt | | | FD | | |
|---|---|---|---|---|---|---|---|---|---|---|
| **Tract** | N | statistic | p-value | adj p | statistic | p-value | adj p | statistic | p-value | adj p |
| ***Association Tracts*** | | | | | | | | | | |
| AF (L) | 83 | 1.3404 | 0.1840 | ns | -1.4041 | 0.1642 | ns | -2.7668 | 0.0070 | **0.0176** |
| AF (R) | 83 | 0.0777 | 0.9382 | ns | -1.4549 | 0.1497 | ns | -1.1706 | 0.2453 | ns |
| CG (L) | 83 | 0.4322 | 0.6668 | ns | -1.1128 | 0.2692 | ns | -2.0862 | 0.0402 | ns |
| CG (R) | 83 | 0.6208 | 0.5365 | ns | -0.4699 | 0.6397 | ns | -1.2945 | 0.1993 | ns |
| FX (L) | 83 | -1.2586 | 0.2119 | ns | 0.2182 | 0.8279 | ns | 0.0838 | 0.9334 | ns |
| FX (R) | 83 | -1.0000 | 0.2883 | ns | -0.3594 | 0.7203 | ns | -0.8627 | 0.3909 | ns |
| IFOF (L) | 83 | 0.2981 | 0.7664 | ns | -0.5843 | 0.5607 | ns | -1.1916 | 0.2370 | ns |
| IFOF (R) | 83 | -1.0898 | 0.2791 | ns | -0.5276 | 0.5992 | ns | -0.8180 | 0.4158 | ns |
| ILF (L) | 83 | 0.4894 | 0.6259 | ns | -0.3538 | 0.7245 | ns | -0.8028 | 0.4245 | ns |
| ILF (R) | 83 | -1.2750 | 0.2060 | ns | -0.3735 | 0.7098 | ns | 0.7067 | 0.4818 | ns |
| SLF1 (L) | 82 | 0.7935 | 0.4299 | ns | -0.2917 | 0.7713 | ns | -1.1157 | 0.2680 | ns |
| SLF1 (R) | 82 | 0.1050 | 0.9167 | ns | -0.0680 | 0.9460 | ns | -1.0808 | 0.2831 | ns |
| SLF2 (L) | 83 | 0.7492 | 0.4560 | ns | -1.9440 | 0.0555 | ns | -4.1441 | 0.0001 | **0.0003** |
| SLF2 (R) | 83 | 0.3672 | 0.7145 | ns | -0.5532 | 0.5817 | ns | -0.9890 | 0.3257 | ns |
| SLF3 (L) | 83 | 1.3674 | 0.1754 | ns | -1.2180 | 0.2268 | ns | -2.4932 | 0.0147 | **0.0338** |
| SLF3 (R) | 83 | 0.0050 | 0.9960 | ns | -0.6692 | 0.5053 | ns | -1.1172 | 0.2673 | ns |
| UF (L) | 83 | 1.1968 | 0.2350 | ns | -0.8132 | 0.4185 | ns | -0.9967 | 0.3219 | ns |
| UF (R) | 83 | 0.8023 | 0.4248 | ns | -0.6405 | 0.5237 | ns | -2.0819 | 0.0406 | ns |
| ***Projection Tracts*** | | | | | | | | | | |
| CR (L) | 83 | 1.0810 | 0.2830 | ns | -1.7953 | 0.0764 | ns | -1.7566 | 0.0829 | ns |
| CR (R) | 83 | 0.6288 | 0.5313 | ns | -0.9705 | 0.3348 | ns | -0.9170 | 0.3619 | ns |
| CST (L) | 83 | 0.6414 | 0.5231 | ns | -1.7264 | 0.0882 | ns | -1.7133 | 0.0906 | ns |
| CST (R) | 83 | 1.1909 | 0.2373 | ns | -0.6567 | 0.5133 | ns | -0.7963 | 0.4282 | ns |
| OR (L) | 83 | 0.4395 | 0.6615 | ns | -1.1504 | 0.2534 | ns | -1.5422 | 0.1270 | ns |
| OR (R) | 83 | -1.4913 | 0.1399 | ns | -1.1942 | 0.2360 | ns | -0.2631 | 0.7932 | ns |
| ***Corpus Callosum*** | | | | | | | | | | |
| Rostrum | 83 | -0.0750 | 0.9404 | ns | -0.7661 | 0.4459 | ns | -1.2245 | 0.2244 | ns |
| Genu (A) | 83 | 0.0628 | 0.9501 | ns | -0.4008 | 0.6896 | ns | -0.6928 | 0.4905 | ns |
| Genu (P) | 83 | 0.2716 | 0.7867 | ns | -0.3049 | 0.7613 | ns | -0.5060 | 0.6143 | ns |
| Rostral Body | 83 | 0.4940 | 0.6226 | ns | -0.3849 | 0.7014 | ns | -1.0515 | 0.2962 | ns |
| Mid-Body (A) | 83 | 0.8628 | 0.3909 | ns | -0.8193 | 0.4151 | ns | -1.3758 | 0.1728 | ns |
| Mid-Body (P) | 83 | 1.7139 | 0.0905 | ns | -1.1160 | 0.2678 | ns | -1.8233 | 0.0720 | ns |
| Isthmus | 83 | 0.6921 | 0.4909 | ns | -0.1809 | 0.8569 | ns | -0.9417 | 0.3492 | ns |
| Splenium | 83 | 0.8067 | 0.4223 | ns | -2.0815 | 0.0406 | ns | -1.8938 | 0.0619 | ns |
| ***Cerebellar Tracts*** | | | | | | | | | | |
| MCP | 83 | -0.5661 | 0.5729 | ns | -1.6840 | 0.0961 | ns | 0.7375 | 0.4630 | ns |
| SCP (L) | 83 | -0.3679 | 0.7139 | ns | -0.7190 | 0.4742 | ns | 0.0006 | 0.9995 | ns |
| SCP (R) | 83 | 0.1060 | 0.9158 | ns | 0.2269 | 0.8211 | ns | 0.6936 | 0.4900 | ns |

Abbreviations: AF, arcuate fasciculus; CG, cingulum; IFOF, inferior frontal occipital fasciculus; ILF, inferior longitudinal fasciculus; SLF, superior longitudinal fasciculus; UF, uncinate fasciculus; CR, corona radiata; CST, corticospinal tract; OR, optic radiation; SCP, superior cerebellar peduncle; MCP, middle cerebellar peduncle; A, anterior; P, posterior; L, left; R, right; FDR, false discovery rate; FW, free-water; FAt, tissue fractional anisotropy; FD, fiber-specific apparent fiber density; ns, not significant; adj, adjusted. Significant p values at a threshold of $p < 0.05$ are indicated in bold font.

**Supplementary Table 6.** Comparison of tract-average MDt, RDt, and ADt measures between control and COVID-19 groups, adjusting for multiple comparisons (FDR). Age and sex were included as covariates.

| | | **MDt** | | | **RDt** | | | **ADt** | | |
|---|---|---|---|---|---|---|---|---|---|---|
| **Tract** | N | statistic | p-value | adj p | statistic | p-value | adj p | statistic | p-value | adj p |
| ***Association Tracts*** | | | | | | | | | | |
| AF (L) | 83 | 2.1929 | 0.0313 | ns | 2.0792 | 0.0408 | ns | 1.5321 | 0.1295 | ns |
| AF (R) | 83 | -0.8679 | 0.3881 | ns | 0.2660 | 0.7909 | ns | -2.6631 | 0.0094 | **0.0285** |
| CG (L) | 83 | 1.3288 | 0.1878 | ns | 1.4035 | 0.1644 | ns | 0.2878 | 0.7742 | ns |
| CG (R) | 83 | 0.0564 | 0.9552 | ns | 0.2337 | 0.8158 | ns | -0.2846 | 0.7767 | ns |
| FX (L) | 83 | -1.8241 | 0.0719 | ns | -1.6269 | 0.1077 | ns | -1.8417 | 0.0693 | ns |
| FX (R) | 83 | -0.7085 | 0.4807 | ns | -0.7633 | 0.4475 | ns | -0.6263 | 0.5329 | ns |
| IFOF (L) | 83 | 0.6257 | 0.5333 | ns | 0.7522 | 0.4541 | ns | -0.1598 | 0.8734 | ns |
| IFOF (R) | 83 | -1.1838 | 0.2401 | ns | -0.2003 | 0.8418 | ns | -1.9240 | 0.0580 | ns |
| ILF (L) | 83 | 0.6882 | 0.4934 | ns | 0.4707 | 0.6391 | ns | 0.5611 | 0.5763 | ns |
| ILF (R) | 83 | -1.1994 | 0.2340 | ns | -0.5102 | 0.6113 | ns | -1.4912 | 0.1399 | ns |
| SLF1 (L) | 82 | 1.6094 | 0.1116 | ns | 1.0080 | 0.3166 | ns | 1.8319 | 0.0708 | ns |
| SLF1 (R) | 82 | -0.1877 | 0.8516 | ns | -0.2141 | 0.8310 | ns | -0.0399 | 0.9683 | ns |
| SLF2 (L) | 83 | 2.0859 | 0.0402 | ns | 2.3632 | 0.0206 | **0.0430** | 0.8378 | 0.4047 | ns |
| SLF2 (R) | 83 | -0.6642 | 0.5085 | ns | -0.0191 | 0.9848 | ns | -1.4407 | 0.1536 | ns |
| SLF3 (L) | 83 | 2.1166 | 0.0374 | ns | 1.9132 | 0.0593 | ns | 1.6985 | 0.0933 | ns |
| SLF3 (R) | 83 | -0.8427 | 0.4019 | ns | -0.0810 | 0.9357 | ns | -1.9718 | 0.0521 | ns |
| UF (L) | 83 | 1.1821 | 0.2407 | ns | 1.1402 | 0.2577 | ns | 0.4096 | 0.6832 | ns |
| UF (R) | 83 | 0.0332 | 0.9736 | ns | 0.5227 | 0.6027 | ns | -0.8499 | 0.3979 | ns |
| ***Projection Tracts*** | | | | | | | | | | |
| L CR | 83 | 1.4985 | 0.1380 | ns | 1.9211 | 0.0583 | ns | 0.0287 | 0.9772 | ns |
| R CR | 83 | -0.5312 | 0.5968 | ns | 0.3399 | 0.7348 | ns | -1.3631 | 0.1767 | ns |
| L CST | 83 | 1.0825 | 0.2823 | ns | 1.6493 | 0.1031 | ns | -0.1215 | 0.9036 | ns |
| R CST | 83 | -0.3254 | 0.7458 | ns | 0.2484 | 0.8045 | ns | -0.8342 | 0.4067 | ns |
| L OR | 83 | 1.0443 | 0.2996 | ns | 1.1044 | 0.2728 | ns | 0.3050 | 0.7612 | ns |
| R OR | 83 | -0.8172 | 0.4163 | ns | 0.1995 | 0.8424 | ns | -1.9637 | 0.0531 | ns |
| ***Corpus Callosum*** | | | | | | | | | | |
| Rostrum | 83 | -0.0924 | 0.9266 | ns | 0.5232 | 0.6023 | ns | -0.9860 | 0.3271 | ns |
| Genu (A) | 83 | -0.0339 | 0.9730 | ns | 0.4381 | 0.6625 | ns | -0.6113 | 0.5428 | ns |
| Genu (P) | 83 | 0.4856 | 0.6286 | ns | 0.6013 | 0.5494 | ns | -0.0025 | 0.9980 | ns |
| Rostral Body | 83 | 0.4179 | 0.6771 | ns | 0.5382 | 0.5920 | ns | -0.0482 | 0.9617 | ns |
| Mid-body (A) | 83 | 0.4803 | 0.6323 | ns | 0.7926 | 0.4304 | ns | -0.2377 | 0.8128 | ns |
| Mid-body (P) | 83 | 1.4107 | 0.1623 | ns | 1.4722 | 0.1449 | ns | 0.5678 | 0.5717 | ns |
| Isthmus | 83 | 1.2853 | 0.2024 | ns | 0.6038 | 0.5477 | ns | 1.3354 | 0.1856 | ns |
| Splenium | 83 | 1.0173 | 0.3121 | ns | 1.8827 | 0.0634 | ns | -0.6168 | 0.5391 | ns |
| ***Cerebellar Tracts*** | | | | | | | | | | |
| MCP | 83 | 0.1954 | 0.8456 | ns | 1.1784 | 0.2422 | ns | -1.0125 | 0.3144 | ns |
| L SCP | 83 | -0.1880 | 0.8514 | ns | 0.7507 | 0.4551 | ns | -0.6707 | 0.5043 | ns |
| R SCP | 83 | -0.1208 | 0.9041 | ns | -0.0771 | 0.9387 | ns | -0.0611 | 0.9514 | ns |

Abbreviations: AF, arcuate fasciculus; CG, cingulum; IFOF, inferior frontal occipital fasciculus; ILF, inferior longitudinal fasciculus; SLF, superior longitudinal fasciculus; UF, uncinate fasciculus; CR, corona radiata; CST, corticospinal tract; OR, optic radiation; SCP, superior cerebellar peduncle; MCP, middle cerebellar peduncle; A, anterior; P, posterior; L, left; R, right; FDR, false discovery rate; MDt, tissue mean diffusivity; RDt, tissue radial diffusivity; ADt, tissue axial diffusivity; ns, not significant; adj, adjusted.

**Supplementary Table 7.** Tract profiles showing significant between-group differences. The t- and p-values, and the corrected significance threshold computed from permutations (threshold) are shown. The difference is significant when the p-value is lower than 0.05, and the t-value in absolute value is greater than the significance threshold. It is a significant increase in control group versus COVID-19 when the t-value is positive, and a significant decrease when it is negative.

**[The table is in a separate file]**

**Supplementary Figure 1.** Results of the between-group comparisons on tractometry analysis (association tracts): control (red line) and COVID-19 (green line) groups. Only results with a p<0.05 and a t-value greater than the significance threshold are reported. The dashed red line indicates whether the FD values of the COVID-19 group were significantly lower than those of the control group.

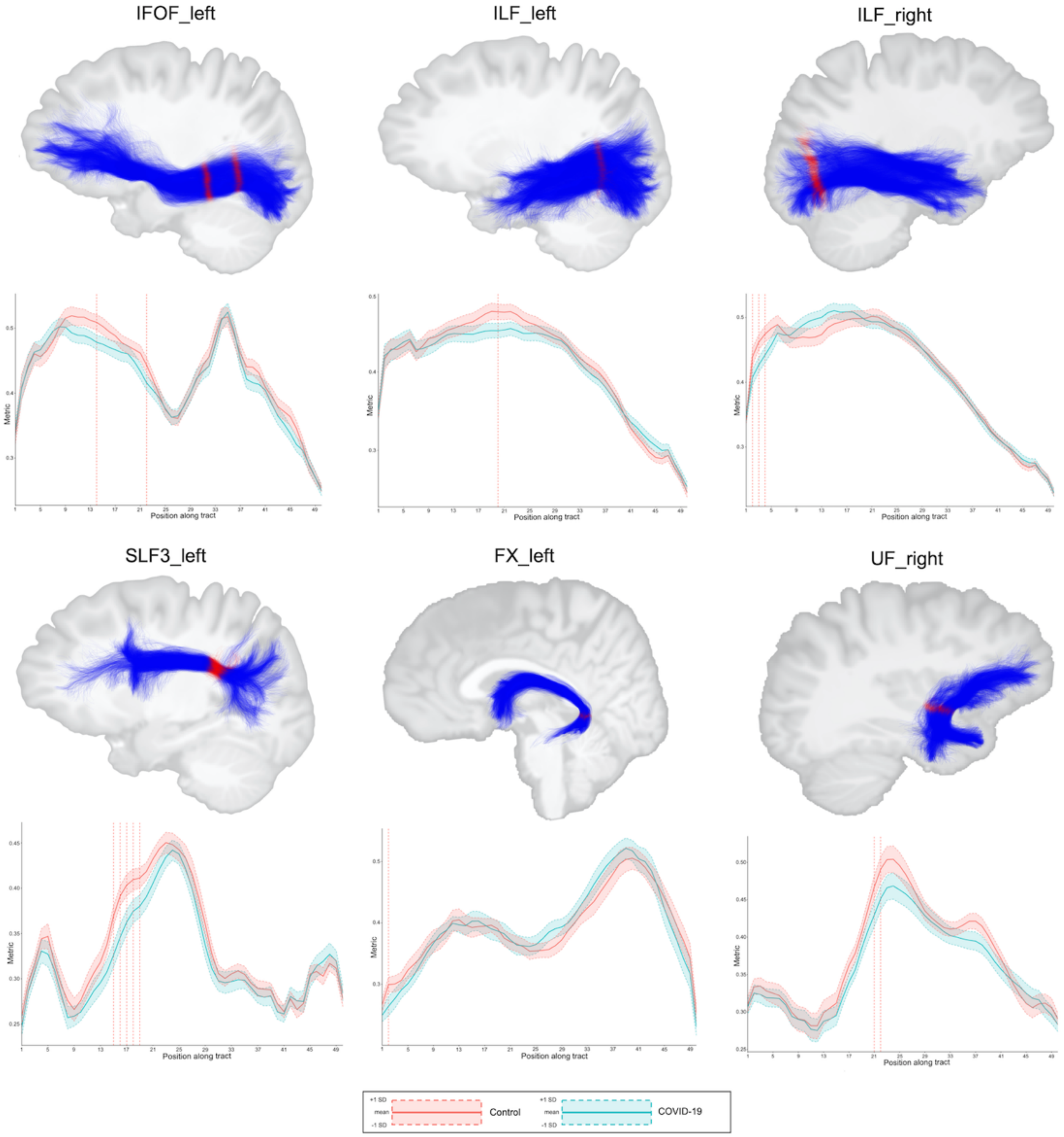


Abbreviations: FX, fornix; IFOF, inferior frontal occipital fasciculus; ILF, inferior longitudinal fasciculus; SLF, superior longitudinal fasciculus; FD, fiber-specific apparent fiber density.

The figures illustrate the tracts in blue and the regions with significance in red.

**Supplementary Figure 2.** Associations between diffusion measures and total CFQ-11 score in the COV+ group. (A–C) Associations between FD and total CFQ-11 in the posterior mid-body of the corpus callosum and middle cerebellar peduncle. (B–D) Partial correlations between diffusion measures (average diffusion measure in the bundle) and total CFQ-11 score controlling for age, sex, and education were performed. The partial correlation coefficient for each diffusion measure in the bundles is reported as bar graphs (*p<0.05), with adjustment for multiple comparisons (FDR).

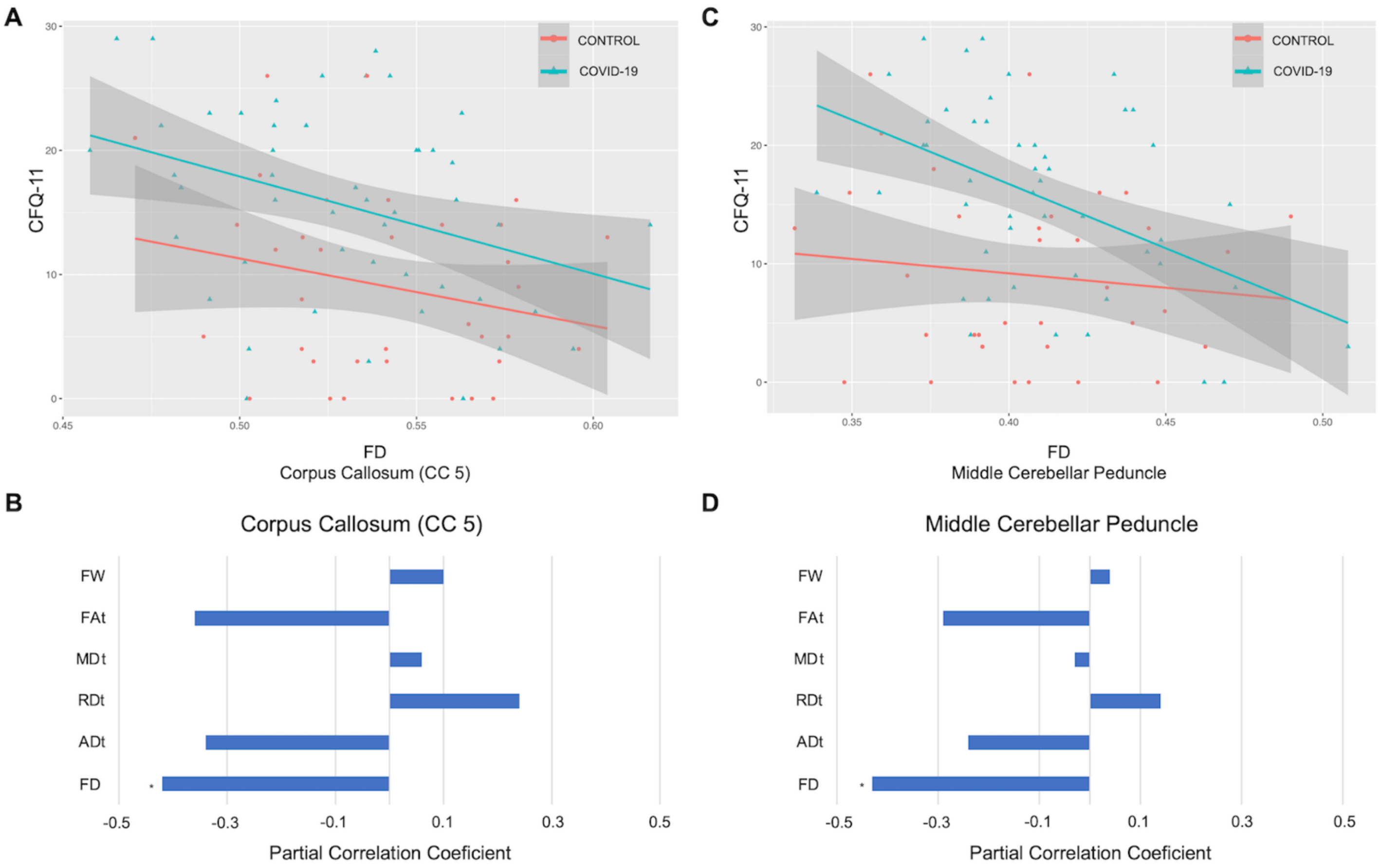


Abbreviations: CFQ-11, Chalder Fatigue Scale; ADt, tissue axial diffusivity; FD, fiber-specific apparent fiber density; FAt, tissue fractional anisotropy; MDt, tissue mean diffusivity; RDt, tissue radial diffusivity; FW, free-water index; CC 5, posterior mid-body of the corpus callosum; FDR = false discovery rate.

Legend: * p < 0.05
