## Supplementary material 2 for "Brain microstructural changes and fatigue after COVID-19"

**Supplementary Table 7.** Tract profiles showing significant between-group differences. The t- and p-values, and the corrected significance threshold computed from permutations (threshold) are shown. The difference is significant when the p-value is lower than 0.05, and the t-value in absolute value is greater than the significance threshold. It is a significant increase in control group versus COVID-19 when the t-value is positive, and a significant decrease when it is negative.

| metric | tract | section | t | threshold | pvalue | sig |
|---|---|---|---|---|---|---|
| AF_L_FD | AF_L | 1 | -0.742335815 | 2.351931482 | 0.460040484 | 0 |
| AF_L_FD | AF_L | 2 | -1.126797122 | 2.386766367 | 0.263400664 | 0 |
| AF_L_FD | AF_L | 3 | 0.414970919 | 2.428177199 | 0.67931922 | 0 |
| AF_L_FD | AF_L | 4 | 1.55821804 | 2.362300545 | 0.124945016 | 0 |
| AF_L_FD | AF_L | 5 | -0.447385838 | 2.412234132 | 0.656013945 | 0 |
| AF_L_FD | AF_L | 6 | -0.817365517 | 2.400507978 | 0.416141609 | 0 |
| AF_L_FD | AF_L | 7 | -0.518396526 | 2.400245647 | 0.605796543 | 0 |
| AF_L_FD | AF_L | 8 | -0.566253728 | 2.385615023 | 0.572994361 | 0 |
| AF_L_FD | AF_L | 9 | 0.112277073 | 2.374687299 | 0.910891122 | 0 |
| AF_L_FD | AF_L | 10 | 0.633254294 | 2.413079532 | 0.528452836 | 0 |
| AF_L_FD | AF_L | 11 | 0.58478113 | 2.316325757 | 0.560710392 | 0 |
| AF_L_FD | AF_L | 12 | 0.069132211 | 2.37314397 | 0.945118763 | 0 |
| AF_L_FD | AF_L | 13 | 0.429174415 | 2.370302732 | 0.669308096 | 0 |
| AF_L_FD | AF_L | 14 | 1.552320841 | 2.35449507 | 0.125372967 | 0 |
| AF_L_FD | AF_L | 15 | 1.704073744 | 2.38970081 | 0.092812164 | 0 |
| AF_L_FD | AF_L | 16 | 2.011459342 | 2.402856457 | 0.048118735 | 0 |
| AF_L_FD | AF_L | 17 | 2.010156397 | 2.421496988 | 0.048141062 | 0 |
| AF_L_FD | AF_L | 18 | 2.384958629 | 2.352856565 | 0.019468758 | 1 |
| AF_L_FD | AF_L | 19 | 1.725976657 | 2.339365195 | 0.088387148 | 0 |
| AF_L_FD | AF_L | 20 | 1.057265674 | 2.346255012 | 0.293755609 | 0 |
| AF_L_FD | AF_L | 21 | 0.434932138 | 2.395482994 | 0.66478719 | 0 |
| AF_L_FD | AF_L | 22 | 1.676623604 | 2.328999057 | 0.097608028 | 0 |
| AF_L_FD | AF_L | 23 | 1.237498252 | 2.413387693 | 0.219934406 | 0 |
| AF_L_FD | AF_L | 24 | 1.272150156 | 2.400341078 | 0.207726468 | 0 |
| AF_L_FD | AF_L | 25 | 2.592244223 | 2.344707757 | 0.011788248 | 1 |
| AF_L_FD | AF_L | 26 | 3.818360511 | 2.391757182 | 0.000302296 | 1 |
| AF_L_FD | AF_L | 27 | 3.352825158 | 2.392554991 | 0.001272423 | 1 |
| AF_L_FD | AF_L | 28 | 2.672561025 | 2.361553714 | 0.009278529 | 1 |
| AF_L_FD | AF_L | 29 | 2.181386185 | 2.341907416 | 0.033006752 | 0 |
| AF_L_FD | AF_L | 30 | 2.194497927 | 2.326057315 | 0.031622413 | 0 |
| AF_L_FD | AF_L | 31 | 0.879227089 | 2.405656189 | 0.382274849 | 0 |
| AF_L_FD | AF_L | 32 | 0.900393074 | 2.340001737 | 0.371174254 | 0 |
| AF_L_FD | AF_L | 33 | 0.562269043 | 2.359597783 | 0.575568372 | 0 |
| AF_L_FD | AF_L | 34 | 0.430030156 | 2.42819303 | 0.668315631 | 0 |
| AF_L_FD | AF_L | 35 | 1.002836127 | 2.361388098 | 0.31897898 | 0 |
| AF_L_FD | AF_L | 36 | 1.550836274 | 2.352810462 | 0.124858539 | 0 |
| AF_L_FD | AF_L | 37 | 0.543271163 | 2.381499209 | 0.588439074 | 0 |

| | | | | | | |
|---|---|---|---|---|---|---|
| AF_L_FD | AF_L | 38 | -0.109603653 | 2.430935421 | 0.912998603 | 0 |
| AF_L_FD | AF_L | 39 | -0.665664939 | 2.409096094 | 0.507558349 | 0 |
| AF_L_FD | AF_L | 40 | -1.830341505 | 2.391060264 | 0.071000475 | 0 |
| AF_L_FD | AF_L | 41 | -1.09996027 | 2.374500133 | 0.274711677 | 0 |
| AF_L_FD | AF_L | 42 | 0.674384887 | 2.42646598 | 0.502189701 | 0 |
| AF_L_FD | AF_L | 43 | 1.11651797 | 2.43746255 | 0.268029474 | 0 |
| AF_L_FD | AF_L | 44 | -0.193815582 | 2.40374278 | 0.846813435 | 0 |
| AF_L_FD | AF_L | 45 | -1.235516035 | 2.420195994 | 0.220365624 | 0 |
| AF_L_FD | AF_L | 46 | -2.173715651 | 2.369109675 | 0.032651083 | 0 |
| AF_L_FD | AF_L | 47 | -1.980822378 | 2.355626752 | 0.051067557 | 0 |
| AF_L_FD | AF_L | 48 | -1.72695673 | 2.439624625 | 0.088067497 | 0 |
| AF_L_FD | AF_L | 49 | -0.471523937 | 2.378174079 | 0.638534515 | 0 |
| AF_L_FD | AF_L | 50 | -0.920095503 | 2.4389262 | 0.360255461 | 0 |
| AF_R_FD | AF_R | 1 | 1.533761722 | 2.360353397 | 0.131603479 | 0 |
| AF_R_FD | AF_R | 2 | 1.293714051 | 2.393313258 | 0.201944802 | 0 |
| AF_R_FD | AF_R | 3 | 0.62381495 | 2.387470326 | 0.534771537 | 0 |
| AF_R_FD | AF_R | 4 | 0.577921818 | 2.417101793 | 0.565141269 | 0 |
| AF_R_FD | AF_R | 5 | 0.890507051 | 2.405792797 | 0.376418981 | 0 |
| AF_R_FD | AF_R | 6 | 0.257018836 | 2.447415464 | 0.797903845 | 0 |
| AF_R_FD | AF_R | 7 | -1.164965682 | 2.409442761 | 0.247772563 | 0 |
| AF_R_FD | AF_R | 8 | -2.418470757 | 2.435913966 | 0.017828564 | 0 |
| AF_R_FD | AF_R | 9 | -0.587097268 | 2.362757493 | 0.558775829 | 0 |
| AF_R_FD | AF_R | 10 | 1.166138626 | 2.420136031 | 0.247955925 | 0 |
| AF_R_FD | AF_R | 11 | 0.775300512 | 2.441397427 | 0.441136306 | 0 |
| AF_R_FD | AF_R | 12 | -0.20560559 | 2.35226225 | 0.837705448 | 0 |
| AF_R_FD | AF_R | 13 | 0.140890827 | 2.444718608 | 0.888347434 | 0 |
| AF_R_FD | AF_R | 14 | 0.307052584 | 2.389998312 | 0.759754964 | 0 |
| AF_R_FD | AF_R | 15 | 0.457962018 | 2.390264766 | 0.648278825 | 0 |
| AF_R_FD | AF_R | 16 | -0.291451161 | 2.42559358 | 0.771505244 | 0 |
| AF_R_FD | AF_R | 17 | -0.943459201 | 2.408103657 | 0.34859477 | 0 |
| AF_R_FD | AF_R | 18 | -0.998689275 | 2.38288466 | 0.321153395 | 0 |
| AF_R_FD | AF_R | 19 | 0.102995176 | 2.414510149 | 0.918242265 | 0 |
| AF_R_FD | AF_R | 20 | -1.753519964 | 2.425860136 | 0.083597146 | 0 |
| AF_R_FD | AF_R | 21 | -1.016364188 | 2.40135574 | 0.313184966 | 0 |
| AF_R_FD | AF_R | 22 | -0.575025028 | 2.348692114 | 0.566881519 | 0 |
| AF_R_FD | AF_R | 23 | -0.0240758 | 2.356485416 | 0.980856058 | 0 |
| AF_R_FD | AF_R | 24 | 1.260430668 | 2.394522164 | 0.211295946 | 0 |
| AF_R_FD | AF_R | 25 | 1.154472057 | 2.399731588 | 0.251853566 | 0 |
| AF_R_FD | AF_R | 26 | 0.67499065 | 2.416566461 | 0.501767761 | 0 |
| AF_R_FD | AF_R | 27 | 0.797685053 | 2.438798315 | 0.427956936 | 0 |
| AF_R_FD | AF_R | 28 | 1.781398916 | 2.369105707 | 0.079910224 | 0 |
| AF_R_FD | AF_R | 29 | 2.090517424 | 2.408563109 | 0.040461752 | 0 |
| AF_R_FD | AF_R | 30 | 1.661035733 | 2.394734953 | 0.101721823 | 0 |

| | | | | | | |
|---|---|---|---|---|---|---|
| AF_R_FD | AF_R | 31 | 0.77069387 | 2.354301266 | 0.443615935 | 0 |
| AF_R_FD | AF_R | 32 | 0.007386249 | 2.38091505 | 0.994129689 | 0 |
| AF_R_FD | AF_R | 33 | 0.214824375 | 2.37560996 | 0.830635567 | 0 |
| AF_R_FD | AF_R | 34 | 0.745476414 | 2.365130335 | 0.458819733 | 0 |
| AF_R_FD | AF_R | 35 | 0.791634238 | 2.371404744 | 0.431228105 | 0 |
| AF_R_FD | AF_R | 36 | -0.493686837 | 2.409893031 | 0.622933228 | 0 |
| AF_R_FD | AF_R | 37 | -0.453228331 | 2.421894319 | 0.65160253 | 0 |
| AF_R_FD | AF_R | 38 | -0.0566332 | 2.408224104 | 0.954981592 | 0 |
| AF_R_FD | AF_R | 39 | -0.523394231 | 2.43241941 | 0.602227396 | 0 |
| AF_R_FD | AF_R | 40 | -0.318627101 | 2.393541209 | 0.750848456 | 0 |
| AF_R_FD | AF_R | 41 | 0.198013794 | 2.410656547 | 0.84353424 | 0 |
| AF_R_FD | AF_R | 42 | 1.151285997 | 2.363086963 | 0.253229227 | 0 |
| AF_R_FD | AF_R | 43 | 1.286173932 | 2.300233303 | 0.202055192 | 0 |
| AF_R_FD | AF_R | 44 | 0.683319913 | 2.339895753 | 0.496435972 | 0 |
| AF_R_FD | AF_R | 45 | 0.11793832 | 2.406868116 | 0.906430947 | 0 |
| AF_R_FD | AF_R | 46 | -1.261277381 | 2.361249261 | 0.210930841 | 0 |
| AF_R_FD | AF_R | 47 | 0.381164535 | 2.342878674 | 0.704132767 | 0 |
| AF_R_FD | AF_R | 48 | 0.564411189 | 2.365624184 | 0.57443574 | 0 |
| AF_R_FD | AF_R | 49 | -0.788140303 | 2.450312136 | 0.433419968 | 0 |
| AF_R_FD | AF_R | 50 | -0.416026174 | 2.307562896 | 0.678643716 | 0 |
| CC_1_FD | CC_1 | 1 | 0.615573358 | 2.363906395 | 0.539902528 | 0 |
| CC_1_FD | CC_1 | 2 | 0.308170245 | 2.460128298 | 0.758744517 | 0 |
| CC_1_FD | CC_1 | 3 | 0.737960357 | 2.327639331 | 0.462678864 | 0 |
| CC_1_FD | CC_1 | 4 | 0.590840505 | 2.412322273 | 0.556293097 | 0 |
| CC_1_FD | CC_1 | 5 | 0.362837174 | 2.403756988 | 0.717676589 | 0 |
| CC_1_FD | CC_1 | 6 | -0.161175252 | 2.372805864 | 0.872368201 | 0 |
| CC_1_FD | CC_1 | 7 | -0.837309522 | 2.356726782 | 0.405089651 | 0 |
| CC_1_FD | CC_1 | 8 | -0.864827846 | 2.410459206 | 0.389867081 | 0 |
| CC_1_FD | CC_1 | 9 | -0.725269545 | 2.348965215 | 0.470655308 | 0 |
| CC_1_FD | CC_1 | 10 | -0.869000308 | 2.387564702 | 0.387856875 | 0 |
| CC_1_FD | CC_1 | 11 | -0.656492683 | 2.316422155 | 0.513665993 | 0 |
| CC_1_FD | CC_1 | 12 | -0.066447842 | 2.418435302 | 0.947190715 | 0 |
| CC_1_FD | CC_1 | 13 | 0.067871541 | 2.341984641 | 0.946055666 | 0 |
| CC_1_FD | CC_1 | 14 | 0.256750042 | 2.419348289 | 0.798024215 | 0 |
| CC_1_FD | CC_1 | 15 | 1.043339245 | 2.417283448 | 0.29989924 | 0 |
| CC_1_FD | CC_1 | 16 | 1.588806523 | 2.386888923 | 0.116016974 | 0 |
| CC_1_FD | CC_1 | 17 | 1.813234317 | 2.376926292 | 0.073503273 | 0 |
| CC_1_FD | CC_1 | 18 | 1.865878648 | 2.386600724 | 0.065750202 | 0 |
| CC_1_FD | CC_1 | 19 | 1.923799577 | 2.374266131 | 0.058088884 | 0 |
| CC_1_FD | CC_1 | 20 | 1.688173966 | 2.43705188 | 0.095529821 | 0 |
| CC_1_FD | CC_1 | 21 | 1.05552659 | 2.422010553 | 0.29445066 | 0 |
| CC_1_FD | CC_1 | 22 | 0.874111337 | 2.331420003 | 0.384657247 | 0 |
| CC_1_FD | CC_1 | 23 | 0.934685402 | 2.366495299 | 0.352742325 | 0 |

| | | | | | | |
|---|---|---|---|---|---|---|
| CC_1_FD | CC_1 | 24 | 0.87703014 | 2.346176477 | 0.383085518 | 0 |
| CC_1_FD | CC_1 | 25 | 1.083575726 | 2.404363805 | 0.281801117 | 0 |
| CC_1_FD | CC_1 | 26 | 1.12559082 | 2.394745071 | 0.263927183 | 0 |
| CC_1_FD | CC_1 | 27 | 1.334641321 | 2.404368141 | 0.186012776 | 0 |
| CC_1_FD | CC_1 | 28 | 1.623330869 | 2.351766553 | 0.108514229 | 0 |
| CC_1_FD | CC_1 | 29 | 2.128405162 | 2.383467032 | 0.036354836 | 0 |
| CC_1_FD | CC_1 | 30 | 2.171968034 | 2.398050418 | 0.032782584 | 0 |
| CC_1_FD | CC_1 | 31 | 1.824935214 | 2.3633523 | 0.071703361 | 0 |
| CC_1_FD | CC_1 | 32 | 1.777043937 | 2.355309436 | 0.079424109 | 0 |
| CC_1_FD | CC_1 | 33 | 1.40799091 | 2.342952343 | 0.163362888 | 0 |
| CC_1_FD | CC_1 | 34 | 1.190584486 | 2.394687838 | 0.237920791 | 0 |
| CC_1_FD | CC_1 | 35 | 1.169024091 | 2.439322122 | 0.246338039 | 0 |
| CC_1_FD | CC_1 | 36 | 1.117639198 | 2.37046039 | 0.267423671 | 0 |
| CC_1_FD | CC_1 | 37 | 0.897173421 | 2.332572078 | 0.37276835 | 0 |
| CC_1_FD | CC_1 | 38 | 0.569300303 | 2.379055136 | 0.57110151 | 0 |
| CC_1_FD | CC_1 | 39 | 0.566963992 | 2.399237377 | 0.572698806 | 0 |
| CC_1_FD | CC_1 | 40 | 0.002703676 | 2.419287996 | 0.997850075 | 0 |
| CC_1_FD | CC_1 | 41 | 0.049649874 | 2.369147464 | 0.960526672 | 0 |
| CC_1_FD | CC_1 | 42 | -0.340734249 | 2.426650652 | 0.73424476 | 0 |
| CC_1_FD | CC_1 | 43 | -0.438496889 | 2.382910395 | 0.662368774 | 0 |
| CC_1_FD | CC_1 | 44 | -0.648421949 | 2.375319672 | 0.518783017 | 0 |
| CC_1_FD | CC_1 | 45 | -0.766269764 | 2.377140122 | 0.445866234 | 0 |
| CC_1_FD | CC_1 | 46 | -0.299122999 | 2.348725554 | 0.765623531 | 0 |
| CC_1_FD | CC_1 | 47 | 0.41356104 | 2.404339228 | 0.680293873 | 0 |
| CC_1_FD | CC_1 | 48 | 0.388234394 | 2.37001051 | 0.698877843 | 0 |
| CC_1_FD | CC_1 | 49 | 0.2499555 | 2.366876145 | 0.803255785 | 0 |
| CC_1_FD | CC_1 | 50 | 0.010705906 | 2.322904647 | 0.991484427 | 0 |
| CC_2a_FD | CC_2a | 1 | -1.002014194 | 2.364274989 | 0.319426144 | 0 |
| CC_2a_FD | CC_2a | 2 | -0.621153058 | 2.381182474 | 0.536286567 | 0 |
| CC_2a_FD | CC_2a | 3 | -0.263648315 | 2.410834304 | 0.792764695 | 0 |
| CC_2a_FD | CC_2a | 4 | -0.573790793 | 2.408157553 | 0.567931869 | 0 |
| CC_2a_FD | CC_2a | 5 | -0.87390661 | 2.402906085 | 0.38503396 | 0 |
| CC_2a_FD | CC_2a | 6 | -0.670596725 | 2.407620185 | 0.504473151 | 0 |
| CC_2a_FD | CC_2a | 7 | -0.284177943 | 2.400542221 | 0.777008847 | 0 |
| CC_2a_FD | CC_2a | 8 | -0.232047785 | 2.44054959 | 0.817086307 | 0 |
| CC_2a_FD | CC_2a | 9 | -0.486493129 | 2.387401214 | 0.62793131 | 0 |
| CC_2a_FD | CC_2a | 10 | -0.024163607 | 2.389332585 | 0.980785274 | 0 |
| CC_2a_FD | CC_2a | 11 | -0.022790238 | 2.379898163 | 0.981878482 | 0 |
| CC_2a_FD | CC_2a | 12 | -0.683341508 | 2.436939242 | 0.496398958 | 0 |
| CC_2a_FD | CC_2a | 13 | -0.508388231 | 2.346343638 | 0.612670611 | 0 |
| CC_2a_FD | CC_2a | 14 | 0.01639174 | 2.365951949 | 0.9869712 | 0 |
| CC_2a_FD | CC_2a | 15 | 0.584839808 | 2.368493912 | 0.560796576 | 0 |
| CC_2a_FD | CC_2a | 16 | 0.719039397 | 2.358756303 | 0.474775638 | 0 |

| | | | | | | |
|---|---|---|---|---|---|---|
| CC_2a_FD | CC_2a | 17 | 0.873791521 | 2.377192086 | 0.385323803 | 0 |
| CC_2a_FD | CC_2a | 18 | 0.971975364 | 2.360519738 | 0.334314486 | 0 |
| CC_2a_FD | CC_2a | 19 | 1.163870999 | 2.314802281 | 0.248043474 | 0 |
| CC_2a_FD | CC_2a | 20 | 1.351074162 | 2.363746967 | 0.180479802 | 0 |
| CC_2a_FD | CC_2a | 21 | 1.320963995 | 2.378295395 | 0.190308103 | 0 |
| CC_2a_FD | CC_2a | 22 | 0.971785464 | 2.388677676 | 0.334297885 | 0 |
| CC_2a_FD | CC_2a | 23 | 0.662082852 | 2.387831948 | 0.510055103 | 0 |
| CC_2a_FD | CC_2a | 24 | 0.509858356 | 2.399863764 | 0.61167753 | 0 |
| CC_2a_FD | CC_2a | 25 | 0.479397405 | 2.337959126 | 0.632988625 | 0 |
| CC_2a_FD | CC_2a | 26 | 0.909083134 | 2.394250379 | 0.366051408 | 0 |
| CC_2a_FD | CC_2a | 27 | 1.238224815 | 2.349546875 | 0.219217787 | 0 |
| CC_2a_FD | CC_2a | 28 | 1.076738192 | 2.351390408 | 0.284799266 | 0 |
| CC_2a_FD | CC_2a | 29 | 0.964728252 | 2.384042735 | 0.337632891 | 0 |
| CC_2a_FD | CC_2a | 30 | 1.177597257 | 2.402280521 | 0.242575607 | 0 |
| CC_2a_FD | CC_2a | 31 | 1.357175745 | 2.403621659 | 0.17866676 | 0 |
| CC_2a_FD | CC_2a | 32 | 1.307415776 | 2.411699219 | 0.195010305 | 0 |
| CC_2a_FD | CC_2a | 33 | 0.978255675 | 2.329017012 | 0.331119133 | 0 |
| CC_2a_FD | CC_2a | 34 | 0.764615625 | 2.390147955 | 0.447220589 | 0 |
| CC_2a_FD | CC_2a | 35 | 0.888737267 | 2.356744314 | 0.377658907 | 0 |
| CC_2a_FD | CC_2a | 36 | 0.853697252 | 2.432319188 | 0.396368424 | 0 |
| CC_2a_FD | CC_2a | 37 | 0.621023086 | 2.359662503 | 0.536608445 | 0 |
| CC_2a_FD | CC_2a | 38 | 0.179559989 | 2.430659642 | 0.858006523 | 0 |
| CC_2a_FD | CC_2a | 39 | 0.136474727 | 2.395929745 | 0.891830428 | 0 |
| CC_2a_FD | CC_2a | 40 | 0.075730631 | 2.363443449 | 0.939838792 | 0 |
| CC_2a_FD | CC_2a | 41 | 0.240612512 | 2.362857962 | 0.81046411 | 0 |
| CC_2a_FD | CC_2a | 42 | 0.294334315 | 2.420362219 | 0.769255512 | 0 |
| CC_2a_FD | CC_2a | 43 | 0.134714402 | 2.470787164 | 0.893171879 | 0 |
| CC_2a_FD | CC_2a | 44 | 0.405778489 | 2.432340911 | 0.685979278 | 0 |
| CC_2a_FD | CC_2a | 45 | 0.30654777 | 2.400404192 | 0.760002885 | 0 |
| CC_2a_FD | CC_2a | 46 | 0.042095567 | 2.341313395 | 0.966535887 | 0 |
| CC_2a_FD | CC_2a | 47 | -0.48640091 | 2.403187749 | 0.628216076 | 0 |
| CC_2a_FD | CC_2a | 48 | -0.443789527 | 2.414065348 | 0.658559826 | 0 |
| CC_2a_FD | CC_2a | 49 | -0.20527408 | 2.406040701 | 0.837953625 | 0 |
| CC_2a_FD | CC_2a | 50 | -0.287253249 | 2.439398208 | 0.774773291 | 0 |
| CC_2b_FD | CC_2b | 1 | -0.44624442 | 2.41285606 | 0.656761212 | 0 |
| CC_2b_FD | CC_2b | 2 | -0.145090429 | 2.376140966 | 0.885047971 | 0 |
| CC_2b_FD | CC_2b | 3 | -0.901576978 | 2.394388621 | 0.370290765 | 0 |
| CC_2b_FD | CC_2b | 4 | -1.382578021 | 2.378730457 | 0.171056766 | 0 |
| CC_2b_FD | CC_2b | 5 | -1.359750354 | 2.360947557 | 0.178163127 | 0 |
| CC_2b_FD | CC_2b | 6 | -1.453185834 | 2.40200973 | 0.150100818 | 0 |
| CC_2b_FD | CC_2b | 7 | -1.028805526 | 2.374128992 | 0.306642298 | 0 |
| CC_2b_FD | CC_2b | 8 | -0.235063811 | 2.378106608 | 0.814796666 | 0 |
| CC_2b_FD | CC_2b | 9 | 0.415386398 | 2.379497906 | 0.6791566 | 0 |

| | | | | | | |
|---|---|---|---|---|---|---|
| CC_2b_FD | CC_2b | 10 | 0.360703912 | 2.397982429 | 0.719494058 | 0 |
| CC_2b_FD | CC_2b | 11 | -1.037840883 | 2.386201319 | 0.30263639 | 0 |
| CC_2b_FD | CC_2b | 12 | -1.311327989 | 2.443222446 | 0.193734027 | 0 |
| CC_2b_FD | CC_2b | 13 | -1.092975806 | 2.483470991 | 0.277774262 | 0 |
| CC_2b_FD | CC_2b | 14 | -0.682553748 | 2.386195319 | 0.497072888 | 0 |
| CC_2b_FD | CC_2b | 15 | -0.586551268 | 2.383596636 | 0.559514768 | 0 |
| CC_2b_FD | CC_2b | 16 | -0.497011971 | 2.435035277 | 0.620931696 | 0 |
| CC_2b_FD | CC_2b | 17 | -0.244425969 | 2.34984104 | 0.807685614 | 0 |
| CC_2b_FD | CC_2b | 18 | -0.067436788 | 2.382288996 | 0.946445974 | 0 |
| CC_2b_FD | CC_2b | 19 | 0.182388561 | 2.335036697 | 0.855852568 | 0 |
| CC_2b_FD | CC_2b | 20 | 0.480709968 | 2.365390642 | 0.632295354 | 0 |
| CC_2b_FD | CC_2b | 21 | 0.693920097 | 2.304833969 | 0.489986042 | 0 |
| CC_2b_FD | CC_2b | 22 | 0.772972908 | 2.372625639 | 0.442009042 | 0 |
| CC_2b_FD | CC_2b | 23 | 1.182826476 | 2.379351454 | 0.240572399 | 0 |
| CC_2b_FD | CC_2b | 24 | 1.70148084 | 2.328934817 | 0.092769909 | 0 |
| CC_2b_FD | CC_2b | 25 | 1.787548913 | 2.31209992 | 0.077610912 | 0 |
| CC_2b_FD | CC_2b | 26 | 0.677145303 | 2.34146247 | 0.500246312 | 0 |
| CC_2b_FD | CC_2b | 27 | -0.380725095 | 2.369383366 | 0.704409856 | 0 |
| CC_2b_FD | CC_2b | 28 | -0.679448199 | 2.391588255 | 0.498886218 | 0 |
| CC_2b_FD | CC_2b | 29 | -0.136177876 | 2.344720153 | 0.892059759 | 0 |
| CC_2b_FD | CC_2b | 30 | 0.577786127 | 2.423057979 | 0.56529945 | 0 |
| CC_2b_FD | CC_2b | 31 | 1.388327592 | 2.438031187 | 0.169363111 | 0 |
| CC_2b_FD | CC_2b | 32 | 2.272502886 | 2.363704848 | 0.026033489 | 0 |
| CC_2b_FD | CC_2b | 33 | 2.875363353 | 2.382500453 | 0.005275068 | 1 |
| CC_2b_FD | CC_2b | 34 | 2.927689646 | 2.32996521 | 0.004557399 | 1 |
| CC_2b_FD | CC_2b | 35 | 2.945972975 | 2.385453444 | 0.004340208 | 1 |
| CC_2b_FD | CC_2b | 36 | 3.063940491 | 2.453279241 | 0.003071488 | 1 |
| CC_2b_FD | CC_2b | 37 | 2.755808403 | 2.408121144 | 0.00742276 | 1 |
| CC_2b_FD | CC_2b | 38 | 2.299959155 | 2.432981122 | 0.024548303 | 0 |
| CC_2b_FD | CC_2b | 39 | 1.677093129 | 2.376861693 | 0.098442591 | 0 |
| CC_2b_FD | CC_2b | 40 | 1.019251268 | 2.410179353 | 0.311830249 | 0 |
| CC_2b_FD | CC_2b | 41 | 0.43085414 | 2.371207924 | 0.667998967 | 0 |
| CC_2b_FD | CC_2b | 42 | -0.409128575 | 2.397727022 | 0.683900348 | 0 |
| CC_2b_FD | CC_2b | 43 | -0.179929159 | 2.41391638 | 0.857751371 | 0 |
| CC_2b_FD | CC_2b | 44 | 0.019504121 | 2.420934915 | 0.984491869 | 0 |
| CC_2b_FD | CC_2b | 45 | -0.283878837 | 2.422619466 | 0.777227821 | 0 |
| CC_2b_FD | CC_2b | 46 | -0.795778905 | 2.3878288 | 0.428504584 | 0 |
| CC_2b_FD | CC_2b | 47 | -0.580642364 | 2.393833607 | 0.563101241 | 0 |
| CC_2b_FD | CC_2b | 48 | -0.40171356 | 2.347089958 | 0.689022508 | 0 |
| CC_2b_FD | CC_2b | 49 | -0.402798265 | 2.428837298 | 0.688336757 | 0 |
| CC_2b_FD | CC_2b | 50 | -0.286615762 | 2.365599685 | 0.775230643 | 0 |
| CC_3_FD | CC_3 | 1 | -0.839890042 | 2.346016536 | 0.403571402 | 0 |
| CC_3_FD | CC_3 | 2 | -1.166414222 | 2.317700718 | 0.246870312 | 0 |

| | | | | | | |
|---|---|---|---|---|---|---|
| CC_3_FD | CC_3 | 3 | -0.936361365 | 2.423073187 | 0.351900001 | 0 |
| CC_3_FD | CC_3 | 4 | -0.127793078 | 2.351444736 | 0.898639885 | 0 |
| CC_3_FD | CC_3 | 5 | 0.242961764 | 2.380151918 | 0.808683902 | 0 |
| CC_3_FD | CC_3 | 6 | 0.434955157 | 2.378896607 | 0.664809496 | 0 |
| CC_3_FD | CC_3 | 7 | 0.348668882 | 2.355933562 | 0.728328814 | 0 |
| CC_3_FD | CC_3 | 8 | 0.427421185 | 2.391775345 | 0.670302034 | 0 |
| CC_3_FD | CC_3 | 9 | -0.294611324 | 2.403333718 | 0.769076322 | 0 |
| CC_3_FD | CC_3 | 10 | -0.234684984 | 2.392519719 | 0.815107816 | 0 |
| CC_3_FD | CC_3 | 11 | 0.134766564 | 2.413598112 | 0.893197445 | 0 |
| CC_3_FD | CC_3 | 12 | 0.103190851 | 2.44150394 | 0.918098703 | 0 |
| CC_3_FD | CC_3 | 13 | -0.302306069 | 2.38461831 | 0.763320675 | 0 |
| CC_3_FD | CC_3 | 14 | -0.304278661 | 2.367928717 | 0.76184756 | 0 |
| CC_3_FD | CC_3 | 15 | -0.394512071 | 2.394759688 | 0.694469805 | 0 |
| CC_3_FD | CC_3 | 16 | -0.21056536 | 2.334834482 | 0.833897198 | 0 |
| CC_3_FD | CC_3 | 17 | 0.239372549 | 2.3776016 | 0.811599486 | 0 |
| CC_3_FD | CC_3 | 18 | 0.627301805 | 2.383390035 | 0.532748613 | 0 |
| CC_3_FD | CC_3 | 19 | 0.928955878 | 2.391501166 | 0.356333177 | 0 |
| CC_3_FD | CC_3 | 20 | 1.161079709 | 2.405385175 | 0.249487015 | 0 |
| CC_3_FD | CC_3 | 21 | 1.612057485 | 2.359810623 | 0.11111245 | 0 |
| CC_3_FD | CC_3 | 22 | 1.887602956 | 2.304490357 | 0.062756942 | 0 |
| CC_3_FD | CC_3 | 23 | 1.832401553 | 2.370372801 | 0.070609832 | 0 |
| CC_3_FD | CC_3 | 24 | 1.877411393 | 2.367239794 | 0.064138134 | 0 |
| CC_3_FD | CC_3 | 25 | 1.785255713 | 2.44499138 | 0.078174241 | 0 |
| CC_3_FD | CC_3 | 26 | 0.933244674 | 2.423437071 | 0.35377545 | 0 |
| CC_3_FD | CC_3 | 27 | 0.269509158 | 2.324533094 | 0.788379656 | 0 |
| CC_3_FD | CC_3 | 28 | 0.410805608 | 2.381102826 | 0.682578091 | 0 |
| CC_3_FD | CC_3 | 29 | 0.962511409 | 2.402826475 | 0.339002468 | 0 |
| CC_3_FD | CC_3 | 30 | 1.459368061 | 2.417038011 | 0.148527035 | 0 |
| CC_3_FD | CC_3 | 31 | 2.041714126 | 2.325046033 | 0.044503798 | 0 |
| CC_3_FD | CC_3 | 32 | 2.376170619 | 2.330367388 | 0.01988871 | 1 |
| CC_3_FD | CC_3 | 33 | 2.388770335 | 2.296751429 | 0.019363748 | 1 |
| CC_3_FD | CC_3 | 34 | 1.819047974 | 2.346708318 | 0.073147135 | 0 |
| CC_3_FD | CC_3 | 35 | 1.328138242 | 2.343116084 | 0.188650165 | 0 |
| CC_3_FD | CC_3 | 36 | 0.772927498 | 2.369356739 | 0.442355307 | 0 |
| CC_3_FD | CC_3 | 37 | 0.377944995 | 2.453572824 | 0.706693496 | 0 |
| CC_3_FD | CC_3 | 38 | 0.344384333 | 2.372876013 | 0.731637915 | 0 |
| CC_3_FD | CC_3 | 39 | 0.412456999 | 2.405504935 | 0.681368759 | 0 |
| CC_3_FD | CC_3 | 40 | 0.955457854 | 2.395985269 | 0.343049036 | 0 |
| CC_3_FD | CC_3 | 41 | 1.737241829 | 2.416661988 | 0.0868588 | 0 |
| CC_3_FD | CC_3 | 42 | 1.668023087 | 2.377923051 | 0.099640668 | 0 |
| CC_3_FD | CC_3 | 43 | 0.372621361 | 2.411346711 | 0.710470635 | 0 |
| CC_3_FD | CC_3 | 44 | -0.692506191 | 2.373078756 | 0.490640812 | 0 |
| CC_3_FD | CC_3 | 45 | -1.234820125 | 2.411855385 | 0.220512807 | 0 |

| | | | | | | |
|---|---|---|---|---|---|---|
| CC_3_FD | CC_3 | 46 | -1.255639351 | 2.401451881 | 0.213286412 | 0 |
| CC_3_FD | CC_3 | 47 | -0.88296657 | 2.387714245 | 0.380951906 | 0 |
| CC_3_FD | CC_3 | 48 | -0.193927119 | 2.411125422 | 0.846935026 | 0 |
| CC_3_FD | CC_3 | 49 | 0.800339097 | 2.407624393 | 0.426658947 | 0 |
| CC_3_FD | CC_3 | 50 | 0.643510544 | 2.41446412 | 0.52207213 | 0 |
| CC_4_FD | CC_4 | 1 | 1.095501563 | 2.402234151 | 0.277292684 | 0 |
| CC_4_FD | CC_4 | 2 | 0.131328777 | 2.399893528 | 0.895967409 | 0 |
| CC_4_FD | CC_4 | 3 | -0.366307112 | 2.38091127 | 0.715369884 | 0 |
| CC_4_FD | CC_4 | 4 | -0.336470165 | 2.350903513 | 0.737527489 | 0 |
| CC_4_FD | CC_4 | 5 | 0.353901578 | 2.318749351 | 0.724450515 | 0 |
| CC_4_FD | CC_4 | 6 | 0.912306194 | 2.402405774 | 0.364906183 | 0 |
| CC_4_FD | CC_4 | 7 | 1.199096409 | 2.373795432 | 0.234876214 | 0 |
| CC_4_FD | CC_4 | 8 | 1.803891988 | 2.400678224 | 0.07580297 | 0 |
| CC_4_FD | CC_4 | 9 | 2.170102656 | 2.428054398 | 0.033305229 | 0 |
| CC_4_FD | CC_4 | 10 | 1.370103282 | 2.416187485 | 0.17460647 | 0 |
| CC_4_FD | CC_4 | 11 | 0.787327651 | 2.370158384 | 0.43343208 | 0 |
| CC_4_FD | CC_4 | 12 | 0.25090984 | 2.382455941 | 0.802787468 | 0 |
| CC_4_FD | CC_4 | 13 | 0.140620039 | 2.435596564 | 0.888691559 | 0 |
| CC_4_FD | CC_4 | 14 | -0.117915672 | 2.404604825 | 0.906548471 | 0 |
| CC_4_FD | CC_4 | 15 | -0.222332193 | 2.376672721 | 0.824801318 | 0 |
| CC_4_FD | CC_4 | 16 | 0.177273094 | 2.360394124 | 0.859855243 | 0 |
| CC_4_FD | CC_4 | 17 | 0.812373516 | 2.451624551 | 0.419617004 | 0 |
| CC_4_FD | CC_4 | 18 | 1.276935118 | 2.327480802 | 0.205975149 | 0 |
| CC_4_FD | CC_4 | 19 | 1.847894814 | 2.465750132 | 0.06857748 | 0 |
| CC_4_FD | CC_4 | 20 | 2.307807487 | 2.388689086 | 0.023621375 | 0 |
| CC_4_FD | CC_4 | 21 | 1.796812124 | 2.363449942 | 0.076150665 | 0 |
| CC_4_FD | CC_4 | 22 | 1.210213703 | 2.409775727 | 0.229800953 | 0 |
| CC_4_FD | CC_4 | 23 | 0.893480422 | 2.332085847 | 0.37428051 | 0 |
| CC_4_FD | CC_4 | 24 | 0.619586026 | 2.362277556 | 0.53729968 | 0 |
| CC_4_FD | CC_4 | 25 | 0.514071161 | 2.441955387 | 0.608635428 | 0 |
| CC_4_FD | CC_4 | 26 | 0.642524352 | 2.376934787 | 0.522368455 | 0 |
| CC_4_FD | CC_4 | 27 | 0.529531197 | 2.366072491 | 0.597919163 | 0 |
| CC_4_FD | CC_4 | 28 | 0.366446749 | 2.407523011 | 0.715039066 | 0 |
| CC_4_FD | CC_4 | 29 | 0.578537616 | 2.405730446 | 0.564680921 | 0 |
| CC_4_FD | CC_4 | 30 | 0.849225358 | 2.385285365 | 0.398518618 | 0 |
| CC_4_FD | CC_4 | 31 | 1.269532779 | 2.43325544 | 0.208303618 | 0 |
| CC_4_FD | CC_4 | 32 | 1.637228295 | 2.405363901 | 0.106145429 | 0 |
| CC_4_FD | CC_4 | 33 | 1.719598019 | 2.398593196 | 0.089828491 | 0 |
| CC_4_FD | CC_4 | 34 | 1.829859909 | 2.327126403 | 0.071390766 | 0 |
| CC_4_FD | CC_4 | 35 | 1.64516886 | 2.349037214 | 0.104374702 | 0 |
| CC_4_FD | CC_4 | 36 | 1.17674335 | 2.45096901 | 0.24334824 | 0 |
| CC_4_FD | CC_4 | 37 | 0.618002229 | 2.350142055 | 0.538561924 | 0 |
| CC_4_FD | CC_4 | 38 | 0.354511422 | 2.360121227 | 0.724016401 | 0 |

| | | | | | | |
|---|---|---|---|---|---|---|
| CC_4_FD | CC_4 | 39 | 0.173123671 | 2.344968228 | 0.863043303 | 0 |
| CC_4_FD | CC_4 | 40 | -0.290649578 | 2.379379504 | 0.772121644 | 0 |
| CC_4_FD | CC_4 | 41 | -0.748411592 | 2.424144451 | 0.456653755 | 0 |
| CC_4_FD | CC_4 | 42 | -0.795420414 | 2.373763899 | 0.428927567 | 0 |
| CC_4_FD | CC_4 | 43 | -0.697755022 | 2.425720149 | 0.487932464 | 0 |
| CC_4_FD | CC_4 | 44 | -0.818774518 | 2.395286396 | 0.415974168 | 0 |
| CC_4_FD | CC_4 | 45 | -0.905231184 | 2.378057308 | 0.368771057 | 0 |
| CC_4_FD | CC_4 | 46 | -0.719593856 | 2.378374187 | 0.474454402 | 0 |
| CC_4_FD | CC_4 | 47 | 0.037890128 | 2.395958051 | 0.96989506 | 0 |
| CC_4_FD | CC_4 | 48 | 0.989839227 | 2.380579122 | 0.325985213 | 0 |
| CC_4_FD | CC_4 | 49 | 1.252652459 | 2.387864038 | 0.214551217 | 0 |
| CC_4_FD | CC_4 | 50 | 0.354252195 | 2.381073976 | 0.724128145 | 0 |
| CC_5_FD | CC_5 | 1 | 1.514366235 | 2.403923051 | 0.134454109 | 0 |
| CC_5_FD | CC_5 | 2 | 2.196950858 | 2.350090786 | 0.031748389 | 0 |
| CC_5_FD | CC_5 | 3 | 1.907189908 | 2.410606825 | 0.061504117 | 0 |
| CC_5_FD | CC_5 | 4 | 1.237806196 | 2.383609611 | 0.220998403 | 0 |
| CC_5_FD | CC_5 | 5 | 1.060216526 | 2.374547204 | 0.293357755 | 0 |
| CC_5_FD | CC_5 | 6 | 0.924269653 | 2.391930803 | 0.358608094 | 0 |
| CC_5_FD | CC_5 | 7 | 0.613491275 | 2.3713788 | 0.541548045 | 0 |
| CC_5_FD | CC_5 | 8 | 0.201903534 | 2.3692536 | 0.840568196 | 0 |
| CC_5_FD | CC_5 | 9 | -0.110847261 | 2.436638887 | 0.912053796 | 0 |
| CC_5_FD | CC_5 | 10 | -0.159213225 | 2.414326084 | 0.873934013 | 0 |
| CC_5_FD | CC_5 | 11 | -0.410052936 | 2.410123103 | 0.682898718 | 0 |
| CC_5_FD | CC_5 | 12 | -0.629831551 | 2.375956099 | 0.530716488 | 0 |
| CC_5_FD | CC_5 | 13 | -0.558576295 | 2.371598807 | 0.578333353 | 0 |
| CC_5_FD | CC_5 | 14 | -0.294968574 | 2.417171827 | 0.768973044 | 0 |
| CC_5_FD | CC_5 | 15 | 0.083725137 | 2.378748101 | 0.933538687 | 0 |
| CC_5_FD | CC_5 | 16 | 0.118258887 | 2.361407308 | 0.906231823 | 0 |
| CC_5_FD | CC_5 | 17 | 0.796534161 | 2.42211072 | 0.428360992 | 0 |
| CC_5_FD | CC_5 | 18 | 1.288946789 | 2.353390452 | 0.201345046 | 0 |
| CC_5_FD | CC_5 | 19 | 1.644878845 | 2.322193833 | 0.10389976 | 0 |
| CC_5_FD | CC_5 | 20 | 1.856080474 | 2.366551415 | 0.067084058 | 0 |
| CC_5_FD | CC_5 | 21 | 1.954692265 | 2.364614705 | 0.054119566 | 0 |
| CC_5_FD | CC_5 | 22 | 1.691375635 | 2.361321712 | 0.094677225 | 0 |
| CC_5_FD | CC_5 | 23 | 1.916604058 | 2.395450123 | 0.059004736 | 0 |
| CC_5_FD | CC_5 | 24 | 1.610999028 | 2.33972719 | 0.111554197 | 0 |
| CC_5_FD | CC_5 | 25 | 1.59069819 | 2.406189335 | 0.115826465 | 0 |
| CC_5_FD | CC_5 | 26 | 1.339768616 | 2.404236546 | 0.184124415 | 0 |
| CC_5_FD | CC_5 | 27 | 0.977074621 | 2.356937253 | 0.331557786 | 0 |
| CC_5_FD | CC_5 | 28 | 0.638084344 | 2.430956859 | 0.525413068 | 0 |
| CC_5_FD | CC_5 | 29 | 0.543070821 | 2.362006122 | 0.588732891 | 0 |
| CC_5_FD | CC_5 | 30 | 0.569777205 | 2.402779064 | 0.57055737 | 0 |
| CC_5_FD | CC_5 | 31 | 0.707399093 | 2.381125653 | 0.481511062 | 0 |

| | | | | | | |
|---|---|---|---|---|---|---|
| CC_5_FD | CC_5 | 32 | 1.207195826 | 2.367475017 | 0.23131072 | 0 |
| CC_5_FD | CC_5 | 33 | 0.876059958 | 2.354602677 | 0.384224469 | 0 |
| CC_5_FD | CC_5 | 34 | 0.676693204 | 2.366080099 | 0.501336963 | 0 |
| CC_5_FD | CC_5 | 35 | 0.394960359 | 2.409960138 | 0.694393593 | 0 |
| CC_5_FD | CC_5 | 36 | 0.4761178 | 2.388887377 | 0.635836827 | 0 |
| CC_5_FD | CC_5 | 37 | 0.785300182 | 2.39302463 | 0.435477758 | 0 |
| CC_5_FD | CC_5 | 38 | 0.828606588 | 2.451868425 | 0.410618016 | 0 |
| CC_5_FD | CC_5 | 39 | 0.777704632 | 2.494897957 | 0.439282347 | 0 |
| CC_5_FD | CC_5 | 40 | 0.09081501 | 2.369275113 | 0.927895947 | 0 |
| CC_5_FD | CC_5 | 41 | -0.564082972 | 2.374270551 | 0.574731641 | 0 |
| CC_5_FD | CC_5 | 42 | -0.758341995 | 2.419047929 | 0.451096978 | 0 |
| CC_5_FD | CC_5 | 43 | -0.324513366 | 2.306379659 | 0.746576224 | 0 |
| CC_5_FD | CC_5 | 44 | 0.195302177 | 2.343611009 | 0.84573911 | 0 |
| CC_5_FD | CC_5 | 45 | 0.307459545 | 2.397036855 | 0.759515671 | 0 |
| CC_5_FD | CC_5 | 46 | 0.33260601 | 2.416859278 | 0.740588882 | 0 |
| CC_5_FD | CC_5 | 47 | -0.126804243 | 2.393988368 | 0.899550505 | 0 |
| CC_5_FD | CC_5 | 48 | -0.820431579 | 2.390706803 | 0.415308426 | 0 |
| CC_5_FD | CC_5 | 49 | -0.62209103 | 2.471662805 | 0.536231934 | 0 |
| CC_5_FD | CC_5 | 50 | -0.00421495 | 2.419363733 | 0.996648288 | 0 |
| CC_6_FD | CC_6 | 1 | 1.693818728 | 2.367308193 | 0.095083606 | 0 |
| CC_6_FD | CC_6 | 2 | 1.061073551 | 2.354065832 | 0.293014697 | 0 |
| CC_6_FD | CC_6 | 3 | 0.838803121 | 2.361334026 | 0.404804257 | 0 |
| CC_6_FD | CC_6 | 4 | 0.598437562 | 2.346871381 | 0.551663058 | 0 |
| CC_6_FD | CC_6 | 5 | 0.557297151 | 2.367305931 | 0.579154067 | 0 |
| CC_6_FD | CC_6 | 6 | 0.260322298 | 2.398484824 | 0.795413464 | 0 |
| CC_6_FD | CC_6 | 7 | 0.601634439 | 2.411787456 | 0.54940156 | 0 |
| CC_6_FD | CC_6 | 8 | 1.298057536 | 2.433785592 | 0.198655442 | 0 |
| CC_6_FD | CC_6 | 9 | 1.378533849 | 2.37119647 | 0.1720655 | 0 |
| CC_6_FD | CC_6 | 10 | 0.947986381 | 2.402720031 | 0.345983049 | 0 |
| CC_6_FD | CC_6 | 11 | 0.332001515 | 2.357231058 | 0.740799897 | 0 |
| CC_6_FD | CC_6 | 12 | 0.149195442 | 2.404104875 | 0.881834276 | 0 |
| CC_6_FD | CC_6 | 13 | 1.077616066 | 2.355554573 | 0.285277092 | 0 |
| CC_6_FD | CC_6 | 14 | 1.106257202 | 2.411703169 | 0.272979884 | 0 |
| CC_6_FD | CC_6 | 15 | 0.271393767 | 2.421073504 | 0.787049011 | 0 |
| CC_6_FD | CC_6 | 16 | -0.207291355 | 2.361979102 | 0.836471311 | 0 |
| CC_6_FD | CC_6 | 17 | -0.089916788 | 2.367009402 | 0.928620639 | 0 |
| CC_6_FD | CC_6 | 18 | 0.443525869 | 2.451364043 | 0.658596341 | 0 |
| CC_6_FD | CC_6 | 19 | 0.85682528 | 2.356426264 | 0.394069498 | 0 |
| CC_6_FD | CC_6 | 20 | 0.446570525 | 2.388779125 | 0.656380931 | 0 |
| CC_6_FD | CC_6 | 21 | 0.22529087 | 2.363792225 | 0.822327922 | 0 |
| CC_6_FD | CC_6 | 22 | 0.604613072 | 2.380481693 | 0.5471362 | 0 |
| CC_6_FD | CC_6 | 23 | 0.358762144 | 2.351736311 | 0.720744723 | 0 |
| CC_6_FD | CC_6 | 24 | 0.262163525 | 2.363394049 | 0.793921399 | 0 |

| | | | | | | |
|---|---|---|---|---|---|---|
| CC_6_FD | CC_6 | 25 | 0.092299208 | 2.375735418 | 0.926694423 | 0 |
| CC_6_FD | CC_6 | 26 | -0.136539041 | 2.363544821 | 0.891738914 | 0 |
| CC_6_FD | CC_6 | 27 | -0.270659482 | 2.388469726 | 0.787411656 | 0 |
| CC_6_FD | CC_6 | 28 | -0.155871426 | 2.379492107 | 0.876528563 | 0 |
| CC_6_FD | CC_6 | 29 | -0.376269581 | 2.402401994 | 0.707700318 | 0 |
| CC_6_FD | CC_6 | 30 | -0.986354824 | 2.394776671 | 0.326903018 | 0 |
| CC_6_FD | CC_6 | 31 | -0.770164924 | 2.401569626 | 0.443451957 | 0 |
| CC_6_FD | CC_6 | 32 | -0.546975451 | 2.415214897 | 0.585924167 | 0 |
| CC_6_FD | CC_6 | 33 | -0.510872896 | 2.386961603 | 0.610975031 | 0 |
| CC_6_FD | CC_6 | 34 | -0.550989537 | 2.364025921 | 0.583455597 | 0 |
| CC_6_FD | CC_6 | 35 | -0.379698643 | 2.362580335 | 0.705404913 | 0 |
| CC_6_FD | CC_6 | 36 | -0.173762625 | 2.383401815 | 0.862592135 | 0 |
| CC_6_FD | CC_6 | 37 | -0.544690098 | 2.371838101 | 0.58781756 | 0 |
| CC_6_FD | CC_6 | 38 | -0.482911125 | 2.346618412 | 0.630708236 | 0 |
| CC_6_FD | CC_6 | 39 | 0.452878021 | 2.404359932 | 0.65205426 | 0 |
| CC_6_FD | CC_6 | 40 | 0.90504956 | 2.401413595 | 0.368379736 | 0 |
| CC_6_FD | CC_6 | 41 | 1.064694149 | 2.374092066 | 0.290530811 | 0 |
| CC_6_FD | CC_6 | 42 | 1.063431888 | 2.362322595 | 0.291454205 | 0 |
| CC_6_FD | CC_6 | 43 | 0.898548125 | 2.407870335 | 0.372037291 | 0 |
| CC_6_FD | CC_6 | 44 | 0.548042475 | 2.437553212 | 0.585408983 | 0 |
| CC_6_FD | CC_6 | 45 | 0.064521481 | 2.362100194 | 0.948739961 | 0 |
| CC_6_FD | CC_6 | 46 | -0.160764913 | 2.370496426 | 0.8727708 | 0 |
| CC_6_FD | CC_6 | 47 | 0.070499753 | 2.393016152 | 0.943992994 | 0 |
| CC_6_FD | CC_6 | 48 | -0.159849558 | 2.377708669 | 0.873401002 | 0 |
| CC_6_FD | CC_6 | 49 | -0.743461796 | 2.389618362 | 0.459372322 | 0 |
| CC_6_FD | CC_6 | 50 | -0.254309818 | 2.393349932 | 0.799977165 | 0 |
| CC_7_FD | CC_7 | 1 | 1.292242371 | 2.401445736 | 0.200636435 | 0 |
| CC_7_FD | CC_7 | 2 | 0.297737373 | 2.383901368 | 0.76690203 | 0 |
| CC_7_FD | CC_7 | 3 | -0.870077986 | 2.400713523 | 0.387760668 | 0 |
| CC_7_FD | CC_7 | 4 | -0.944732423 | 2.37215375 | 0.348105788 | 0 |
| CC_7_FD | CC_7 | 5 | -0.19403258 | 2.368661194 | 0.846760425 | 0 |
| CC_7_FD | CC_7 | 6 | 0.190789888 | 2.381832486 | 0.84920426 | 0 |
| CC_7_FD | CC_7 | 7 | -0.802765818 | 2.348448338 | 0.425092387 | 0 |
| CC_7_FD | CC_7 | 8 | -0.605792878 | 2.423977103 | 0.546824047 | 0 |
| CC_7_FD | CC_7 | 9 | -0.010464548 | 2.393307848 | 0.991687842 | 0 |
| CC_7_FD | CC_7 | 10 | 0.636558083 | 2.408319204 | 0.526405167 | 0 |
| CC_7_FD | CC_7 | 11 | 1.39984535 | 2.427497569 | 0.165882018 | 0 |
| CC_7_FD | CC_7 | 12 | 1.833510689 | 2.369757641 | 0.071066025 | 0 |
| CC_7_FD | CC_7 | 13 | 1.854326664 | 2.387739228 | 0.068517163 | 0 |
| CC_7_FD | CC_7 | 14 | 1.917482647 | 2.36717524 | 0.05988412 | 0 |
| CC_7_FD | CC_7 | 15 | 1.830182403 | 2.420447926 | 0.072085841 | 0 |
| CC_7_FD | CC_7 | 16 | 1.790709655 | 2.421728644 | 0.078265718 | 0 |
| CC_7_FD | CC_7 | 17 | 1.569753799 | 2.363129328 | 0.1218223 | 0 |

| | | | | | | |
|---|---|---|---|---|---|---|
| CC_7_FD | CC_7 | 18 | 1.395166293 | 2.447723369 | 0.168305041 | 0 |
| CC_7_FD | CC_7 | 19 | 1.022627334 | 2.346131073 | 0.310703297 | 0 |
| CC_7_FD | CC_7 | 20 | 1.06610211 | 2.394012202 | 0.29059959 | 0 |
| CC_7_FD | CC_7 | 21 | 1.164005077 | 2.351932448 | 0.248370347 | 0 |
| CC_7_FD | CC_7 | 22 | 1.373076258 | 2.413138592 | 0.173975983 | 0 |
| CC_7_FD | CC_7 | 23 | 1.552434313 | 2.400342955 | 0.125200701 | 0 |
| CC_7_FD | CC_7 | 24 | 1.388978593 | 2.378890853 | 0.169515488 | 0 |
| CC_7_FD | CC_7 | 25 | 1.399034144 | 2.364297839 | 0.16664852 | 0 |
| CC_7_FD | CC_7 | 26 | 1.876570714 | 2.4224257 | 0.065093829 | 0 |
| CC_7_FD | CC_7 | 27 | 1.645429018 | 2.400186508 | 0.104939635 | 0 |
| CC_7_FD | CC_7 | 28 | 1.08762428 | 2.41072161 | 0.281004402 | 0 |
| CC_7_FD | CC_7 | 29 | 0.807692266 | 2.35776732 | 0.422232438 | 0 |
| CC_7_FD | CC_7 | 30 | 0.599917094 | 2.357326486 | 0.550492186 | 0 |
| CC_7_FD | CC_7 | 31 | 0.216486095 | 2.392316497 | 0.829208856 | 0 |
| CC_7_FD | CC_7 | 32 | -0.057917244 | 2.402106325 | 0.953976283 | 0 |
| CC_7_FD | CC_7 | 33 | -0.09439843 | 2.295399573 | 0.925067224 | 0 |
| CC_7_FD | CC_7 | 34 | 0.112776141 | 2.322725228 | 0.910540231 | 0 |
| CC_7_FD | CC_7 | 35 | 0.740285391 | 2.435978428 | 0.461570414 | 0 |
| CC_7_FD | CC_7 | 36 | 1.171363277 | 2.360154813 | 0.24518477 | 0 |
| CC_7_FD | CC_7 | 37 | 1.015005296 | 2.410939585 | 0.313402634 | 0 |
| CC_7_FD | CC_7 | 38 | 0.662167877 | 2.391281791 | 0.510026276 | 0 |
| CC_7_FD | CC_7 | 39 | 0.579684206 | 2.420639666 | 0.56399814 | 0 |
| CC_7_FD | CC_7 | 40 | 0.682304658 | 2.418338475 | 0.497288356 | 0 |
| CC_7_FD | CC_7 | 41 | 0.127203832 | 2.417403765 | 0.899123893 | 0 |
| CC_7_FD | CC_7 | 42 | 0.035865642 | 2.367601648 | 0.97148832 | 0 |
| CC_7_FD | CC_7 | 43 | 0.315397718 | 2.378295118 | 0.75351177 | 0 |
| CC_7_FD | CC_7 | 44 | -0.913266434 | 2.403931112 | 0.36459261 | 0 |
| CC_7_FD | CC_7 | 45 | -1.087677304 | 2.466496976 | 0.280404955 | 0 |
| CC_7_FD | CC_7 | 46 | -0.352753992 | 2.382291722 | 0.725210672 | 0 |
| CC_7_FD | CC_7 | 47 | -0.535762349 | 2.402017059 | 0.593619158 | 0 |
| CC_7_FD | CC_7 | 48 | -0.279096792 | 2.379873048 | 0.780955385 | 0 |
| CC_7_FD | CC_7 | 49 | -0.121565033 | 2.424003058 | 0.903619813 | 0 |
| CC_7_FD | CC_7 | 50 | 0.651981635 | 2.317184721 | 0.516809947 | 0 |
| CG_L_FD | CG_L | 1 | -1.838343445 | 2.407333941 | 0.070464972 | 0 |
| CG_L_FD | CG_L | 2 | -0.532289532 | 2.349072783 | 0.596060874 | 0 |
| CG_L_FD | CG_L | 3 | -0.013232945 | 2.365712244 | 0.989479789 | 0 |
| CG_L_FD | CG_L | 4 | 1.858551262 | 2.385367144 | 0.067900262 | 0 |
| CG_L_FD | CG_L | 5 | 0.437753834 | 2.352312246 | 0.662807458 | 0 |
| CG_L_FD | CG_L | 6 | 0.715670299 | 2.346064784 | 0.476619853 | 0 |
| CG_L_FD | CG_L | 7 | 2.259122482 | 2.347151829 | 0.026861974 | 0 |
| CG_L_FD | CG_L | 8 | 2.399902742 | 2.40504835 | 0.01933114 | 0 |
| CG_L_FD | CG_L | 9 | 1.619023136 | 2.413460309 | 0.109790049 | 0 |
| CG_L_FD | CG_L | 10 | 2.285962538 | 2.409607432 | 0.026094348 | 0 |

| | | | | | | |
|---|---|---|---|---|---|---|
| CG_L_FD | CG_L | 11 | 3.067664376 | 2.398273191 | 0.003310323 | 1 |
| CG_L_FD | CG_L | 12 | 2.581357996 | 2.404694963 | 0.012123819 | 1 |
| CG_L_FD | CG_L | 13 | 1.818778626 | 2.408687144 | 0.073710898 | 0 |
| CG_L_FD | CG_L | 14 | 1.917589904 | 2.36391296 | 0.059734445 | 0 |
| CG_L_FD | CG_L | 15 | 2.614614741 | 2.333192423 | 0.011110261 | 1 |
| CG_L_FD | CG_L | 16 | 2.745997866 | 2.340613766 | 0.007585722 | 1 |
| CG_L_FD | CG_L | 17 | 2.507468113 | 2.413806539 | 0.014386942 | 1 |
| CG_L_FD | CG_L | 18 | 2.285349217 | 2.376374832 | 0.025251892 | 0 |
| CG_L_FD | CG_L | 19 | 1.759441812 | 2.366275277 | 0.082682183 | 0 |
| CG_L_FD | CG_L | 20 | 1.640981951 | 2.389482014 | 0.105071507 | 0 |
| CG_L_FD | CG_L | 21 | 2.112953233 | 2.371117875 | 0.038161005 | 0 |
| CG_L_FD | CG_L | 22 | 2.740757302 | 2.374349323 | 0.007860262 | 1 |
| CG_L_FD | CG_L | 23 | 2.388763448 | 2.427467505 | 0.019392113 | 0 |
| CG_L_FD | CG_L | 24 | 1.203125139 | 2.334275977 | 0.232537534 | 0 |
| CG_L_FD | CG_L | 25 | 0.962959925 | 2.353287525 | 0.339013656 | 0 |
| CG_L_FD | CG_L | 26 | 1.37633992 | 2.395580643 | 0.173510804 | 0 |
| CG_L_FD | CG_L | 27 | 1.924023483 | 2.412632547 | 0.058399767 | 0 |
| CG_L_FD | CG_L | 28 | 1.099432313 | 2.422097062 | 0.275133866 | 0 |
| CG_L_FD | CG_L | 29 | 0.757535098 | 2.375102021 | 0.451091775 | 0 |
| CG_L_FD | CG_L | 30 | 0.670750728 | 2.412715258 | 0.504533245 | 0 |
| CG_L_FD | CG_L | 31 | 0.512799108 | 2.306209892 | 0.60966011 | 0 |
| CG_L_FD | CG_L | 32 | 0.460656591 | 2.452237668 | 0.646401317 | 0 |
| CG_L_FD | CG_L | 33 | -0.076758992 | 2.326298019 | 0.939021649 | 0 |
| CG_L_FD | CG_L | 34 | -0.616975432 | 2.335601058 | 0.53929122 | 0 |
| CG_L_FD | CG_L | 35 | -0.351930411 | 2.355421653 | 0.725954012 | 0 |
| CG_L_FD | CG_L | 36 | -0.808753957 | 2.334195644 | 0.421212133 | 0 |
| CG_L_FD | CG_L | 37 | -1.704535902 | 2.368416183 | 0.092665432 | 0 |
| CG_L_FD | CG_L | 38 | -1.583299567 | 2.391410494 | 0.117843449 | 0 |
| CG_L_FD | CG_L | 39 | -0.598727718 | 2.373724479 | 0.551241978 | 0 |
| CG_L_FD | CG_L | 40 | -1.839773079 | 2.41663464 | 0.070473082 | 0 |
| CG_L_FD | CG_L | 41 | -1.743235736 | 2.369077091 | 0.085810292 | 0 |
| CG_L_FD | CG_L | 42 | 0.198579893 | 2.355118889 | 0.843178896 | 0 |
| CG_L_FD | CG_L | 43 | 0.419834154 | 2.356081742 | 0.67613305 | 0 |
| CG_L_FD | CG_L | 44 | -0.729361864 | 2.388007054 | 0.469738875 | 0 |
| CG_L_FD | CG_L | 45 | -2.529501006 | 2.544844411 | 0.022012959 | 0 |
| CG_L_FD | CG_L | 46 | 1.339126949 | 2.406995431 | 0.2228512 | 0 |
| CG_L_FD | CG_L | 47 | NA | NA | NA | NA |
| CG_L_FD | CG_L | 48 | NA | NA | NA | NA |
| CG_L_FD | CG_L | 49 | NA | NA | NA | NA |
| CG_L_FD | CG_L | 50 | NA | NA | NA | NA |
| CG_R_FD | CG_R | 1 | 0.241003455 | 2.391268771 | 0.810360429 | 0 |
| CG_R_FD | CG_R | 2 | -0.799259635 | 2.39821229 | 0.426855781 | 0 |
| CG_R_FD | CG_R | 3 | -0.864976125 | 2.371079969 | 0.389908455 | 0 |

| | | | | | | |
|---|---|---|---|---|---|---|
| CG_R_FD | CG_R | 4 | -0.919668054 | 2.399659534 | 0.360751737 | 0 |
| CG_R_FD | CG_R | 5 | -1.639037943 | 2.408480422 | 0.105142252 | 0 |
| CG_R_FD | CG_R | 6 | -1.869357918 | 2.435790726 | 0.06534041 | 0 |
| CG_R_FD | CG_R | 7 | -0.426388992 | 2.361169394 | 0.670993507 | 0 |
| CG_R_FD | CG_R | 8 | 0.326396005 | 2.391165989 | 0.74506438 | 0 |
| CG_R_FD | CG_R | 9 | 0.289545972 | 2.350118387 | 0.77298589 | 0 |
| CG_R_FD | CG_R | 10 | 0.31442122 | 2.476791713 | 0.754087777 | 0 |
| CG_R_FD | CG_R | 11 | 0.05601692 | 2.381076844 | 0.9554726 | 0 |
| CG_R_FD | CG_R | 12 | 0.905064421 | 2.400438604 | 0.368577221 | 0 |
| CG_R_FD | CG_R | 13 | 1.884427043 | 2.352969055 | 0.063955739 | 0 |
| CG_R_FD | CG_R | 14 | 2.451094679 | 2.355531009 | 0.016940875 | 1 |
| CG_R_FD | CG_R | 15 | 1.172107491 | 2.417556559 | 0.245259034 | 0 |
| CG_R_FD | CG_R | 16 | 0.602381032 | 2.362202741 | 0.5488627 | 0 |
| CG_R_FD | CG_R | 17 | 1.195124814 | 2.381949559 | 0.236354811 | 0 |
| CG_R_FD | CG_R | 18 | 1.706931794 | 2.367193504 | 0.092956546 | 0 |
| CG_R_FD | CG_R | 19 | 1.274291019 | 2.383545295 | 0.207025438 | 0 |
| CG_R_FD | CG_R | 20 | 0.243394113 | 2.333589897 | 0.80839422 | 0 |
| CG_R_FD | CG_R | 21 | 0.264666015 | 2.379604778 | 0.792143924 | 0 |
| CG_R_FD | CG_R | 22 | 0.674022973 | 2.448691204 | 0.503030436 | 0 |
| CG_R_FD | CG_R | 23 | 1.241122975 | 2.388951498 | 0.21932058 | 0 |
| CG_R_FD | CG_R | 24 | 1.205701472 | 2.377912612 | 0.232009686 | 0 |
| CG_R_FD | CG_R | 25 | 1.379174448 | 2.398290294 | 0.171833878 | 0 |
| CG_R_FD | CG_R | 26 | 1.809445507 | 2.375004447 | 0.074412916 | 0 |
| CG_R_FD | CG_R | 27 | 1.153113337 | 2.402781529 | 0.253025301 | 0 |
| CG_R_FD | CG_R | 28 | 0.219294271 | 2.392654772 | 0.827067142 | 0 |
| CG_R_FD | CG_R | 29 | 0.090641 | 2.347169219 | 0.928033504 | 0 |
| CG_R_FD | CG_R | 30 | 0.422925301 | 2.380394051 | 0.673557844 | 0 |
| CG_R_FD | CG_R | 31 | 0.269809508 | 2.444416243 | 0.788010651 | 0 |
| CG_R_FD | CG_R | 32 | 0.051692548 | 2.430613181 | 0.958930152 | 0 |
| CG_R_FD | CG_R | 33 | -0.455170522 | 2.403442703 | 0.650486749 | 0 |
| CG_R_FD | CG_R | 34 | -0.161358458 | 2.385400779 | 0.872290688 | 0 |
| CG_R_FD | CG_R | 35 | 0.907276417 | 2.403776192 | 0.367294678 | 0 |
| CG_R_FD | CG_R | 36 | -0.027364358 | 2.374964883 | 0.978244521 | 0 |
| CG_R_FD | CG_R | 37 | -0.529649392 | 2.343181715 | 0.597881582 | 0 |
| CG_R_FD | CG_R | 38 | -0.421971941 | 2.35958773 | 0.674173837 | 0 |
| CG_R_FD | CG_R | 39 | -0.433982585 | 2.3692731 | 0.665525694 | 0 |
| CG_R_FD | CG_R | 40 | -0.401310162 | 2.370693356 | 0.689372699 | 0 |
| CG_R_FD | CG_R | 41 | 0.652765305 | 2.377833262 | 0.516469481 | 0 |
| CG_R_FD | CG_R | 42 | 0.147850887 | 2.388497894 | 0.883096211 | 0 |
| CG_R_FD | CG_R | 43 | -1.671333766 | 2.377725228 | 0.10227573 | 0 |
| CG_R_FD | CG_R | 44 | 0.830292887 | 2.496944603 | 0.418814499 | 0 |
| CG_R_FD | CG_R | 45 | 1.318064936 | 2.565242095 | 0.220734954 | 0 |
| CG_R_FD | CG_R | 46 | NA | NA | NA | NA |

| | | | | | | |
|---|---|---|---|---|---|---|
| CG_R_FD | CG_R | 47 | NA | NA | NA | NA |
| CR_L_FD | CR_L | 1 | -2.000091253 | 2.373871596 | 0.048986293 | 0 |
| CR_L_FD | CR_L | 2 | -1.303402601 | 2.432815971 | 0.19727599 | 0 |
| CR_L_FD | CR_L | 3 | -0.802592952 | 2.424858279 | 0.424888835 | 0 |
| CR_L_FD | CR_L | 4 | -0.192176964 | 2.371591068 | 0.848145356 | 0 |
| CR_L_FD | CR_L | 5 | 0.457427032 | 2.372409966 | 0.648709005 | 0 |
| CR_L_FD | CR_L | 6 | 0.730710874 | 2.369756488 | 0.467078803 | 0 |
| CR_L_FD | CR_L | 7 | 0.200826205 | 2.417089605 | 0.841385889 | 0 |
| CR_L_FD | CR_L | 8 | 0.356299104 | 2.397505915 | 0.722717048 | 0 |
| CR_L_FD | CR_L | 9 | 0.07642498 | 2.447711701 | 0.939315666 | 0 |
| CR_L_FD | CR_L | 10 | -0.501044774 | 2.362771554 | 0.617851254 | 0 |
| CR_L_FD | CR_L | 11 | -0.113873315 | 2.34620605 | 0.909638745 | 0 |
| CR_L_FD | CR_L | 12 | 0.581416266 | 2.398796117 | 0.562637136 | 0 |
| CR_L_FD | CR_L | 13 | -0.080730531 | 2.434522917 | 0.935857588 | 0 |
| CR_L_FD | CR_L | 14 | -0.920507094 | 2.4635346 | 0.360158376 | 0 |
| CR_L_FD | CR_L | 15 | -0.879292517 | 2.388489797 | 0.382072464 | 0 |
| CR_L_FD | CR_L | 16 | -0.354144407 | 2.336548976 | 0.724206427 | 0 |
| CR_L_FD | CR_L | 17 | -0.125494288 | 2.416271664 | 0.900451456 | 0 |
| CR_L_FD | CR_L | 18 | -0.335689577 | 2.382676437 | 0.737987683 | 0 |
| CR_L_FD | CR_L | 19 | -0.81513626 | 2.382906931 | 0.41755269 | 0 |
| CR_L_FD | CR_L | 20 | -0.816196033 | 2.423608482 | 0.416878212 | 0 |
| CR_L_FD | CR_L | 21 | -0.41129215 | 2.341951232 | 0.681954131 | 0 |
| CR_L_FD | CR_L | 22 | -0.360742689 | 2.401918417 | 0.719259761 | 0 |
| CR_L_FD | CR_L | 23 | 0.035932799 | 2.358304481 | 0.971430629 | 0 |
| CR_L_FD | CR_L | 24 | 0.727663812 | 2.387150753 | 0.469125751 | 0 |
| CR_L_FD | CR_L | 25 | 1.357340609 | 2.409366077 | 0.178622342 | 0 |
| CR_L_FD | CR_L | 26 | 1.877261123 | 2.384309019 | 0.064153577 | 0 |
| CR_L_FD | CR_L | 27 | 1.958155631 | 2.334379251 | 0.05380325 | 0 |
| CR_L_FD | CR_L | 28 | 1.808371203 | 2.430259437 | 0.074759737 | 0 |
| CR_L_FD | CR_L | 29 | 1.931848082 | 2.382546302 | 0.057444207 | 0 |
| CR_L_FD | CR_L | 30 | 2.231855328 | 2.409593146 | 0.0290348 | 0 |
| CR_L_FD | CR_L | 31 | 2.598276342 | 2.401633122 | 0.01138976 | 1 |
| CR_L_FD | CR_L | 32 | 2.477868935 | 2.420673468 | 0.015635206 | 1 |
| CR_L_FD | CR_L | 33 | 1.942660624 | 2.334792075 | 0.05602628 | 0 |
| CR_L_FD | CR_L | 34 | 1.70811507 | 2.374088565 | 0.092068862 | 0 |
| CR_L_FD | CR_L | 35 | 1.711678843 | 2.404404389 | 0.092021754 | 0 |
| CR_L_FD | CR_L | 36 | 1.990425719 | 2.370379699 | 0.05191781 | 0 |
| CR_L_FD | CR_L | 37 | 2.246769513 | 2.417785504 | 0.029409852 | 0 |
| CR_L_FD | CR_L | 38 | 2.385252487 | 2.435878157 | 0.021275105 | 0 |
| CR_L_FD | CR_L | 39 | 2.557892799 | 2.454375816 | 0.013896386 | 1 |
| CR_L_FD | CR_L | 40 | 2.086286955 | 2.395834678 | 0.041916916 | 0 |
| CR_L_FD | CR_L | 41 | 1.152518761 | 2.407456203 | 0.253491072 | 0 |
| CR_L_FD | CR_L | 42 | 0.359362562 | 2.349598164 | 0.720464189 | 0 |

| | | | | | | |
|---|---|---|---|---|---|---|
| CR_L_FD | CR_L | 43 | 0.140314765 | 2.379094378 | 0.888868426 | 0 |
| CR_L_FD | CR_L | 44 | -0.037634378 | 2.38407356 | 0.970085773 | 0 |
| CR_L_FD | CR_L | 45 | 0.281410805 | 2.411226302 | 0.779156144 | 0 |
| CR_L_FD | CR_L | 46 | 0.972166087 | 2.404766714 | 0.334217727 | 0 |
| CR_L_FD | CR_L | 47 | 1.230826715 | 2.39762982 | 0.222259685 | 0 |
| CR_L_FD | CR_L | 48 | 1.738715925 | 2.370517441 | 0.085942033 | 0 |
| CR_L_FD | CR_L | 49 | 1.930249561 | 2.3849961 | 0.057187607 | 0 |
| CR_L_FD | CR_L | 50 | 1.598134857 | 2.3934869 | 0.114474873 | 0 |
| CR_R_FD | CR_R | 1 | -0.728645375 | 2.429650253 | 0.468365419 | 0 |
| CR_R_FD | CR_R | 2 | -1.563207255 | 2.36264797 | 0.122026253 | 0 |
| CR_R_FD | CR_R | 3 | -1.191446464 | 2.382580653 | 0.237561344 | 0 |
| CR_R_FD | CR_R | 4 | -0.785817843 | 2.399511373 | 0.434847725 | 0 |
| CR_R_FD | CR_R | 5 | -0.908690595 | 2.386238118 | 0.366851001 | 0 |
| CR_R_FD | CR_R | 6 | -0.835539021 | 2.44365071 | 0.406277595 | 0 |
| CR_R_FD | CR_R | 7 | -0.656787325 | 2.38757578 | 0.513418593 | 0 |
| CR_R_FD | CR_R | 8 | -0.71462172 | 2.374220055 | 0.47719056 | 0 |
| CR_R_FD | CR_R | 9 | -1.099771065 | 2.380103941 | 0.275004623 | 0 |
| CR_R_FD | CR_R | 10 | -0.700566547 | 2.473121708 | 0.485829304 | 0 |
| CR_R_FD | CR_R | 11 | -0.435964308 | 2.394063358 | 0.664355169 | 0 |
| CR_R_FD | CR_R | 12 | 0.043317996 | 2.432214013 | 0.965572144 | 0 |
| CR_R_FD | CR_R | 13 | 0.405938231 | 2.44840441 | 0.6859633 | 0 |
| CR_R_FD | CR_R | 14 | -0.542304758 | 2.405499436 | 0.58914309 | 0 |
| CR_R_FD | CR_R | 15 | -0.896362069 | 2.40580856 | 0.372720351 | 0 |
| CR_R_FD | CR_R | 16 | -0.881392566 | 2.41192422 | 0.380935914 | 0 |
| CR_R_FD | CR_R | 17 | -0.831294655 | 2.340485916 | 0.408571581 | 0 |
| CR_R_FD | CR_R | 18 | -0.630980051 | 2.363205705 | 0.529926486 | 0 |
| CR_R_FD | CR_R | 19 | -0.363616467 | 2.439945955 | 0.717135916 | 0 |
| CR_R_FD | CR_R | 20 | -0.562650294 | 2.370054196 | 0.575397575 | 0 |
| CR_R_FD | CR_R | 21 | -0.94137496 | 2.382462972 | 0.349835867 | 0 |
| CR_R_FD | CR_R | 22 | -0.537466185 | 2.354632983 | 0.592686083 | 0 |
| CR_R_FD | CR_R | 23 | -0.090490007 | 2.337671198 | 0.928140833 | 0 |
| CR_R_FD | CR_R | 24 | 0.328242168 | 2.346211504 | 0.743630631 | 0 |
| CR_R_FD | CR_R | 25 | 0.279710066 | 2.365872082 | 0.780488498 | 0 |
| CR_R_FD | CR_R | 26 | 0.41140178 | 2.339307787 | 0.68191983 | 0 |
| CR_R_FD | CR_R | 27 | 0.799496994 | 2.349954724 | 0.42637388 | 0 |
| CR_R_FD | CR_R | 28 | 1.287202179 | 2.387713048 | 0.201809933 | 0 |
| CR_R_FD | CR_R | 29 | 1.64850077 | 2.363952061 | 0.103415491 | 0 |
| CR_R_FD | CR_R | 30 | 1.865014292 | 2.392455426 | 0.066393498 | 0 |
| CR_R_FD | CR_R | 31 | 1.946048043 | 2.334393634 | 0.055904425 | 0 |
| CR_R_FD | CR_R | 32 | 2.03098192 | 2.413517121 | 0.046649013 | 0 |
| CR_R_FD | CR_R | 33 | 2.041787709 | 2.423978258 | 0.045795701 | 0 |
| CR_R_FD | CR_R | 34 | 2.270498044 | 2.332569483 | 0.027084479 | 0 |
| CR_R_FD | CR_R | 35 | 2.315884643 | 2.431654924 | 0.024573649 | 0 |

| | | | | | | |
|---|---|---|---|---|---|---|
| CR_R_FD | CR_R | 36 | 2.057503164 | 2.478525052 | 0.044583314 | 0 |
| CR_R_FD | CR_R | 37 | 2.181590765 | 2.404879021 | 0.033605274 | 0 |
| CR_R_FD | CR_R | 38 | 1.807086039 | 2.404440873 | 0.076884541 | 0 |
| CR_R_FD | CR_R | 39 | 1.292195456 | 2.405220769 | 0.202067958 | 0 |
| CR_R_FD | CR_R | 40 | 1.598188717 | 2.342841904 | 0.115759994 | 0 |
| CR_R_FD | CR_R | 41 | 1.660413375 | 2.359447352 | 0.10217809 | 0 |
| CR_R_FD | CR_R | 42 | 1.050640842 | 2.44120733 | 0.297643299 | 0 |
| CR_R_FD | CR_R | 43 | 0.846644185 | 2.390440057 | 0.40050331 | 0 |
| CR_R_FD | CR_R | 44 | 0.760849525 | 2.416550332 | 0.449623894 | 0 |
| CR_R_FD | CR_R | 45 | 0.630005952 | 2.369241643 | 0.530921986 | 0 |
| CR_R_FD | CR_R | 46 | 0.453369878 | 2.345644334 | 0.651735711 | 0 |
| CR_R_FD | CR_R | 47 | -0.313760326 | 2.358598642 | 0.754564176 | 0 |
| CR_R_FD | CR_R | 48 | -0.47196784 | 2.427911664 | 0.638258532 | 0 |
| CR_R_FD | CR_R | 49 | 0.142827589 | 2.382353493 | 0.886795838 | 0 |
| CR_R_FD | CR_R | 50 | 0.746806855 | 2.399574429 | 0.457379813 | 0 |
| CST_L_FD | CST_L | 1 | -1.163429691 | 2.373045181 | 0.248295428 | 0 |
| CST_L_FD | CST_L | 2 | 0.149173757 | 2.396958146 | 0.881858337 | 0 |
| CST_L_FD | CST_L | 3 | -0.692354857 | 2.376614233 | 0.491029393 | 0 |
| CST_L_FD | CST_L | 4 | -0.974146088 | 2.395914144 | 0.333585995 | 0 |
| CST_L_FD | CST_L | 5 | -0.833544326 | 2.384747439 | 0.407599601 | 0 |
| CST_L_FD | CST_L | 6 | -0.049886873 | 2.418650787 | 0.960344746 | 0 |
| CST_L_FD | CST_L | 7 | 0.446737236 | 2.360432822 | 0.656341865 | 0 |
| CST_L_FD | CST_L | 8 | 0.090864008 | 2.414436417 | 0.927835614 | 0 |
| CST_L_FD | CST_L | 9 | -0.194553917 | 2.417134974 | 0.846259182 | 0 |
| CST_L_FD | CST_L | 10 | -0.641916667 | 2.391892397 | 0.522816473 | 0 |
| CST_L_FD | CST_L | 11 | -0.502955305 | 2.415133994 | 0.616418463 | 0 |
| CST_L_FD | CST_L | 12 | -0.034497075 | 2.427725999 | 0.972567255 | 0 |
| CST_L_FD | CST_L | 13 | 0.85490425 | 2.376342567 | 0.395239296 | 0 |
| CST_L_FD | CST_L | 14 | 0.910150873 | 2.385482783 | 0.365596589 | 0 |
| CST_L_FD | CST_L | 15 | 0.324081647 | 2.323803869 | 0.746788505 | 0 |
| CST_L_FD | CST_L | 16 | -0.017868505 | 2.384183553 | 0.985790112 | 0 |
| CST_L_FD | CST_L | 17 | 0.032633977 | 2.3943344 | 0.974048905 | 0 |
| CST_L_FD | CST_L | 18 | -0.036661375 | 2.391845349 | 0.970850783 | 0 |
| CST_L_FD | CST_L | 19 | -0.242855414 | 2.411296459 | 0.808787358 | 0 |
| CST_L_FD | CST_L | 20 | -0.506056769 | 2.437932227 | 0.614319331 | 0 |
| CST_L_FD | CST_L | 21 | -0.458455266 | 2.401796432 | 0.647971544 | 0 |
| CST_L_FD | CST_L | 22 | 0.169572489 | 2.398349296 | 0.865791031 | 0 |
| CST_L_FD | CST_L | 23 | 0.257559271 | 2.416837503 | 0.797427995 | 0 |
| CST_L_FD | CST_L | 24 | 0.781631087 | 2.410651712 | 0.436794769 | 0 |
| CST_L_FD | CST_L | 25 | 1.223066917 | 2.369388462 | 0.224918053 | 0 |
| CST_L_FD | CST_L | 26 | 1.459976798 | 2.363111635 | 0.148211372 | 0 |
| CST_L_FD | CST_L | 27 | 1.682822285 | 2.388925145 | 0.096519548 | 0 |
| CST_L_FD | CST_L | 28 | 2.249449291 | 2.399075476 | 0.027431204 | 0 |

| | | | | | | |
|---|---|---|---|---|---|---|
| CST_L_FD | CST_L | 29 | 2.287073412 | 2.407356672 | 0.02495586 | 0 |
| CST_L_FD | CST_L | 30 | 2.476967728 | 2.438335795 | 0.015465625 | 1 |
| CST_L_FD | CST_L | 31 | 2.515944126 | 2.40747484 | 0.014146237 | 1 |
| CST_L_FD | CST_L | 32 | 2.650057836 | 2.393565505 | 0.010214065 | 1 |
| CST_L_FD | CST_L | 33 | 2.870216603 | 2.39662015 | 0.005729749 | 1 |
| CST_L_FD | CST_L | 34 | 2.551839167 | 2.405336269 | 0.013326654 | 1 |
| CST_L_FD | CST_L | 35 | 1.952288025 | 2.504411423 | 0.055535904 | 0 |
| CST_L_FD | CST_L | 36 | 1.437761924 | 2.400323834 | 0.155795815 | 0 |
| CST_L_FD | CST_L | 37 | 1.499672646 | 2.392549639 | 0.138774305 | 0 |
| CST_L_FD | CST_L | 38 | 2.291701581 | 2.37149435 | 0.026036645 | 0 |
| CST_L_FD | CST_L | 39 | 1.867541923 | 2.410138474 | 0.067865319 | 0 |
| CST_L_FD | CST_L | 40 | 0.383144994 | 2.362915361 | 0.702903581 | 0 |
| CST_L_FD | CST_L | 41 | 1.512042411 | 2.360772172 | 0.135465667 | 0 |
| CST_L_FD | CST_L | 42 | 2.865362442 | 2.39004447 | 0.005536239 | 1 |
| CST_L_FD | CST_L | 43 | 2.051239248 | 2.429518438 | 0.043697689 | 0 |
| CST_L_FD | CST_L | 44 | 0.65864362 | 2.303436592 | 0.512065492 | 0 |
| CST_L_FD | CST_L | 45 | 1.217373928 | 2.40962145 | 0.227646468 | 0 |
| CST_L_FD | CST_L | 46 | 1.924103908 | 2.398869256 | 0.058843292 | 0 |
| CST_L_FD | CST_L | 47 | 1.92410465 | 2.393453072 | 0.058557854 | 0 |
| CST_L_FD | CST_L | 48 | 1.304915386 | 2.377509742 | 0.19586626 | 0 |
| CST_L_FD | CST_L | 49 | 0.591771996 | 2.361021725 | 0.555747304 | 0 |
| CST_L_FD | CST_L | 50 | -0.387932138 | 2.457759942 | 0.69919026 | 0 |
| CST_R_FD | CST_R | 1 | -0.782489489 | 2.445809602 | 0.436208725 | 0 |
| CST_R_FD | CST_R | 2 | -0.812811177 | 2.403608697 | 0.418723191 | 0 |
| CST_R_FD | CST_R | 3 | -1.458723552 | 2.355834925 | 0.148850009 | 0 |
| CST_R_FD | CST_R | 4 | -0.976047352 | 2.411942959 | 0.332240743 | 0 |
| CST_R_FD | CST_R | 5 | -0.141989494 | 2.387250377 | 0.887481359 | 0 |
| CST_R_FD | CST_R | 6 | 0.423039965 | 2.339607399 | 0.673574791 | 0 |
| CST_R_FD | CST_R | 7 | 0.473188375 | 2.386545033 | 0.637656411 | 0 |
| CST_R_FD | CST_R | 8 | 0.027661888 | 2.422191848 | 0.978012728 | 0 |
| CST_R_FD | CST_R | 9 | -0.246168264 | 2.39724048 | 0.80633931 | 0 |
| CST_R_FD | CST_R | 10 | -0.478843 | 2.38019133 | 0.633569531 | 0 |
| CST_R_FD | CST_R | 11 | -0.550241888 | 2.38887964 | 0.583950026 | 0 |
| CST_R_FD | CST_R | 12 | -0.488350234 | 2.393438643 | 0.626805117 | 0 |
| CST_R_FD | CST_R | 13 | -0.634588551 | 2.384977527 | 0.527603643 | 0 |
| CST_R_FD | CST_R | 14 | -0.98810329 | 2.40791664 | 0.326059171 | 0 |
| CST_R_FD | CST_R | 15 | -1.657252065 | 2.350563882 | 0.101341171 | 0 |
| CST_R_FD | CST_R | 16 | -1.608523743 | 2.403287315 | 0.111688664 | 0 |
| CST_R_FD | CST_R | 17 | -1.392421345 | 2.399771444 | 0.167881164 | 0 |
| CST_R_FD | CST_R | 18 | -1.195278313 | 2.43863024 | 0.23570558 | 0 |
| CST_R_FD | CST_R | 19 | -0.800734041 | 2.349003657 | 0.425906254 | 0 |
| CST_R_FD | CST_R | 20 | -0.853303632 | 2.464453746 | 0.396493823 | 0 |
| CST_R_FD | CST_R | 21 | -1.113883914 | 2.409636844 | 0.269091685 | 0 |

| | | | | | | |
|---|---|---|---|---|---|---|
| CST_R_FD | CST_R | 22 | -1.013198819 | 2.313702338 | 0.314199095 | 0 |
| CST_R_FD | CST_R | 23 | -0.282495877 | 2.359751172 | 0.778309633 | 0 |
| CST_R_FD | CST_R | 24 | 0.324558442 | 2.363863554 | 0.746378788 | 0 |
| CST_R_FD | CST_R | 25 | 0.610967348 | 2.365820433 | 0.543003031 | 0 |
| CST_R_FD | CST_R | 26 | 1.019750881 | 2.351660475 | 0.310927648 | 0 |
| CST_R_FD | CST_R | 27 | 1.247901114 | 2.388187711 | 0.215730067 | 0 |
| CST_R_FD | CST_R | 28 | 1.433231963 | 2.377122748 | 0.155700855 | 0 |
| CST_R_FD | CST_R | 29 | 1.485886191 | 2.39450955 | 0.141560664 | 0 |
| CST_R_FD | CST_R | 30 | 1.541287116 | 2.357294237 | 0.128013717 | 0 |
| CST_R_FD | CST_R | 31 | 1.812092414 | 2.378414548 | 0.075130769 | 0 |
| CST_R_FD | CST_R | 32 | 2.224391463 | 2.426152722 | 0.03031932 | 0 |
| CST_R_FD | CST_R | 33 | 2.142352493 | 2.419893441 | 0.036884742 | 0 |
| CST_R_FD | CST_R | 34 | 1.741413675 | 2.413966515 | 0.087795307 | 0 |
| CST_R_FD | CST_R | 35 | 1.475437802 | 2.397786477 | 0.146699881 | 0 |
| CST_R_FD | CST_R | 36 | 1.156605215 | 2.436992533 | 0.253157631 | 0 |
| CST_R_FD | CST_R | 37 | 1.371527845 | 2.418540534 | 0.175677267 | 0 |
| CST_R_FD | CST_R | 38 | 1.66585867 | 2.365780051 | 0.101875443 | 0 |
| CST_R_FD | CST_R | 39 | 1.8685186 | 2.452700628 | 0.067344073 | 0 |
| CST_R_FD | CST_R | 40 | 1.177424708 | 2.415086697 | 0.243262615 | 0 |
| CST_R_FD | CST_R | 41 | 0.709651793 | 2.405614493 | 0.480718155 | 0 |
| CST_R_FD | CST_R | 42 | 0.390745458 | 2.363383209 | 0.69739753 | 0 |
| CST_R_FD | CST_R | 43 | -0.087607054 | 2.353510878 | 0.930461453 | 0 |
| CST_R_FD | CST_R | 44 | 0.452383628 | 2.395464893 | 0.652338383 | 0 |
| CST_R_FD | CST_R | 45 | 0.737427084 | 2.387363072 | 0.463000683 | 0 |
| CST_R_FD | CST_R | 46 | 0.626581695 | 2.40636538 | 0.532751236 | 0 |
| CST_R_FD | CST_R | 47 | 0.364402486 | 2.39605788 | 0.716723254 | 0 |
| CST_R_FD | CST_R | 48 | -0.477515076 | 2.459464859 | 0.634932392 | 0 |
| CST_R_FD | CST_R | 49 | 0.35524964 | 2.378363178 | 0.723656791 | 0 |
| CST_R_FD | CST_R | 50 | 1.354493319 | 2.41686769 | 0.17984066 | 0 |
| IFOF_L_FD | IFOF_L | 1 | -0.760787791 | 2.355566709 | 0.449477032 | 0 |
| IFOF_L_FD | IFOF_L | 2 | 0.110290882 | 2.420703103 | 0.912508761 | 0 |
| IFOF_L_FD | IFOF_L | 3 | -0.146491917 | 2.405008664 | 0.88395136 | 0 |
| IFOF_L_FD | IFOF_L | 4 | -0.156504004 | 2.443249527 | 0.876100357 | 0 |
| IFOF_L_FD | IFOF_L | 5 | -0.695550732 | 2.373979308 | 0.489069252 | 0 |
| IFOF_L_FD | IFOF_L | 6 | -1.085630167 | 2.398676253 | 0.281541885 | 0 |
| IFOF_L_FD | IFOF_L | 7 | -1.099050263 | 2.362114666 | 0.275639367 | 0 |
| IFOF_L_FD | IFOF_L | 8 | -0.098971707 | 2.388872072 | 0.921448497 | 0 |
| IFOF_L_FD | IFOF_L | 9 | 0.972180774 | 2.422759306 | 0.334387939 | 0 |
| IFOF_L_FD | IFOF_L | 10 | 1.857567289 | 2.369654642 | 0.067392426 | 0 |
| IFOF_L_FD | IFOF_L | 11 | 2.076896588 | 2.344632332 | 0.041510809 | 0 |
| IFOF_L_FD | IFOF_L | 12 | 2.231014559 | 2.334941495 | 0.028755271 | 0 |
| IFOF_L_FD | IFOF_L | 13 | 2.354264051 | 2.358097781 | 0.02129556 | 0 |
| IFOF_L_FD | IFOF_L | 14 | 2.60712761 | 2.374561203 | 0.011163759 | 1 |

| | | | | | | |
|---|---|---|---|---|---|---|
| IFOF_L_FD | IFOF_L | 15 | 2.198319913 | 2.378485542 | 0.031363134 | 0 |
| IFOF_L_FD | IFOF_L | 16 | 1.968339775 | 2.422934586 | 0.052931116 | 0 |
| IFOF_L_FD | IFOF_L | 17 | 1.803553622 | 2.339559511 | 0.075045572 | 0 |
| IFOF_L_FD | IFOF_L | 18 | 1.197939072 | 2.402547497 | 0.234493738 | 0 |
| IFOF_L_FD | IFOF_L | 19 | 0.953048896 | 2.32140604 | 0.343551431 | 0 |
| IFOF_L_FD | IFOF_L | 20 | 1.514128272 | 2.385600819 | 0.134181931 | 0 |
| IFOF_L_FD | IFOF_L | 21 | 2.263515186 | 2.364700404 | 0.026494641 | 0 |
| IFOF_L_FD | IFOF_L | 22 | 2.380076773 | 2.34473009 | 0.020015408 | 1 |
| IFOF_L_FD | IFOF_L | 23 | 1.451463319 | 2.372038953 | 0.150728511 | 0 |
| IFOF_L_FD | IFOF_L | 24 | 0.418036118 | 2.373070305 | 0.677067872 | 0 |
| IFOF_L_FD | IFOF_L | 25 | -0.155475758 | 2.347868969 | 0.876841991 | 0 |
| IFOF_L_FD | IFOF_L | 26 | 0.013032483 | 2.403474234 | 0.989635575 | 0 |
| IFOF_L_FD | IFOF_L | 27 | -0.352267939 | 2.389254802 | 0.725651698 | 0 |
| IFOF_L_FD | IFOF_L | 28 | 0.10772012 | 2.421291386 | 0.914510552 | 0 |
| IFOF_L_FD | IFOF_L | 29 | 0.048328576 | 2.40900673 | 0.961582363 | 0 |
| IFOF_L_FD | IFOF_L | 30 | -0.14983554 | 2.424819964 | 0.881291244 | 0 |
| IFOF_L_FD | IFOF_L | 31 | 0.720367154 | 2.416700502 | 0.473630967 | 0 |
| IFOF_L_FD | IFOF_L | 32 | 1.003287218 | 2.379022679 | 0.319175864 | 0 |
| IFOF_L_FD | IFOF_L | 33 | 0.738869611 | 2.457109786 | 0.462388899 | 0 |
| IFOF_L_FD | IFOF_L | 34 | 0.155205399 | 2.359564908 | 0.877108787 | 0 |
| IFOF_L_FD | IFOF_L | 35 | -0.496868144 | 2.386330525 | 0.62081221 | 0 |
| IFOF_L_FD | IFOF_L | 36 | -0.316618861 | 2.406648381 | 0.752436651 | 0 |
| IFOF_L_FD | IFOF_L | 37 | 0.561407204 | 2.322795601 | 0.576220375 | 0 |
| IFOF_L_FD | IFOF_L | 38 | 1.429012533 | 2.364223665 | 0.157021637 | 0 |
| IFOF_L_FD | IFOF_L | 39 | 1.8619625 | 2.424105206 | 0.066486208 | 0 |
| IFOF_L_FD | IFOF_L | 40 | 1.366989171 | 2.384602428 | 0.175763109 | 0 |
| IFOF_L_FD | IFOF_L | 41 | 0.364056315 | 2.377718576 | 0.716775426 | 0 |
| IFOF_L_FD | IFOF_L | 42 | 0.812426806 | 2.352916293 | 0.419047385 | 0 |
| IFOF_L_FD | IFOF_L | 43 | 1.082770737 | 2.331476402 | 0.282426946 | 0 |
| IFOF_L_FD | IFOF_L | 44 | 1.407737183 | 2.347601126 | 0.163457854 | 0 |
| IFOF_L_FD | IFOF_L | 45 | 1.98256412 | 2.446316075 | 0.051536626 | 0 |
| IFOF_L_FD | IFOF_L | 46 | 2.163620171 | 2.372955043 | 0.033783773 | 0 |
| IFOF_L_FD | IFOF_L | 47 | 0.603491158 | 2.337741005 | 0.548031503 | 0 |
| IFOF_L_FD | IFOF_L | 48 | -0.199191596 | 2.395431139 | 0.842682062 | 0 |
| IFOF_L_FD | IFOF_L | 49 | -0.047197973 | 2.40356597 | 0.962479863 | 0 |
| IFOF_L_FD | IFOF_L | 50 | 0.760189238 | 2.34749263 | 0.449360037 | 0 |
| IFOF_R_FD | IFOF_R | 1 | -2.277301995 | 2.415668036 | 0.025781136 | 0 |
| IFOF_R_FD | IFOF_R | 2 | -0.7799019 | 2.418522373 | 0.438157855 | 0 |
| IFOF_R_FD | IFOF_R | 3 | 0.876562928 | 2.369424101 | 0.384340264 | 0 |
| IFOF_R_FD | IFOF_R | 4 | 1.928240115 | 2.35338737 | 0.058314585 | 0 |
| IFOF_R_FD | IFOF_R | 5 | 2.007305295 | 2.409572212 | 0.048873664 | 0 |
| IFOF_R_FD | IFOF_R | 6 | 1.214822014 | 2.394428596 | 0.228941665 | 0 |
| IFOF_R_FD | IFOF_R | 7 | 0.157103272 | 2.450792636 | 0.875728441 | 0 |

| | | | | | | |
|---|---|---|---|---|---|---|
| IFOF_R_FD | IFOF_R | 8 | -0.558112526 | 2.360921103 | 0.578853287 | 0 |
| IFOF_R_FD | IFOF_R | 9 | -0.686005469 | 2.338968878 | 0.495178147 | 0 |
| IFOF_R_FD | IFOF_R | 10 | -0.520004102 | 2.353515696 | 0.604874049 | 0 |
| IFOF_R_FD | IFOF_R | 11 | -0.344724287 | 2.342338748 | 0.731484126 | 0 |
| IFOF_R_FD | IFOF_R | 12 | -0.164604892 | 2.349218974 | 0.869740829 | 0 |
| IFOF_R_FD | IFOF_R | 13 | 0.164037259 | 2.378982142 | 0.8701955 | 0 |
| IFOF_R_FD | IFOF_R | 14 | 0.623752803 | 2.301096617 | 0.535044019 | 0 |
| IFOF_R_FD | IFOF_R | 15 | 0.630126782 | 2.403935212 | 0.530793411 | 0 |
| IFOF_R_FD | IFOF_R | 16 | 0.277402213 | 2.412717018 | 0.782263651 | 0 |
| IFOF_R_FD | IFOF_R | 17 | 0.270499873 | 2.336527935 | 0.787497355 | 0 |
| IFOF_R_FD | IFOF_R | 18 | 0.277653802 | 2.355603453 | 0.782022475 | 0 |
| IFOF_R_FD | IFOF_R | 19 | 0.48688508 | 2.365440332 | 0.627759995 | 0 |
| IFOF_R_FD | IFOF_R | 20 | 1.229598482 | 2.338285083 | 0.222734499 | 0 |
| IFOF_R_FD | IFOF_R | 21 | 1.510193527 | 2.436280113 | 0.135082093 | 0 |
| IFOF_R_FD | IFOF_R | 22 | 1.152821027 | 2.441579706 | 0.252388201 | 0 |
| IFOF_R_FD | IFOF_R | 23 | 0.409524024 | 2.389395763 | 0.683285276 | 0 |
| IFOF_R_FD | IFOF_R | 24 | -0.384443988 | 2.381644111 | 0.701795554 | 0 |
| IFOF_R_FD | IFOF_R | 25 | 0.012300371 | 2.352897235 | 0.990218802 | 0 |
| IFOF_R_FD | IFOF_R | 26 | 0.528139827 | 2.378590053 | 0.598974802 | 0 |
| IFOF_R_FD | IFOF_R | 27 | -0.30092924 | 2.361040124 | 0.764316161 | 0 |
| IFOF_R_FD | IFOF_R | 28 | -0.433140827 | 2.374212564 | 0.666180185 | 0 |
| IFOF_R_FD | IFOF_R | 29 | -0.740110949 | 2.383422878 | 0.461790442 | 0 |
| IFOF_R_FD | IFOF_R | 30 | -0.722383726 | 2.418425346 | 0.472822248 | 0 |
| IFOF_R_FD | IFOF_R | 31 | 0.281013558 | 2.451682076 | 0.779545428 | 0 |
| IFOF_R_FD | IFOF_R | 32 | 1.175223104 | 2.374709398 | 0.244014079 | 0 |
| IFOF_R_FD | IFOF_R | 33 | 1.21169667 | 2.396793059 | 0.229985342 | 0 |
| IFOF_R_FD | IFOF_R | 34 | 0.872803274 | 2.405000203 | 0.386084966 | 0 |
| IFOF_R_FD | IFOF_R | 35 | 1.34593966 | 2.399827827 | 0.18209829 | 0 |
| IFOF_R_FD | IFOF_R | 36 | 1.926124753 | 2.365507883 | 0.057740216 | 0 |
| IFOF_R_FD | IFOF_R | 37 | 1.886506862 | 2.401991385 | 0.062982898 | 0 |
| IFOF_R_FD | IFOF_R | 38 | 1.496602497 | 2.384082233 | 0.138601256 | 0 |
| IFOF_R_FD | IFOF_R | 39 | 1.824783131 | 2.381921375 | 0.071866269 | 0 |
| IFOF_R_FD | IFOF_R | 40 | 0.827497569 | 2.326082262 | 0.410644696 | 0 |
| IFOF_R_FD | IFOF_R | 41 | 0.111228143 | 2.421867172 | 0.911741678 | 0 |
| IFOF_R_FD | IFOF_R | 42 | -0.007125299 | 2.385501006 | 0.994333601 | 0 |
| IFOF_R_FD | IFOF_R | 43 | 1.054063738 | 2.406058366 | 0.295211874 | 0 |
| IFOF_R_FD | IFOF_R | 44 | 1.519980915 | 2.380113281 | 0.132760058 | 0 |
| IFOF_R_FD | IFOF_R | 45 | 0.78965685 | 2.418467854 | 0.432182274 | 0 |
| IFOF_R_FD | IFOF_R | 46 | 0.217665692 | 2.408875241 | 0.828263593 | 0 |
| IFOF_R_FD | IFOF_R | 47 | -0.495833721 | 2.326312553 | 0.621450559 | 0 |
| IFOF_R_FD | IFOF_R | 48 | -0.153280387 | 2.401199661 | 0.878600359 | 0 |
| IFOF_R_FD | IFOF_R | 49 | -0.033183099 | 2.391577284 | 0.973626047 | 0 |
| IFOF_R_FD | IFOF_R | 50 | 1.181774439 | 2.428990913 | 0.241079571 | 0 |

| | | | | | | |
|---|---|---|---|---|---|---|
| ILF_L_FD | ILF_L | 1 | 0.008230632 | 2.333609598 | 0.993455343 | 0 |
| ILF_L_FD | ILF_L | 2 | -0.303349839 | 2.370175464 | 0.76249112 | 0 |
| ILF_L_FD | ILF_L | 3 | 0.134272776 | 2.347072324 | 0.893545217 | 0 |
| ILF_L_FD | ILF_L | 4 | -0.259572177 | 2.440117208 | 0.795896903 | 0 |
| ILF_L_FD | ILF_L | 5 | -0.414654899 | 2.367902254 | 0.679572972 | 0 |
| ILF_L_FD | ILF_L | 6 | -0.180421269 | 2.422564107 | 0.85731566 | 0 |
| ILF_L_FD | ILF_L | 7 | -0.22715047 | 2.366804469 | 0.820940784 | 0 |
| ILF_L_FD | ILF_L | 8 | 0.263286371 | 2.381669851 | 0.793084255 | 0 |
| ILF_L_FD | ILF_L | 9 | 0.917095935 | 2.360257083 | 0.36203621 | 0 |
| ILF_L_FD | ILF_L | 10 | 0.766580374 | 2.373336257 | 0.445751352 | 0 |
| ILF_L_FD | ILF_L | 11 | 0.51732542 | 2.427440171 | 0.606537589 | 0 |
| ILF_L_FD | ILF_L | 12 | 0.540127531 | 2.358245848 | 0.590802482 | 0 |
| ILF_L_FD | ILF_L | 13 | 0.623100968 | 2.395309595 | 0.535251603 | 0 |
| ILF_L_FD | ILF_L | 14 | 0.915011887 | 2.417621226 | 0.363376694 | 0 |
| ILF_L_FD | ILF_L | 15 | 1.215158451 | 2.354636683 | 0.228387262 | 0 |
| ILF_L_FD | ILF_L | 16 | 1.417999889 | 2.407258069 | 0.160639503 | 0 |
| ILF_L_FD | ILF_L | 17 | 1.70872763 | 2.356447503 | 0.091902259 | 0 |
| ILF_L_FD | ILF_L | 18 | 2.070909759 | 2.358720622 | 0.042104963 | 0 |
| ILF_L_FD | ILF_L | 19 | 2.387248337 | 2.423133759 | 0.019727064 | 0 |
| ILF_L_FD | ILF_L | 20 | 2.421148642 | 2.349773463 | 0.018079811 | 1 |
| ILF_L_FD | ILF_L | 21 | 2.367642959 | 2.376876261 | 0.020688209 | 0 |
| ILF_L_FD | ILF_L | 22 | 2.289696582 | 2.342380405 | 0.025120073 | 0 |
| ILF_L_FD | ILF_L | 23 | 2.095990084 | 2.371837131 | 0.039868731 | 0 |
| ILF_L_FD | ILF_L | 24 | 1.746226048 | 2.377305798 | 0.08531191 | 0 |
| ILF_L_FD | ILF_L | 25 | 1.301645103 | 2.40180165 | 0.197366861 | 0 |
| ILF_L_FD | ILF_L | 26 | 0.992333091 | 2.413268572 | 0.324454387 | 0 |
| ILF_L_FD | ILF_L | 27 | 0.670162848 | 2.422703299 | 0.504849366 | 0 |
| ILF_L_FD | ILF_L | 28 | 0.449368483 | 2.35728487 | 0.654457503 | 0 |
| ILF_L_FD | ILF_L | 29 | 0.390328641 | 2.444071042 | 0.697380456 | 0 |
| ILF_L_FD | ILF_L | 30 | 0.247992041 | 2.347263611 | 0.804799282 | 0 |
| ILF_L_FD | ILF_L | 31 | 0.218568987 | 2.399548208 | 0.82759257 | 0 |
| ILF_L_FD | ILF_L | 32 | 0.386653152 | 2.376960633 | 0.700233067 | 0 |
| ILF_L_FD | ILF_L | 33 | 0.625837132 | 2.391757904 | 0.533572957 | 0 |
| ILF_L_FD | ILF_L | 34 | 0.92706439 | 2.372810169 | 0.357246719 | 0 |
| ILF_L_FD | ILF_L | 35 | 1.182469722 | 2.360709018 | 0.241328114 | 0 |
| ILF_L_FD | ILF_L | 36 | 1.085316252 | 2.3679767 | 0.281772248 | 0 |
| ILF_L_FD | ILF_L | 37 | 1.037631081 | 2.369064227 | 0.303133414 | 0 |
| ILF_L_FD | ILF_L | 38 | 0.938518652 | 2.394744422 | 0.351018432 | 0 |
| ILF_L_FD | ILF_L | 39 | 0.754647245 | 2.383849681 | 0.45284507 | 0 |
| ILF_L_FD | ILF_L | 40 | -0.030718248 | 2.391478622 | 0.975580011 | 0 |
| ILF_L_FD | ILF_L | 41 | -1.028718275 | 2.409524413 | 0.306942934 | 0 |
| ILF_L_FD | ILF_L | 42 | -1.380338912 | 2.464879824 | 0.171437707 | 0 |
| ILF_L_FD | ILF_L | 43 | -1.374722567 | 2.400590612 | 0.173148065 | 0 |

| | | | | | | |
|---|---|---|---|---|---|---|
| ILF_L_FD | ILF_L | 44 | -1.754138227 | 2.396039538 | 0.083251288 | 0 |
| ILF_L_FD | ILF_L | 45 | -1.975459631 | 2.39917004 | 0.051649739 | 0 |
| ILF_L_FD | ILF_L | 46 | -1.380966001 | 2.341639206 | 0.171274655 | 0 |
| ILF_L_FD | ILF_L | 47 | -0.905876988 | 2.404808212 | 0.367893097 | 0 |
| ILF_L_FD | ILF_L | 48 | -0.742999309 | 2.387831104 | 0.459670189 | 0 |
| ILF_L_FD | ILF_L | 49 | -0.408335755 | 2.387556838 | 0.684162055 | 0 |
| ILF_L_FD | ILF_L | 50 | -1.268860489 | 2.378279638 | 0.208327968 | 0 |
| ILF_R_FD | ILF_R | 1 | -0.520709785 | 2.430701595 | 0.604043549 | 0 |
| ILF_R_FD | ILF_R | 2 | 2.412629341 | 2.356372991 | 0.018419363 | 1 |
| ILF_R_FD | ILF_R | 3 | 2.998585001 | 2.405631975 | 0.003716146 | 1 |
| ILF_R_FD | ILF_R | 4 | 2.63426755 | 2.394744316 | 0.010283255 | 1 |
| ILF_R_FD | ILF_R | 5 | 1.967475955 | 2.313804384 | 0.053105514 | 0 |
| ILF_R_FD | ILF_R | 6 | 0.946695145 | 2.29026453 | 0.347300657 | 0 |
| ILF_R_FD | ILF_R | 7 | 0.312981967 | 2.335237893 | 0.755388965 | 0 |
| ILF_R_FD | ILF_R | 8 | -0.2639787 | 2.401937632 | 0.792693484 | 0 |
| ILF_R_FD | ILF_R | 9 | -0.916298178 | 2.443153575 | 0.362975824 | 0 |
| ILF_R_FD | ILF_R | 10 | -1.326079361 | 2.370655118 | 0.18941005 | 0 |
| ILF_R_FD | ILF_R | 11 | -1.915115208 | 2.391074906 | 0.059839874 | 0 |
| ILF_R_FD | ILF_R | 12 | -2.161177676 | 2.371544823 | 0.034321897 | 0 |
| ILF_R_FD | ILF_R | 13 | -1.995278485 | 2.323828018 | 0.050311242 | 0 |
| ILF_R_FD | ILF_R | 14 | -1.68536282 | 2.392791356 | 0.096858073 | 0 |
| ILF_R_FD | ILF_R | 15 | -1.538213305 | 2.322945522 | 0.128862722 | 0 |
| ILF_R_FD | ILF_R | 16 | -1.17556153 | 2.332002408 | 0.243896979 | 0 |
| ILF_R_FD | ILF_R | 17 | -0.989915481 | 2.433599682 | 0.325534891 | 0 |
| ILF_R_FD | ILF_R | 18 | -0.805077741 | 2.428983462 | 0.423372579 | 0 |
| ILF_R_FD | ILF_R | 19 | -0.357997393 | 2.369910139 | 0.721416139 | 0 |
| ILF_R_FD | ILF_R | 20 | 0.230395882 | 2.351272366 | 0.818464879 | 0 |
| ILF_R_FD | ILF_R | 21 | 0.750207055 | 2.386525627 | 0.455689095 | 0 |
| ILF_R_FD | ILF_R | 22 | 0.612636366 | 2.40972464 | 0.542205212 | 0 |
| ILF_R_FD | ILF_R | 23 | 0.292166914 | 2.390614456 | 0.771075243 | 0 |
| ILF_R_FD | ILF_R | 24 | 0.2805945 | 2.385843811 | 0.779830919 | 0 |
| ILF_R_FD | ILF_R | 25 | -0.048701201 | 2.354794006 | 0.961284099 | 0 |
| ILF_R_FD | ILF_R | 26 | -0.398500739 | 2.386700156 | 0.691370544 | 0 |
| ILF_R_FD | ILF_R | 27 | -0.441024584 | 2.449606301 | 0.660430897 | 0 |
| ILF_R_FD | ILF_R | 28 | -0.234363682 | 2.407061541 | 0.815313837 | 0 |
| ILF_R_FD | ILF_R | 29 | 0.002450204 | 2.368447709 | 0.998051219 | 0 |
| ILF_R_FD | ILF_R | 30 | 0.223982241 | 2.396669038 | 0.82334223 | 0 |
| ILF_R_FD | ILF_R | 31 | 0.372333077 | 2.396790147 | 0.710623864 | 0 |
| ILF_R_FD | ILF_R | 32 | 0.284256234 | 2.371118997 | 0.776958991 | 0 |
| ILF_R_FD | ILF_R | 33 | 0.338183381 | 2.341613335 | 0.736129221 | 0 |
| ILF_R_FD | ILF_R | 34 | 0.382257514 | 2.419590319 | 0.703329328 | 0 |
| ILF_R_FD | ILF_R | 35 | 0.372183783 | 2.409100395 | 0.710826285 | 0 |
| ILF_R_FD | ILF_R | 36 | -0.127646694 | 2.443342531 | 0.89876562 | 0 |

| | | | | | | |
|---|---|---|---|---|---|---|
| ILF_R_FD | ILF_R | 37 | -0.161510893 | 2.370393368 | 0.872107768 | 0 |
| ILF_R_FD | ILF_R | 38 | 0.369850136 | 2.384196484 | 0.712465797 | 0 |
| ILF_R_FD | ILF_R | 39 | 0.712407325 | 2.358843563 | 0.478279316 | 0 |
| ILF_R_FD | ILF_R | 40 | 0.265969165 | 2.454366944 | 0.790952657 | 0 |
| ILF_R_FD | ILF_R | 41 | 0.212006037 | 2.386141904 | 0.832637697 | 0 |
| ILF_R_FD | ILF_R | 42 | 0.068107581 | 2.39131923 | 0.945870538 | 0 |
| ILF_R_FD | ILF_R | 43 | -0.224986839 | 2.360232401 | 0.822580864 | 0 |
| ILF_R_FD | ILF_R | 44 | -0.914806796 | 2.373277096 | 0.363194646 | 0 |
| ILF_R_FD | ILF_R | 45 | -1.251782044 | 2.412439436 | 0.21446528 | 0 |
| ILF_R_FD | ILF_R | 46 | -1.279704491 | 2.321059991 | 0.204459607 | 0 |
| ILF_R_FD | ILF_R | 47 | -0.429889055 | 2.428269964 | 0.668443508 | 0 |
| ILF_R_FD | ILF_R | 48 | 0.024247977 | 2.372409485 | 0.980720799 | 0 |
| ILF_R_FD | ILF_R | 49 | 0.257826416 | 2.325383909 | 0.79726264 | 0 |
| ILF_R_FD | ILF_R | 50 | -0.118701232 | 2.388435142 | 0.905806116 | 0 |
| MCP_FD | MCP | 1 | -0.6776434 | 2.423408259 | 0.500543878 | 0 |
| MCP_FD | MCP | 2 | -0.489098146 | 2.41296779 | 0.626491144 | 0 |
| MCP_FD | MCP | 3 | -0.892852638 | 2.401870303 | 0.375494227 | 0 |
| MCP_FD | MCP | 4 | -1.051405121 | 2.357464561 | 0.296624642 | 0 |
| MCP_FD | MCP | 5 | -0.851997718 | 2.355351746 | 0.397051042 | 0 |
| MCP_FD | MCP | 6 | -0.70998466 | 2.389489911 | 0.479964492 | 0 |
| MCP_FD | MCP | 7 | -0.157847913 | 2.389069232 | 0.875000506 | 0 |
| MCP_FD | MCP | 8 | 0.166847652 | 2.364222614 | 0.867952663 | 0 |
| MCP_FD | MCP | 9 | 0.24730262 | 2.369191303 | 0.805401961 | 0 |
| MCP_FD | MCP | 10 | 0.163991251 | 2.362299338 | 0.870199183 | 0 |
| MCP_FD | MCP | 11 | 0.340739436 | 2.381721477 | 0.734275074 | 0 |
| MCP_FD | MCP | 12 | 0.229630735 | 2.346476992 | 0.819012891 | 0 |
| MCP_FD | MCP | 13 | 0.051176951 | 2.38674675 | 0.959335251 | 0 |
| MCP_FD | MCP | 14 | 0.479271905 | 2.334443762 | 0.633233739 | 0 |
| MCP_FD | MCP | 15 | 1.244736879 | 2.407998534 | 0.216994428 | 0 |
| MCP_FD | MCP | 16 | 1.873810459 | 2.408783918 | 0.06460971 | 0 |
| MCP_FD | MCP | 17 | 1.344169993 | 2.330879027 | 0.182722904 | 0 |
| MCP_FD | MCP | 18 | 0.303125006 | 2.403858843 | 0.762620903 | 0 |
| MCP_FD | MCP | 19 | -0.951716939 | 2.421500726 | 0.34433916 | 0 |
| MCP_FD | MCP | 20 | -0.490753242 | 2.376929545 | 0.624991613 | 0 |
| MCP_FD | MCP | 21 | -0.093940255 | 2.414996329 | 0.925393471 | 0 |
| MCP_FD | MCP | 22 | -1.388942531 | 2.366532912 | 0.169048618 | 0 |
| MCP_FD | MCP | 23 | -1.977121575 | 2.36764382 | 0.051887838 | 0 |
| MCP_FD | MCP | 24 | -1.56279358 | 2.410013926 | 0.122236204 | 0 |
| MCP_FD | MCP | 25 | -0.827252469 | 2.365363072 | 0.41067653 | 0 |
| MCP_FD | MCP | 26 | -1.954534718 | 2.434497271 | 0.054270082 | 0 |
| MCP_FD | MCP | 27 | -1.415297194 | 2.4028003 | 0.160877412 | 0 |
| MCP_FD | MCP | 28 | -0.680981288 | 2.394920356 | 0.497888432 | 0 |
| MCP_FD | MCP | 29 | 0.848670898 | 2.387836401 | 0.39871045 | 0 |

| | | | | | | |
|---|---|---|---|---|---|---|
| MCP_FD | MCP | 30 | 1.288506348 | 2.403874995 | 0.201358049 | 0 |
| MCP_FD | MCP | 31 | 1.288815337 | 2.370242565 | 0.201201183 | 0 |
| MCP_FD | MCP | 32 | 0.832370539 | 2.402648089 | 0.407730331 | 0 |
| MCP_FD | MCP | 33 | 0.472299376 | 2.378054385 | 0.638007175 | 0 |
| MCP_FD | MCP | 34 | 0.440641007 | 2.360530836 | 0.660700983 | 0 |
| MCP_FD | MCP | 35 | 0.066182429 | 2.437009886 | 0.947404568 | 0 |
| MCP_FD | MCP | 36 | -0.191899155 | 2.370875349 | 0.848337501 | 0 |
| MCP_FD | MCP | 37 | -0.311842559 | 2.366955322 | 0.756018366 | 0 |
| MCP_FD | MCP | 38 | -0.467875364 | 2.384316364 | 0.641198878 | 0 |
| MCP_FD | MCP | 39 | -0.549676664 | 2.406091788 | 0.584082798 | 0 |
| MCP_FD | MCP | 40 | -0.531776643 | 2.416633806 | 0.596340776 | 0 |
| MCP_FD | MCP | 41 | -0.283755707 | 2.406722745 | 0.777323464 | 0 |
| MCP_FD | MCP | 42 | -0.167528893 | 2.366969155 | 0.867375245 | 0 |
| MCP_FD | MCP | 43 | -0.472875548 | 2.388844961 | 0.637582621 | 0 |
| MCP_FD | MCP | 44 | -0.969365108 | 2.404593905 | 0.335254337 | 0 |
| MCP_FD | MCP | 45 | -1.389121142 | 2.387612447 | 0.168604117 | 0 |
| MCP_FD | MCP | 46 | -1.617661649 | 2.402392963 | 0.10965831 | 0 |
| MCP_FD | MCP | 47 | -1.975663291 | 2.440564027 | 0.051634869 | 0 |
| MCP_FD | MCP | 48 | -1.978385089 | 2.39773428 | 0.051338917 | 0 |
| MCP_FD | MCP | 49 | -0.780324216 | 2.366811093 | 0.438080162 | 0 |
| MCP_FD | MCP | 50 | -0.723211311 | 2.452774562 | 0.472260081 | 0 |
| OR_L_FD | OR_L | 1 | 0.175994868 | 2.349249123 | 0.860788332 | 0 |
| OR_L_FD | OR_L | 2 | 0.824689163 | 2.392067767 | 0.412577862 | 0 |
| OR_L_FD | OR_L | 3 | 0.79735087 | 2.340087211 | 0.42826092 | 0 |
| OR_L_FD | OR_L | 4 | 0.846523055 | 2.44051893 | 0.400401978 | 0 |
| OR_L_FD | OR_L | 5 | 0.574170936 | 2.376985476 | 0.567842647 | 0 |
| OR_L_FD | OR_L | 6 | 0.150880713 | 2.402002707 | 0.880564956 | 0 |
| OR_L_FD | OR_L | 7 | -0.001809329 | 2.359219109 | 0.998562195 | 0 |
| OR_L_FD | OR_L | 8 | 0.048612346 | 2.371068013 | 0.961381016 | 0 |
| OR_L_FD | OR_L | 9 | -0.189457651 | 2.45623117 | 0.85031546 | 0 |
| OR_L_FD | OR_L | 10 | -0.219473865 | 2.402970865 | 0.826938839 | 0 |
| OR_L_FD | OR_L | 11 | 0.18226997 | 2.384062089 | 0.855914508 | 0 |
| OR_L_FD | OR_L | 12 | 0.213836441 | 2.395773957 | 0.831259639 | 0 |
| OR_L_FD | OR_L | 13 | 0.392343628 | 2.373069931 | 0.695896839 | 0 |
| OR_L_FD | OR_L | 14 | 0.722843285 | 2.397997616 | 0.472029579 | 0 |
| OR_L_FD | OR_L | 15 | 0.903254106 | 2.381548278 | 0.369428544 | 0 |
| OR_L_FD | OR_L | 16 | 0.592552449 | 2.440884565 | 0.55556272 | 0 |
| OR_L_FD | OR_L | 17 | 0.396657374 | 2.400617303 | 0.692926139 | 0 |
| OR_L_FD | OR_L | 18 | 0.43433938 | 2.353022794 | 0.665288255 | 0 |
| OR_L_FD | OR_L | 19 | 0.580979416 | 2.40629664 | 0.562940603 | 0 |
| OR_L_FD | OR_L | 20 | 0.787995886 | 2.415052921 | 0.433125161 | 0 |
| OR_L_FD | OR_L | 21 | 1.136722419 | 2.372245404 | 0.259189102 | 0 |
| OR_L_FD | OR_L | 22 | 1.588911611 | 2.379343889 | 0.116451751 | 0 |

| | | | | | | |
|---|---|---|---|---|---|---|
| OR_L_FD | OR_L | 23 | 1.91838336 | 2.468684876 | 0.059137099 | 0 |
| OR_L_FD | OR_L | 24 | 2.234309403 | 2.384837391 | 0.028796678 | 0 |
| OR_L_FD | OR_L | 25 | 2.352803387 | 2.405354578 | 0.021618483 | 0 |
| OR_L_FD | OR_L | 26 | 2.10231478 | 2.399943651 | 0.039519445 | 0 |
| OR_L_FD | OR_L | 27 | 1.438544374 | 2.367867034 | 0.155268359 | 0 |
| OR_L_FD | OR_L | 28 | 1.012272021 | 2.412686182 | 0.315178431 | 0 |
| OR_L_FD | OR_L | 29 | 1.010434311 | 2.40160655 | 0.316004568 | 0 |
| OR_L_FD | OR_L | 30 | 0.894507025 | 2.400482288 | 0.374181419 | 0 |
| OR_L_FD | OR_L | 31 | 0.780930103 | 2.325687648 | 0.437444923 | 0 |
| OR_L_FD | OR_L | 32 | 0.657409835 | 2.369796884 | 0.513004227 | 0 |
| OR_L_FD | OR_L | 33 | 0.770167251 | 2.407581261 | 0.443618679 | 0 |
| OR_L_FD | OR_L | 34 | 0.897594943 | 2.39018023 | 0.37219359 | 0 |
| OR_L_FD | OR_L | 35 | 1.356269244 | 2.423900049 | 0.179084406 | 0 |
| OR_L_FD | OR_L | 36 | 1.802011699 | 2.376129693 | 0.07585199 | 0 |
| OR_L_FD | OR_L | 37 | 1.127363814 | 2.383634075 | 0.263658527 | 0 |
| OR_L_FD | OR_L | 38 | 0.141508398 | 2.410494963 | 0.887923804 | 0 |
| OR_L_FD | OR_L | 39 | 0.000686216 | 2.403824483 | 0.999454616 | 0 |
| OR_L_FD | OR_L | 40 | 0.630779822 | 2.382944859 | 0.530152591 | 0 |
| OR_L_FD | OR_L | 41 | 1.035715944 | 2.415832303 | 0.30352806 | 0 |
| OR_L_FD | OR_L | 42 | 1.517265441 | 2.414567162 | 0.133206964 | 0 |
| OR_L_FD | OR_L | 43 | 1.422585601 | 2.421089714 | 0.158904979 | 0 |
| OR_L_FD | OR_L | 44 | 0.842466581 | 2.341718929 | 0.402109923 | 0 |
| OR_L_FD | OR_L | 45 | 0.928076695 | 2.391236208 | 0.356135588 | 0 |
| OR_L_FD | OR_L | 46 | 0.625803379 | 2.352523921 | 0.533249454 | 0 |
| OR_L_FD | OR_L | 47 | 0.808072845 | 2.379838677 | 0.421676798 | 0 |
| OR_L_FD | OR_L | 48 | 1.287698458 | 2.425848274 | 0.201792934 | 0 |
| OR_L_FD | OR_L | 49 | 1.223326037 | 2.369487546 | 0.224998484 | 0 |
| OR_L_FD | OR_L | 50 | 0.891888487 | 2.347049467 | 0.37538506 | 0 |
| OR_R_FD | OR_R | 1 | 0.248430992 | 2.402008413 | 0.804442853 | 0 |
| OR_R_FD | OR_R | 2 | -0.373046693 | 2.425349232 | 0.710103549 | 0 |
| OR_R_FD | OR_R | 3 | -0.017474646 | 2.462133368 | 0.986101236 | 0 |
| OR_R_FD | OR_R | 4 | 0.440337666 | 2.358579598 | 0.66087822 | 0 |
| OR_R_FD | OR_R | 5 | 0.574822914 | 2.427501748 | 0.56705502 | 0 |
| OR_R_FD | OR_R | 6 | 0.448289378 | 2.373437388 | 0.655311913 | 0 |
| OR_R_FD | OR_R | 7 | -0.070939582 | 2.372968473 | 0.943655032 | 0 |
| OR_R_FD | OR_R | 8 | -0.798952 | 2.391612939 | 0.426876796 | 0 |
| OR_R_FD | OR_R | 9 | -0.988519452 | 2.413188093 | 0.326106806 | 0 |
| OR_R_FD | OR_R | 10 | -0.820751199 | 2.3613981 | 0.414583282 | 0 |
| OR_R_FD | OR_R | 11 | -0.793565136 | 2.381858426 | 0.429934903 | 0 |
| OR_R_FD | OR_R | 12 | -1.007028886 | 2.359288818 | 0.316964925 | 0 |
| OR_R_FD | OR_R | 13 | -1.242111692 | 2.412685534 | 0.217800982 | 0 |
| OR_R_FD | OR_R | 14 | -1.698505533 | 2.424633928 | 0.093424009 | 0 |
| OR_R_FD | OR_R | 15 | -1.919576578 | 2.397933442 | 0.058875232 | 0 |

| | | | | | | |
|---|---|---|---|---|---|---|
| OR_R_FD | OR_R | 16 | -1.874363656 | 2.448086618 | 0.065142771 | 0 |
| OR_R_FD | OR_R | 17 | -1.593099909 | 2.455216453 | 0.1157037 | 0 |
| OR_R_FD | OR_R | 18 | -1.403074388 | 2.400932287 | 0.164972004 | 0 |
| OR_R_FD | OR_R | 19 | -0.762141115 | 2.38152707 | 0.448482507 | 0 |
| OR_R_FD | OR_R | 20 | -0.187829165 | 2.416089235 | 0.851537434 | 0 |
| OR_R_FD | OR_R | 21 | 0.444201715 | 2.395467757 | 0.658252135 | 0 |
| OR_R_FD | OR_R | 22 | 0.543375905 | 2.404710516 | 0.588713217 | 0 |
| OR_R_FD | OR_R | 23 | 0.897534007 | 2.411948172 | 0.372822031 | 0 |
| OR_R_FD | OR_R | 24 | 1.601420935 | 2.411352353 | 0.114562022 | 0 |
| OR_R_FD | OR_R | 25 | 2.169367372 | 2.38623796 | 0.034091814 | 0 |
| OR_R_FD | OR_R | 26 | 2.31890264 | 2.397741587 | 0.023978364 | 0 |
| OR_R_FD | OR_R | 27 | 2.0097606 | 2.397138445 | 0.048889187 | 0 |
| OR_R_FD | OR_R | 28 | 1.60077462 | 2.370763254 | 0.114250267 | 0 |
| OR_R_FD | OR_R | 29 | 1.381780604 | 2.439074934 | 0.171734566 | 0 |
| OR_R_FD | OR_R | 30 | 1.410402444 | 2.351417726 | 0.163069981 | 0 |
| OR_R_FD | OR_R | 31 | 1.401689398 | 2.34623476 | 0.16548007 | 0 |
| OR_R_FD | OR_R | 32 | 0.914397323 | 2.424593452 | 0.363491902 | 0 |
| OR_R_FD | OR_R | 33 | 0.274578604 | 2.36437778 | 0.784394846 | 0 |
| OR_R_FD | OR_R | 34 | -0.272822639 | 2.395471391 | 0.785740186 | 0 |
| OR_R_FD | OR_R | 35 | -0.198451158 | 2.413807093 | 0.843237471 | 0 |
| OR_R_FD | OR_R | 36 | 0.052519312 | 2.359557057 | 0.95826724 | 0 |
| OR_R_FD | OR_R | 37 | 0.37829608 | 2.361889427 | 0.706460691 | 0 |
| OR_R_FD | OR_R | 38 | -0.154605886 | 2.403006745 | 0.877570445 | 0 |
| OR_R_FD | OR_R | 39 | -0.73447652 | 2.381881022 | 0.46482003 | 0 |
| OR_R_FD | OR_R | 40 | -0.261972205 | 2.421527291 | 0.794055954 | 0 |
| OR_R_FD | OR_R | 41 | 0.491912636 | 2.423385747 | 0.624136465 | 0 |
| OR_R_FD | OR_R | 42 | 1.069758088 | 2.345306507 | 0.287913433 | 0 |
| OR_R_FD | OR_R | 43 | 1.11577085 | 2.422991125 | 0.267868308 | 0 |
| OR_R_FD | OR_R | 44 | 0.046604681 | 2.370305169 | 0.962943395 | 0 |
| OR_R_FD | OR_R | 45 | -0.117402462 | 2.357266167 | 0.90684503 | 0 |
| OR_R_FD | OR_R | 46 | -0.269583609 | 2.392948979 | 0.788248401 | 0 |
| OR_R_FD | OR_R | 47 | 0.421394819 | 2.394920568 | 0.674667821 | 0 |
| OR_R_FD | OR_R | 48 | 1.166463085 | 2.338738559 | 0.246921904 | 0 |
| OR_R_FD | OR_R | 49 | 0.928837484 | 2.394127641 | 0.35596961 | 0 |
| OR_R_FD | OR_R | 50 | 0.817575662 | 2.375503038 | 0.416282599 | 0 |
| SCP_L_FD | SCP_L | 1 | -0.824898291 | 2.409559517 | 0.412061657 | 0 |
| SCP_L_FD | SCP_L | 2 | -0.854390746 | 2.363071045 | 0.39578406 | 0 |
| SCP_L_FD | SCP_L | 3 | -1.160044912 | 2.438135819 | 0.249855209 | 0 |
| SCP_L_FD | SCP_L | 4 | -1.723521701 | 2.386370253 | 0.08886502 | 0 |
| SCP_L_FD | SCP_L | 5 | -2.460002372 | 2.404221246 | 0.016055388 | 1 |
| SCP_L_FD | SCP_L | 6 | -3.083887666 | 2.394276467 | 0.002874778 | 1 |
| SCP_L_FD | SCP_L | 7 | -2.795605648 | 2.372141011 | 0.006619425 | 1 |
| SCP_L_FD | SCP_L | 8 | -1.926178719 | 2.370285832 | 0.058057813 | 0 |

| | | | | | | |
|---|---|---|---|---|---|---|
| SCP_L_FD | SCP_L | 9 | -1.375364344 | 2.361577002 | 0.17325838 | 0 |
| SCP_L_FD | SCP_L | 10 | -0.780299911 | 2.365072853 | 0.437587698 | 0 |
| SCP_L_FD | SCP_L | 11 | -0.594054389 | 2.361303078 | 0.554140568 | 0 |
| SCP_L_FD | SCP_L | 12 | -0.964540813 | 2.413094768 | 0.337649062 | 0 |
| SCP_L_FD | SCP_L | 13 | -1.48983579 | 2.408006087 | 0.140151812 | 0 |
| SCP_L_FD | SCP_L | 14 | -1.632564649 | 2.408524548 | 0.106448195 | 0 |
| SCP_L_FD | SCP_L | 15 | -1.51952027 | 2.437210552 | 0.132532692 | 0 |
| SCP_L_FD | SCP_L | 16 | -1.29451486 | 2.43946377 | 0.199165931 | 0 |
| SCP_L_FD | SCP_L | 17 | -1.070879294 | 2.494710427 | 0.28744292 | 0 |
| SCP_L_FD | SCP_L | 18 | -0.886947041 | 2.442561451 | 0.377806055 | 0 |
| SCP_L_FD | SCP_L | 19 | -0.525043753 | 2.440640116 | 0.601061549 | 0 |
| SCP_L_FD | SCP_L | 20 | -0.306271898 | 2.437689008 | 0.760226504 | 0 |
| SCP_L_FD | SCP_L | 21 | -0.021574042 | 2.454893065 | 0.982844107 | 0 |
| SCP_L_FD | SCP_L | 22 | 0.031794742 | 2.475957451 | 0.974719842 | 0 |
| SCP_L_FD | SCP_L | 23 | 0.037149409 | 2.426125089 | 0.970468163 | 0 |
| SCP_L_FD | SCP_L | 24 | -0.047075799 | 2.428661363 | 0.962582324 | 0 |
| SCP_L_FD | SCP_L | 25 | -0.083796183 | 2.380556258 | 0.933451913 | 0 |
| SCP_L_FD | SCP_L | 26 | 0.070062726 | 2.396215012 | 0.944349537 | 0 |
| SCP_L_FD | SCP_L | 27 | 0.188483508 | 2.443356453 | 0.851086063 | 0 |
| SCP_L_FD | SCP_L | 28 | 0.405744833 | 2.415162808 | 0.686299068 | 0 |
| SCP_L_FD | SCP_L | 29 | 0.539651033 | 2.386859234 | 0.591344679 | 0 |
| SCP_L_FD | SCP_L | 30 | 0.710489962 | 2.450954036 | 0.47996101 | 0 |
| SCP_L_FD | SCP_L | 31 | 0.829334946 | 2.427433367 | 0.409891618 | 0 |
| SCP_L_FD | SCP_L | 32 | 0.904109042 | 2.285659758 | 0.369132617 | 0 |
| SCP_L_FD | SCP_L | 33 | 1.18872067 | 2.328934882 | 0.238469864 | 0 |
| SCP_L_FD | SCP_L | 34 | 1.173308425 | 2.361299579 | 0.244445775 | 0 |
| SCP_L_FD | SCP_L | 35 | 1.366068921 | 2.387470831 | 0.175965049 | 0 |
| SCP_L_FD | SCP_L | 36 | 1.408533863 | 2.378847712 | 0.16301585 | 0 |
| SCP_L_FD | SCP_L | 37 | 1.748649403 | 2.37610923 | 0.08435888 | 0 |
| SCP_L_FD | SCP_L | 38 | 1.672168771 | 2.357540711 | 0.098558394 | 0 |
| SCP_L_FD | SCP_L | 39 | 1.632128956 | 2.337356906 | 0.106651283 | 0 |
| SCP_L_FD | SCP_L | 40 | 1.543500213 | 2.36212719 | 0.126707357 | 0 |
| SCP_L_FD | SCP_L | 41 | 1.402373695 | 2.438240232 | 0.164798539 | 0 |
| SCP_L_FD | SCP_L | 42 | 1.260979578 | 2.436943015 | 0.211085876 | 0 |
| SCP_L_FD | SCP_L | 43 | 1.151288861 | 2.358672023 | 0.253099588 | 0 |
| SCP_L_FD | SCP_L | 44 | 1.280049973 | 2.395599064 | 0.204312047 | 0 |
| SCP_L_FD | SCP_L | 45 | 1.523432612 | 2.389089838 | 0.131809361 | 0 |
| SCP_L_FD | SCP_L | 46 | 1.800013522 | 2.354370404 | 0.076202009 | 0 |
| SCP_L_FD | SCP_L | 47 | 1.613851127 | 2.388362878 | 0.111224293 | 0 |
| SCP_L_FD | SCP_L | 48 | 1.677288578 | 2.43672068 | 0.098312174 | 0 |
| SCP_L_FD | SCP_L | 49 | 1.374033597 | 2.38540224 | 0.174214414 | 0 |
| SCP_L_FD | SCP_L | 50 | 0.704740432 | 2.385841599 | 0.483142161 | 0 |
| SCP_R_FD | SCP_R | 1 | -1.068313699 | 2.227358762 | 0.28890737 | 0 |

| | | | | | | |
|---|---|---|---|---|---|---|
| SCP_R_FD | SCP_R | 2 | -1.459602627 | 2.321031949 | 0.148383683 | 0 |
| SCP_R_FD | SCP_R | 3 | -1.169547892 | 2.350858383 | 0.245696923 | 0 |
| SCP_R_FD | SCP_R | 4 | -1.834745295 | 2.431922447 | 0.070220243 | 0 |
| SCP_R_FD | SCP_R | 5 | -2.109949344 | 2.376839078 | 0.037994209 | 0 |
| SCP_R_FD | SCP_R | 6 | -2.078179714 | 2.380219485 | 0.04095939 | 0 |
| SCP_R_FD | SCP_R | 7 | -1.951619648 | 2.408974569 | 0.054757879 | 0 |
| SCP_R_FD | SCP_R | 8 | -2.044206562 | 2.369509376 | 0.044506989 | 0 |
| SCP_R_FD | SCP_R | 9 | -1.670329212 | 2.446630422 | 0.099063388 | 0 |
| SCP_R_FD | SCP_R | 10 | -1.426150739 | 2.334080201 | 0.15793105 | 0 |
| SCP_R_FD | SCP_R | 11 | -1.582117511 | 2.367905279 | 0.118006848 | 0 |
| SCP_R_FD | SCP_R | 12 | -1.850631838 | 2.331102289 | 0.068450736 | 0 |
| SCP_R_FD | SCP_R | 13 | -1.832214693 | 2.407241852 | 0.070976201 | 0 |
| SCP_R_FD | SCP_R | 14 | -1.656070026 | 2.421300058 | 0.102156932 | 0 |
| SCP_R_FD | SCP_R | 15 | -1.644799695 | 2.361043769 | 0.104789071 | 0 |
| SCP_R_FD | SCP_R | 16 | -1.797908988 | 2.347282403 | 0.077153505 | 0 |
| SCP_R_FD | SCP_R | 17 | -1.831511318 | 2.385500484 | 0.072000531 | 0 |
| SCP_R_FD | SCP_R | 18 | -1.605044323 | 2.449504458 | 0.113376964 | 0 |
| SCP_R_FD | SCP_R | 19 | -1.407208945 | 2.436239425 | 0.16392365 | 0 |
| SCP_R_FD | SCP_R | 20 | -1.33429459 | 2.432867897 | 0.186531673 | 0 |
| SCP_R_FD | SCP_R | 21 | -0.992748184 | 2.431126386 | 0.324170034 | 0 |
| SCP_R_FD | SCP_R | 22 | -0.678775527 | 2.4098308 | 0.499478762 | 0 |
| SCP_R_FD | SCP_R | 23 | -0.563533826 | 2.468822504 | 0.574819199 | 0 |
| SCP_R_FD | SCP_R | 24 | -0.426338464 | 2.38705975 | 0.671071347 | 0 |
| SCP_R_FD | SCP_R | 25 | -0.451012673 | 2.387590939 | 0.653238113 | 0 |
| SCP_R_FD | SCP_R | 26 | -0.493559393 | 2.409683211 | 0.622982965 | 0 |
| SCP_R_FD | SCP_R | 27 | -0.398030926 | 2.443612808 | 0.691661512 | 0 |
| SCP_R_FD | SCP_R | 28 | -0.229094993 | 2.402803058 | 0.819372548 | 0 |
| SCP_R_FD | SCP_R | 29 | -0.323832567 | 2.380977571 | 0.746899474 | 0 |
| SCP_R_FD | SCP_R | 30 | -0.328092067 | 2.406386489 | 0.743690336 | 0 |
| SCP_R_FD | SCP_R | 31 | -0.164969378 | 2.393223613 | 0.869380343 | 0 |
| SCP_R_FD | SCP_R | 32 | 0.089773031 | 2.381630906 | 0.92869197 | 0 |
| SCP_R_FD | SCP_R | 33 | 0.260812588 | 2.345255768 | 0.794920256 | 0 |
| SCP_R_FD | SCP_R | 34 | 0.486722305 | 2.407591276 | 0.627814293 | 0 |
| SCP_R_FD | SCP_R | 35 | 0.724383583 | 2.401578537 | 0.470982604 | 0 |
| SCP_R_FD | SCP_R | 36 | 0.645119127 | 2.386882034 | 0.520764951 | 0 |
| SCP_R_FD | SCP_R | 37 | 0.656910047 | 2.392490267 | 0.513214571 | 0 |
| SCP_R_FD | SCP_R | 38 | 0.862997001 | 2.389297023 | 0.390961039 | 0 |
| SCP_R_FD | SCP_R | 39 | 1.059731798 | 2.395947511 | 0.292909672 | 0 |
| SCP_R_FD | SCP_R | 40 | 1.18431714 | 2.333408436 | 0.240356613 | 0 |
| SCP_R_FD | SCP_R | 41 | 1.078028515 | 2.343895118 | 0.284661201 | 0 |
| SCP_R_FD | SCP_R | 42 | 1.087835172 | 2.434441863 | 0.280152243 | 0 |
| SCP_R_FD | SCP_R | 43 | 0.984791504 | 2.374459382 | 0.32783625 | 0 |
| SCP_R_FD | SCP_R | 44 | 0.667341193 | 2.363885076 | 0.506591128 | 0 |

| | | | | | | |
|---|---|---|---|---|---|---|
| SCP_R_FD | SCP_R | 45 | 0.703058981 | 2.35755376 | 0.484224015 | 0 |
| SCP_R_FD | SCP_R | 46 | 0.818814112 | 2.374097161 | 0.41560793 | 0 |
| SCP_R_FD | SCP_R | 47 | 0.571664763 | 2.33370424 | 0.56926939 | 0 |
| SCP_R_FD | SCP_R | 48 | 0.699475156 | 2.393979295 | 0.486423225 | 0 |
| SCP_R_FD | SCP_R | 49 | 0.65874859 | 2.396274986 | 0.51207769 | 0 |
| SCP_R_FD | SCP_R | 50 | 1.078776275 | 2.384841493 | 0.284210716 | 0 |
| SLF_1_L_FD | SLF_1_L | 1 | -0.209430368 | 2.385900109 | 0.834668264 | 0 |
| SLF_1_L_FD | SLF_1_L | 2 | -0.191743339 | 2.383405539 | 0.848448872 | 0 |
| SLF_1_L_FD | SLF_1_L | 3 | 1.190233261 | 2.389422045 | 0.237597379 | 0 |
| SLF_1_L_FD | SLF_1_L | 4 | -0.39448223 | 2.422507758 | 0.694301524 | 0 |
| SLF_1_L_FD | SLF_1_L | 5 | 0.288972012 | 2.3691301 | 0.773369433 | 0 |
| SLF_1_L_FD | SLF_1_L | 6 | -0.869759889 | 2.371753799 | 0.387304838 | 0 |
| SLF_1_L_FD | SLF_1_L | 7 | 0.090458816 | 2.381781383 | 0.928153443 | 0 |
| SLF_1_L_FD | SLF_1_L | 8 | -0.379239652 | 2.390628254 | 0.705538107 | 0 |
| SLF_1_L_FD | SLF_1_L | 9 | -0.793126076 | 2.351747265 | 0.430112458 | 0 |
| SLF_1_L_FD | SLF_1_L | 10 | -0.295883722 | 2.380661163 | 0.768107981 | 0 |
| SLF_1_L_FD | SLF_1_L | 11 | -1.487104421 | 2.404265194 | 0.140989236 | 0 |
| SLF_1_L_FD | SLF_1_L | 12 | -0.168132387 | 2.37928309 | 0.866919911 | 0 |
| SLF_1_L_FD | SLF_1_L | 13 | 0.608590457 | 2.420167432 | 0.544712129 | 0 |
| SLF_1_L_FD | SLF_1_L | 14 | 0.74368368 | 2.385001619 | 0.459810127 | 0 |
| SLF_1_L_FD | SLF_1_L | 15 | 1.495835289 | 2.407029824 | 0.139804443 | 0 |
| SLF_1_L_FD | SLF_1_L | 16 | 1.330332946 | 2.348444798 | 0.187443095 | 0 |
| SLF_1_L_FD | SLF_1_L | 17 | 0.569291547 | 2.379836114 | 0.570758406 | 0 |
| SLF_1_L_FD | SLF_1_L | 18 | -0.215427494 | 2.396130707 | 0.830039464 | 0 |
| SLF_1_L_FD | SLF_1_L | 19 | 1.179701716 | 2.416065631 | 0.241790587 | 0 |
| SLF_1_L_FD | SLF_1_L | 20 | 0.646290025 | 2.370149897 | 0.519979737 | 0 |
| SLF_1_L_FD | SLF_1_L | 21 | 0.349380144 | 2.372718639 | 0.727737708 | 0 |
| SLF_1_L_FD | SLF_1_L | 22 | -0.068972679 | 2.417920894 | 0.945183485 | 0 |
| SLF_1_L_FD | SLF_1_L | 23 | 0.062980617 | 2.396356313 | 0.949939451 | 0 |
| SLF_1_L_FD | SLF_1_L | 24 | 0.454526156 | 2.383742858 | 0.650693039 | 0 |
| SLF_1_L_FD | SLF_1_L | 25 | -0.780379789 | 2.439247502 | 0.437489579 | 0 |
| SLF_1_L_FD | SLF_1_L | 26 | -0.462398823 | 2.432546138 | 0.64506715 | 0 |
| SLF_1_L_FD | SLF_1_L | 27 | -0.314701896 | 2.379764673 | 0.753848573 | 0 |
| SLF_1_L_FD | SLF_1_L | 28 | -0.152825558 | 2.421283297 | 0.878942872 | 0 |
| SLF_1_L_FD | SLF_1_L | 29 | 0.348022305 | 2.347926084 | 0.728881429 | 0 |
| SLF_1_L_FD | SLF_1_L | 30 | 0.43359349 | 2.385739731 | 0.665816287 | 0 |
| SLF_1_L_FD | SLF_1_L | 31 | 1.154630317 | 2.358458625 | 0.252170563 | 0 |
| SLF_1_L_FD | SLF_1_L | 32 | 1.760008028 | 2.405570514 | 0.082695626 | 0 |
| SLF_1_L_FD | SLF_1_L | 33 | 1.008189453 | 2.367997437 | 0.31697652 | 0 |
| SLF_1_L_FD | SLF_1_L | 34 | 0.104458399 | 2.411843032 | 0.917085155 | 0 |
| SLF_1_L_FD | SLF_1_L | 35 | 0.608876032 | 2.39651807 | 0.544573604 | 0 |
| SLF_1_L_FD | SLF_1_L | 36 | 0.048102696 | 2.370166362 | 0.961782831 | 0 |
| SLF_1_L_FD | SLF_1_L | 37 | 0.82538974 | 2.354578242 | 0.411762182 | 0 |

| | | | | | | |
|---|---|---|---|---|---|---|
| SLF_1_L_FD | SLF_1_L | 38 | 1.178880812 | 2.420169003 | 0.242607653 | 0 |
| SLF_1_L_FD | SLF_1_L | 39 | 1.629475377 | 2.430701691 | 0.107750246 | 0 |
| SLF_1_L_FD | SLF_1_L | 40 | 0.361559776 | 2.364935904 | 0.71888893 | 0 |
| SLF_1_L_FD | SLF_1_L | 41 | -0.542593776 | 2.402370209 | 0.588999353 | 0 |
| SLF_1_L_FD | SLF_1_L | 42 | 0.597064891 | 2.390452536 | 0.552316696 | 0 |
| SLF_1_L_FD | SLF_1_L | 43 | 0.905526892 | 2.403901494 | 0.369280275 | 0 |
| SLF_1_L_FD | SLF_1_L | 44 | 0.487148063 | 2.42622088 | 0.627895629 | 0 |
| SLF_1_L_FD | SLF_1_L | 45 | -0.639048839 | 2.334654628 | 0.524817551 | 0 |
| SLF_1_L_FD | SLF_1_L | 46 | -0.855055867 | 2.362028002 | 0.395167133 | 0 |
| SLF_1_L_FD | SLF_1_L | 47 | -1.714110518 | 2.428687243 | 0.090394175 | 0 |
| SLF_1_L_FD | SLF_1_L | 48 | -0.804019995 | 2.340811696 | 0.423970733 | 0 |
| SLF_1_L_FD | SLF_1_L | 49 | -2.007520023 | 2.395609446 | 0.048237791 | 0 |
| SLF_1_L_FD | SLF_1_L | 50 | -1.498957949 | 2.366032405 | 0.138272636 | 0 |
| SLF_1_R_FD | SLF_1_R | 1 | -0.746038652 | 2.400937299 | 0.458145935 | 0 |
| SLF_1_R_FD | SLF_1_R | 2 | 0.258261759 | 2.382492914 | 0.796969784 | 0 |
| SLF_1_R_FD | SLF_1_R | 3 | -0.016134511 | 2.295214061 | 0.987171325 | 0 |
| SLF_1_R_FD | SLF_1_R | 4 | 0.425240561 | 2.346154997 | 0.6720415 | 0 |
| SLF_1_R_FD | SLF_1_R | 5 | 1.527678614 | 2.401191777 | 0.130773135 | 0 |
| SLF_1_R_FD | SLF_1_R | 6 | 1.211147997 | 2.367189597 | 0.229841689 | 0 |
| SLF_1_R_FD | SLF_1_R | 7 | 0.126585869 | 2.383451568 | 0.899620855 | 0 |
| SLF_1_R_FD | SLF_1_R | 8 | -0.116421565 | 2.37462764 | 0.907627359 | 0 |
| SLF_1_R_FD | SLF_1_R | 9 | -0.7123392 | 2.395371501 | 0.478339579 | 0 |
| SLF_1_R_FD | SLF_1_R | 10 | -0.622344629 | 2.410926787 | 0.535496626 | 0 |
| SLF_1_R_FD | SLF_1_R | 11 | -0.938197055 | 2.38758021 | 0.351000471 | 0 |
| SLF_1_R_FD | SLF_1_R | 12 | -0.646672616 | 2.374721115 | 0.519785009 | 0 |
| SLF_1_R_FD | SLF_1_R | 13 | 0.03469225 | 2.406115762 | 0.97241773 | 0 |
| SLF_1_R_FD | SLF_1_R | 14 | 0.449249372 | 2.366800448 | 0.654724796 | 0 |
| SLF_1_R_FD | SLF_1_R | 15 | 0.418495061 | 2.373474781 | 0.6770507 | 0 |
| SLF_1_R_FD | SLF_1_R | 16 | 0.946497468 | 2.388705945 | 0.347236564 | 0 |
| SLF_1_R_FD | SLF_1_R | 17 | 1.200605635 | 2.382701962 | 0.233896194 | 0 |
| SLF_1_R_FD | SLF_1_R | 18 | 0.238676033 | 2.426901224 | 0.812042348 | 0 |
| SLF_1_R_FD | SLF_1_R | 19 | 0.775980047 | 2.39902051 | 0.440163496 | 0 |
| SLF_1_R_FD | SLF_1_R | 20 | 1.163312641 | 2.419482055 | 0.248307105 | 0 |
| SLF_1_R_FD | SLF_1_R | 21 | 1.012934588 | 2.379799402 | 0.314146788 | 0 |
| SLF_1_R_FD | SLF_1_R | 22 | 0.308731798 | 2.348372349 | 0.758341612 | 0 |
| SLF_1_R_FD | SLF_1_R | 23 | -0.43641157 | 2.389726807 | 0.663774254 | 0 |
| SLF_1_R_FD | SLF_1_R | 24 | -0.530977915 | 2.396621158 | 0.597112536 | 0 |
| SLF_1_R_FD | SLF_1_R | 25 | -0.145168601 | 2.434768452 | 0.884965303 | 0 |
| SLF_1_R_FD | SLF_1_R | 26 | 0.556548832 | 2.381411412 | 0.579440938 | 0 |
| SLF_1_R_FD | SLF_1_R | 27 | 0.217029174 | 2.367901273 | 0.828768821 | 0 |
| SLF_1_R_FD | SLF_1_R | 28 | 0.001930276 | 2.372087111 | 0.998464881 | 0 |
| SLF_1_R_FD | SLF_1_R | 29 | -0.72223853 | 2.383832072 | 0.472391839 | 0 |
| SLF_1_R_FD | SLF_1_R | 30 | -0.504906113 | 2.378347217 | 0.615203088 | 0 |

| | | | | | | |
|---|---|---|---|---|---|---|
| SLF_1_R_FD | SLF_1_R | 31 | 0.548179842 | 2.350022365 | 0.585206816 | 0 |
| SLF_1_R_FD | SLF_1_R | 32 | 0.74564308 | 2.425625342 | 0.458140748 | 0 |
| SLF_1_R_FD | SLF_1_R | 33 | 0.634619303 | 2.386012758 | 0.527854871 | 0 |
| SLF_1_R_FD | SLF_1_R | 34 | 0.779831215 | 2.38555056 | 0.43806293 | 0 |
| SLF_1_R_FD | SLF_1_R | 35 | 1.775037361 | 2.401021087 | 0.0801102 | 0 |
| SLF_1_R_FD | SLF_1_R | 36 | 1.923618959 | 2.366425359 | 0.058061732 | 0 |
| SLF_1_R_FD | SLF_1_R | 37 | 0.607397847 | 2.393463228 | 0.5454749 | 0 |
| SLF_1_R_FD | SLF_1_R | 38 | 0.850982047 | 2.407700225 | 0.397365221 | 0 |
| SLF_1_R_FD | SLF_1_R | 39 | 0.556584028 | 2.350791408 | 0.57947927 | 0 |
| SLF_1_R_FD | SLF_1_R | 40 | 1.247254315 | 2.387071738 | 0.216438158 | 0 |
| SLF_1_R_FD | SLF_1_R | 41 | 0.651590122 | 2.365670786 | 0.516804058 | 0 |
| SLF_1_R_FD | SLF_1_R | 42 | 1.059113723 | 2.405537572 | 0.293109445 | 0 |
| SLF_1_R_FD | SLF_1_R | 43 | -0.231823033 | 2.358335964 | 0.817290052 | 0 |
| SLF_1_R_FD | SLF_1_R | 44 | -0.816271838 | 2.414551111 | 0.416820179 | 0 |
| SLF_1_R_FD | SLF_1_R | 45 | -0.204535881 | 2.362685817 | 0.838481217 | 0 |
| SLF_1_R_FD | SLF_1_R | 46 | 0.920332777 | 2.370790299 | 0.360803439 | 0 |
| SLF_1_R_FD | SLF_1_R | 47 | 1.073181836 | 2.368249849 | 0.286886604 | 0 |
| SLF_1_R_FD | SLF_1_R | 48 | 0.400319884 | 2.39392699 | 0.69004876 | 0 |
| SLF_1_R_FD | SLF_1_R | 49 | -0.959662728 | 2.426647824 | 0.340225295 | 0 |
| SLF_1_R_FD | SLF_1_R | 50 | -1.302392529 | 2.359474149 | 0.196589072 | 0 |
| SLF_2_L_FD | SLF_2_L | 1 | 1.379623312 | 2.383976766 | 0.172068741 | 0 |
| SLF_2_L_FD | SLF_2_L | 2 | 1.236463232 | 2.374079405 | 0.22031456 | 0 |
| SLF_2_L_FD | SLF_2_L | 3 | 0.78402725 | 2.409142744 | 0.435524587 | 0 |
| SLF_2_L_FD | SLF_2_L | 4 | 0.521210792 | 2.394414922 | 0.603718819 | 0 |
| SLF_2_L_FD | SLF_2_L | 5 | -0.193780288 | 2.324976586 | 0.846866059 | 0 |
| SLF_2_L_FD | SLF_2_L | 6 | 0.338935012 | 2.380704471 | 0.735844733 | 0 |
| SLF_2_L_FD | SLF_2_L | 7 | 2.50382338 | 2.436304833 | 0.01462883 | 1 |
| SLF_2_L_FD | SLF_2_L | 8 | 2.865897764 | 2.36059936 | 0.005575268 | 1 |
| SLF_2_L_FD | SLF_2_L | 9 | 1.357087158 | 2.390282401 | 0.179341297 | 0 |
| SLF_2_L_FD | SLF_2_L | 10 | 0.102591796 | 2.366941758 | 0.918567592 | 0 |
| SLF_2_L_FD | SLF_2_L | 11 | -1.033020912 | 2.403012117 | 0.304732692 | 0 |
| SLF_2_L_FD | SLF_2_L | 12 | -0.56783115 | 2.419836041 | 0.571753513 | 0 |
| SLF_2_L_FD | SLF_2_L | 13 | 0.067928741 | 2.385626795 | 0.946012176 | 0 |
| SLF_2_L_FD | SLF_2_L | 14 | 1.550232614 | 2.418119514 | 0.125109408 | 0 |
| SLF_2_L_FD | SLF_2_L | 15 | 2.980143262 | 2.348330493 | 0.003881882 | 1 |
| SLF_2_L_FD | SLF_2_L | 16 | 4.072029543 | 2.31110586 | 0.000109651 | 1 |
| SLF_2_L_FD | SLF_2_L | 17 | 5.286516627 | 2.355582483 | 1.06E-06 | 1 |
| SLF_2_L_FD | SLF_2_L | 18 | 4.789389099 | 2.364037557 | 7.52E-06 | 1 |
| SLF_2_L_FD | SLF_2_L | 19 | 3.853684161 | 2.426823299 | 0.000252542 | 1 |
| SLF_2_L_FD | SLF_2_L | 20 | 2.960510773 | 2.375274991 | 0.004231957 | 1 |
| SLF_2_L_FD | SLF_2_L | 21 | 2.267015988 | 2.378098114 | 0.026438256 | 0 |
| SLF_2_L_FD | SLF_2_L | 22 | 2.083605504 | 2.433414618 | 0.040668051 | 0 |
| SLF_2_L_FD | SLF_2_L | 23 | 2.520890819 | 2.388364456 | 0.013752927 | 1 |

| | | | | | | |
|---|---|---|---|---|---|---|
| SLF_2_L_FD | SLF_2_L | 24 | 1.816757006 | 2.422159494 | 0.073428716 | 0 |
| SLF_2_L_FD | SLF_2_L | 25 | 0.732552373 | 2.334059255 | 0.466441788 | 0 |
| SLF_2_L_FD | SLF_2_L | 26 | 0.485475053 | 2.400981429 | 0.628854907 | 0 |
| SLF_2_L_FD | SLF_2_L | 27 | 0.418950567 | 2.370901174 | 0.676485923 | 0 |
| SLF_2_L_FD | SLF_2_L | 28 | 0.895925759 | 2.382254255 | 0.3730616 | 0 |
| SLF_2_L_FD | SLF_2_L | 29 | 1.242316952 | 2.343577016 | 0.2177738 | 0 |
| SLF_2_L_FD | SLF_2_L | 30 | 1.837955324 | 2.357622276 | 0.069826364 | 0 |
| SLF_2_L_FD | SLF_2_L | 31 | 2.169890309 | 2.449718538 | 0.032975857 | 0 |
| SLF_2_L_FD | SLF_2_L | 32 | 2.018324885 | 2.397122968 | 0.046981721 | 0 |
| SLF_2_L_FD | SLF_2_L | 33 | 2.116452785 | 2.398762058 | 0.037529551 | 0 |
| SLF_2_L_FD | SLF_2_L | 34 | 2.089073686 | 2.412554471 | 0.040512038 | 0 |
| SLF_2_L_FD | SLF_2_L | 35 | 1.861568959 | 2.416167226 | 0.067000195 | 0 |
| SLF_2_L_FD | SLF_2_L | 36 | 1.979714348 | 2.356326323 | 0.051319636 | 0 |
| SLF_2_L_FD | SLF_2_L | 37 | 3.417784724 | 2.442166076 | 0.0009949 | 1 |
| SLF_2_L_FD | SLF_2_L | 38 | 3.000291263 | 2.373677863 | 0.003667061 | 1 |
| SLF_2_L_FD | SLF_2_L | 39 | 2.141351859 | 2.404263392 | 0.035760484 | 0 |
| SLF_2_L_FD | SLF_2_L | 40 | 1.43355791 | 2.376260275 | 0.155782647 | 0 |
| SLF_2_L_FD | SLF_2_L | 41 | 1.920868418 | 2.392116346 | 0.058402263 | 0 |
| SLF_2_L_FD | SLF_2_L | 42 | 1.927952887 | 2.310189561 | 0.057432331 | 0 |
| SLF_2_L_FD | SLF_2_L | 43 | 1.874458407 | 2.376883691 | 0.064725434 | 0 |
| SLF_2_L_FD | SLF_2_L | 44 | 1.8640267 | 2.390062837 | 0.066300999 | 0 |
| SLF_2_L_FD | SLF_2_L | 45 | 0.270344082 | 2.409700726 | 0.787585781 | 0 |
| SLF_2_L_FD | SLF_2_L | 46 | -0.788274214 | 2.393632929 | 0.432843649 | 0 |
| SLF_2_L_FD | SLF_2_L | 47 | -1.43488246 | 2.376789376 | 0.155177324 | 0 |
| SLF_2_L_FD | SLF_2_L | 48 | -0.854655512 | 2.367500707 | 0.395287355 | 0 |
| SLF_2_L_FD | SLF_2_L | 49 | -0.248357428 | 2.432395407 | 0.804543064 | 0 |
| SLF_2_L_FD | SLF_2_L | 50 | -0.202191762 | 2.39644312 | 0.840335533 | 0 |
| SLF_2_R_FD | SLF_2_R | 1 | 0.904323306 | 2.354476382 | 0.36864813 | 0 |
| SLF_2_R_FD | SLF_2_R | 2 | -0.138766774 | 2.423173189 | 0.890057119 | 0 |
| SLF_2_R_FD | SLF_2_R | 3 | -0.921122182 | 2.430612206 | 0.360124224 | 0 |
| SLF_2_R_FD | SLF_2_R | 4 | -1.512791165 | 2.361802935 | 0.134500663 | 0 |
| SLF_2_R_FD | SLF_2_R | 5 | -2.391052726 | 2.35235579 | 0.01928324 | 1 |
| SLF_2_R_FD | SLF_2_R | 6 | -2.637940993 | 2.420284161 | 0.010124902 | 1 |
| SLF_2_R_FD | SLF_2_R | 7 | -1.623540612 | 2.415067799 | 0.10838452 | 0 |
| SLF_2_R_FD | SLF_2_R | 8 | -0.099979067 | 2.42434997 | 0.920610985 | 0 |
| SLF_2_R_FD | SLF_2_R | 9 | 0.201443033 | 2.369683746 | 0.840903357 | 0 |
| SLF_2_R_FD | SLF_2_R | 10 | 0.450419205 | 2.327613269 | 0.653935868 | 0 |
| SLF_2_R_FD | SLF_2_R | 11 | 0.306685096 | 2.413468085 | 0.760042755 | 0 |
| SLF_2_R_FD | SLF_2_R | 12 | 0.234017011 | 2.419903514 | 0.815591352 | 0 |
| SLF_2_R_FD | SLF_2_R | 13 | 0.607871647 | 2.464258281 | 0.544986139 | 0 |
| SLF_2_R_FD | SLF_2_R | 14 | 0.602493533 | 2.343746386 | 0.548551117 | 0 |
| SLF_2_R_FD | SLF_2_R | 15 | 0.805651933 | 2.426544877 | 0.422848897 | 0 |
| SLF_2_R_FD | SLF_2_R | 16 | 0.906499473 | 2.37212852 | 0.367495262 | 0 |

| | | | | | | |
|---|---|---|---|---|---|---|
| SLF_2_R_FD | SLF_2_R | 17 | 1.346646147 | 2.338295639 | 0.182008242 | 0 |
| SLF_2_R_FD | SLF_2_R | 18 | 1.191320999 | 2.344977771 | 0.237233457 | 0 |
| SLF_2_R_FD | SLF_2_R | 19 | 0.897739401 | 2.348848588 | 0.372290411 | 0 |
| SLF_2_R_FD | SLF_2_R | 20 | 0.575335127 | 2.395966053 | 0.566753847 | 0 |
| SLF_2_R_FD | SLF_2_R | 21 | 0.274501201 | 2.381998916 | 0.784421659 | 0 |
| SLF_2_R_FD | SLF_2_R | 22 | 0.460083011 | 2.420719668 | 0.646765572 | 0 |
| SLF_2_R_FD | SLF_2_R | 23 | 0.800155523 | 2.347895631 | 0.426214987 | 0 |
| SLF_2_R_FD | SLF_2_R | 24 | 0.701786491 | 2.345783694 | 0.485272382 | 0 |
| SLF_2_R_FD | SLF_2_R | 25 | 0.43904127 | 2.420377822 | 0.662146908 | 0 |
| SLF_2_R_FD | SLF_2_R | 26 | 0.909982255 | 2.386031709 | 0.366373888 | 0 |
| SLF_2_R_FD | SLF_2_R | 27 | 0.814958555 | 2.394859552 | 0.417862915 | 0 |
| SLF_2_R_FD | SLF_2_R | 28 | 0.101329979 | 2.359625304 | 0.919564163 | 0 |
| SLF_2_R_FD | SLF_2_R | 29 | -0.67565448 | 2.386716737 | 0.501421959 | 0 |
| SLF_2_R_FD | SLF_2_R | 30 | -0.409149309 | 2.31390363 | 0.683558425 | 0 |
| SLF_2_R_FD | SLF_2_R | 31 | -0.059085 | 2.405867856 | 0.953033388 | 0 |
| SLF_2_R_FD | SLF_2_R | 32 | 0.154688309 | 2.419216965 | 0.877454497 | 0 |
| SLF_2_R_FD | SLF_2_R | 33 | 0.557934232 | 2.359335872 | 0.578471836 | 0 |
| SLF_2_R_FD | SLF_2_R | 34 | 0.935049786 | 2.409543421 | 0.35301294 | 0 |
| SLF_2_R_FD | SLF_2_R | 35 | 1.182142757 | 2.428747938 | 0.241216158 | 0 |
| SLF_2_R_FD | SLF_2_R | 36 | 2.040111285 | 2.33254177 | 0.044858271 | 0 |
| SLF_2_R_FD | SLF_2_R | 37 | 1.887086535 | 2.390100624 | 0.063006916 | 0 |
| SLF_2_R_FD | SLF_2_R | 38 | 1.148124294 | 2.430885133 | 0.25480923 | 0 |
| SLF_2_R_FD | SLF_2_R | 39 | 0.441367636 | 2.37736179 | 0.660232804 | 0 |
| SLF_2_R_FD | SLF_2_R | 40 | 0.440732435 | 2.378286511 | 0.660581718 | 0 |
| SLF_2_R_FD | SLF_2_R | 41 | 0.453009641 | 2.370061958 | 0.651766018 | 0 |
| SLF_2_R_FD | SLF_2_R | 42 | 0.603054052 | 2.419812911 | 0.548238995 | 0 |
| SLF_2_R_FD | SLF_2_R | 43 | 1.661853622 | 2.406388374 | 0.101152145 | 0 |
| SLF_2_R_FD | SLF_2_R | 44 | 1.732778809 | 2.412426916 | 0.087721031 | 0 |
| SLF_2_R_FD | SLF_2_R | 45 | 0.509391471 | 2.368062125 | 0.612001826 | 0 |
| SLF_2_R_FD | SLF_2_R | 46 | 0.628849652 | 2.448027719 | 0.531269028 | 0 |
| SLF_2_R_FD | SLF_2_R | 47 | -1.222928171 | 2.365794711 | 0.225136482 | 0 |
| SLF_2_R_FD | SLF_2_R | 48 | -2.029059434 | 2.430927886 | 0.045736967 | 0 |
| SLF_2_R_FD | SLF_2_R | 49 | -1.587724158 | 2.39640961 | 0.116381275 | 0 |
| SLF_2_R_FD | SLF_2_R | 50 | -0.871465924 | 2.401918697 | 0.386306573 | 0 |
| SLF_3_L_FD | SLF_3_L | 1 | 1.315781997 | 2.422108314 | 0.19229948 | 0 |
| SLF_3_L_FD | SLF_3_L | 2 | 0.827625537 | 2.401356971 | 0.410592333 | 0 |
| SLF_3_L_FD | SLF_3_L | 3 | 0.601349058 | 2.414880919 | 0.549367186 | 0 |
| SLF_3_L_FD | SLF_3_L | 4 | 1.129877191 | 2.408613581 | 0.261972186 | 0 |
| SLF_3_L_FD | SLF_3_L | 5 | 1.35941225 | 2.358551921 | 0.178178871 | 0 |
| SLF_3_L_FD | SLF_3_L | 6 | 0.476784472 | 2.350682404 | 0.634909731 | 0 |
| SLF_3_L_FD | SLF_3_L | 7 | 0.641383018 | 2.399984905 | 0.523443526 | 0 |
| SLF_3_L_FD | SLF_3_L | 8 | 1.343869221 | 2.402656253 | 0.183508512 | 0 |
| SLF_3_L_FD | SLF_3_L | 9 | 0.586328886 | 2.434112229 | 0.559643218 | 0 |

| | | | | | | |
|---|---|---|---|---|---|---|
| SLF_3_L_FD | SLF_3_L | 10 | 1.191447368 | 2.34142323 | 0.237655566 | 0 |
| SLF_3_L_FD | SLF_3_L | 11 | 1.508694075 | 2.319053325 | 0.136228112 | 0 |
| SLF_3_L_FD | SLF_3_L | 12 | 1.495794035 | 2.357456339 | 0.138974356 | 0 |
| SLF_3_L_FD | SLF_3_L | 13 | 1.763021236 | 2.372247691 | 0.08198448 | 0 |
| SLF_3_L_FD | SLF_3_L | 14 | 2.388875032 | 2.393907888 | 0.019369151 | 0 |
| SLF_3_L_FD | SLF_3_L | 15 | 3.554545244 | 2.379544269 | 0.000639597 | 1 |
| SLF_3_L_FD | SLF_3_L | 16 | 3.670673839 | 2.341108473 | 0.000433344 | 1 |
| SLF_3_L_FD | SLF_3_L | 17 | 3.168907862 | 2.408435597 | 0.002189172 | 1 |
| SLF_3_L_FD | SLF_3_L | 18 | 2.78328459 | 2.389671863 | 0.006779696 | 1 |
| SLF_3_L_FD | SLF_3_L | 19 | 2.594666417 | 2.404342103 | 0.011323815 | 1 |
| SLF_3_L_FD | SLF_3_L | 20 | 2.433956246 | 2.446560335 | 0.017140509 | 0 |
| SLF_3_L_FD | SLF_3_L | 21 | 2.325289213 | 2.407036324 | 0.022573584 | 0 |
| SLF_3_L_FD | SLF_3_L | 22 | 1.979653399 | 2.372027756 | 0.051199195 | 0 |
| SLF_3_L_FD | SLF_3_L | 23 | 1.345502851 | 2.392799473 | 0.182578475 | 0 |
| SLF_3_L_FD | SLF_3_L | 24 | 0.496048386 | 2.382050822 | 0.6214691 | 0 |
| SLF_3_L_FD | SLF_3_L | 25 | 0.360747942 | 2.328702053 | 0.719387824 | 0 |
| SLF_3_L_FD | SLF_3_L | 26 | 0.867574891 | 2.354760685 | 0.388726891 | 0 |
| SLF_3_L_FD | SLF_3_L | 27 | 1.064449951 | 2.315812981 | 0.291012604 | 0 |
| SLF_3_L_FD | SLF_3_L | 28 | 1.354579131 | 2.406249905 | 0.179963742 | 0 |
| SLF_3_L_FD | SLF_3_L | 29 | 1.686599865 | 2.373045303 | 0.095768465 | 0 |
| SLF_3_L_FD | SLF_3_L | 30 | 1.362163402 | 2.395682844 | 0.177100419 | 0 |
| SLF_3_L_FD | SLF_3_L | 31 | 0.779457457 | 2.353952743 | 0.438017802 | 0 |
| SLF_3_L_FD | SLF_3_L | 32 | 0.455023354 | 2.326699486 | 0.650309931 | 0 |
| SLF_3_L_FD | SLF_3_L | 33 | 0.863202718 | 2.375929952 | 0.39058692 | 0 |
| SLF_3_L_FD | SLF_3_L | 34 | 1.050062154 | 2.366687562 | 0.296817136 | 0 |
| SLF_3_L_FD | SLF_3_L | 35 | 0.707509907 | 2.326821262 | 0.481417974 | 0 |
| SLF_3_L_FD | SLF_3_L | 36 | 0.569023509 | 2.422906737 | 0.571134798 | 0 |
| SLF_3_L_FD | SLF_3_L | 37 | 0.876355699 | 2.321451504 | 0.383801977 | 0 |
| SLF_3_L_FD | SLF_3_L | 38 | 0.954697586 | 2.379206336 | 0.342695598 | 0 |
| SLF_3_L_FD | SLF_3_L | 39 | 0.740162295 | 2.415517571 | 0.461342686 | 0 |
| SLF_3_L_FD | SLF_3_L | 40 | 1.086703774 | 2.4172966 | 0.280652399 | 0 |
| SLF_3_L_FD | SLF_3_L | 41 | 0.327888713 | 2.373841683 | 0.743843262 | 0 |
| SLF_3_L_FD | SLF_3_L | 42 | -0.279732691 | 2.399611319 | 0.780398373 | 0 |
| SLF_3_L_FD | SLF_3_L | 43 | -0.997739092 | 2.388592933 | 0.321582244 | 0 |
| SLF_3_L_FD | SLF_3_L | 44 | -0.190006441 | 2.437666988 | 0.849785383 | 0 |
| SLF_3_L_FD | SLF_3_L | 45 | 0.253799145 | 2.382185194 | 0.800293978 | 0 |
| SLF_3_L_FD | SLF_3_L | 46 | -0.573529907 | 2.411362229 | 0.567878575 | 0 |
| SLF_3_L_FD | SLF_3_L | 47 | -1.467054134 | 2.397483308 | 0.146253429 | 0 |
| SLF_3_L_FD | SLF_3_L | 48 | -0.956752689 | 2.446829279 | 0.341550427 | 0 |
| SLF_3_L_FD | SLF_3_L | 49 | -0.780487389 | 2.375033551 | 0.437403923 | 0 |
| SLF_3_L_FD | SLF_3_L | 50 | -0.752689238 | 2.418318853 | 0.454056801 | 0 |
| SLF_3_R_FD | SLF_3_R | 1 | -0.283686906 | 2.340423902 | 0.777481072 | 0 |
| SLF_3_R_FD | SLF_3_R | 2 | 0.043386295 | 2.347270351 | 0.965536301 | 0 |

| | | | | | | |
|---|---|---|---|---|---|---|
| SLF_3_R_FD | SLF_3_R | 3 | 0.704707844 | 2.414505534 | 0.483385602 | 0 |
| SLF_3_R_FD | SLF_3_R | 4 | 0.672207942 | 2.430464217 | 0.503442751 | 0 |
| SLF_3_R_FD | SLF_3_R | 5 | -0.24603388 | 2.352142513 | 0.806316782 | 0 |
| SLF_3_R_FD | SLF_3_R | 6 | -0.031429855 | 2.417325895 | 0.975011048 | 0 |
| SLF_3_R_FD | SLF_3_R | 7 | 0.640368815 | 2.398429912 | 0.52382558 | 0 |
| SLF_3_R_FD | SLF_3_R | 8 | 0.353274954 | 2.329419468 | 0.724812133 | 0 |
| SLF_3_R_FD | SLF_3_R | 9 | -0.629524917 | 2.40875913 | 0.530844508 | 0 |
| SLF_3_R_FD | SLF_3_R | 10 | -0.205134166 | 2.389941691 | 0.838016721 | 0 |
| SLF_3_R_FD | SLF_3_R | 11 | 0.398263113 | 2.431445153 | 0.691555238 | 0 |
| SLF_3_R_FD | SLF_3_R | 12 | 0.717843999 | 2.363918631 | 0.474995052 | 0 |
| SLF_3_R_FD | SLF_3_R | 13 | 0.785256197 | 2.409796608 | 0.434672762 | 0 |
| SLF_3_R_FD | SLF_3_R | 14 | 0.650413828 | 2.399088527 | 0.517414437 | 0 |
| SLF_3_R_FD | SLF_3_R | 15 | 0.564806809 | 2.385956323 | 0.573891654 | 0 |
| SLF_3_R_FD | SLF_3_R | 16 | 0.767041567 | 2.442512553 | 0.44537529 | 0 |
| SLF_3_R_FD | SLF_3_R | 17 | 1.044168666 | 2.409056938 | 0.299656842 | 0 |
| SLF_3_R_FD | SLF_3_R | 18 | 1.16917311 | 2.372964618 | 0.246223089 | 0 |
| SLF_3_R_FD | SLF_3_R | 19 | 1.064694549 | 2.342907937 | 0.291193655 | 0 |
| SLF_3_R_FD | SLF_3_R | 20 | 0.844217293 | 2.347322632 | 0.401991047 | 0 |
| SLF_3_R_FD | SLF_3_R | 21 | 0.607429579 | 2.400189492 | 0.545806132 | 0 |
| SLF_3_R_FD | SLF_3_R | 22 | 0.384066407 | 2.399773072 | 0.702199764 | 0 |
| SLF_3_R_FD | SLF_3_R | 23 | 0.08067757 | 2.388492127 | 0.935918301 | 0 |
| SLF_3_R_FD | SLF_3_R | 24 | 0.115210211 | 2.341738905 | 0.908575516 | 0 |
| SLF_3_R_FD | SLF_3_R | 25 | 0.497569884 | 2.379318267 | 0.620221271 | 0 |
| SLF_3_R_FD | SLF_3_R | 26 | 1.011610547 | 2.326182895 | 0.315078171 | 0 |
| SLF_3_R_FD | SLF_3_R | 27 | 1.15161308 | 2.370004207 | 0.253525269 | 0 |
| SLF_3_R_FD | SLF_3_R | 28 | 0.924234667 | 2.422965383 | 0.358853172 | 0 |
| SLF_3_R_FD | SLF_3_R | 29 | 0.80714762 | 2.406731581 | 0.422529494 | 0 |
| SLF_3_R_FD | SLF_3_R | 30 | 0.938200755 | 2.344074772 | 0.351753862 | 0 |
| SLF_3_R_FD | SLF_3_R | 31 | 0.620764515 | 2.405140058 | 0.537034465 | 0 |
| SLF_3_R_FD | SLF_3_R | 32 | -0.1501187 | 2.392531802 | 0.881089713 | 0 |
| SLF_3_R_FD | SLF_3_R | 33 | -0.441178487 | 2.368780084 | 0.660344761 | 0 |
| SLF_3_R_FD | SLF_3_R | 34 | -0.189251276 | 2.394484897 | 0.850382895 | 0 |
| SLF_3_R_FD | SLF_3_R | 35 | -0.210960178 | 2.367998477 | 0.833449668 | 0 |
| SLF_3_R_FD | SLF_3_R | 36 | 0.336204999 | 2.383854627 | 0.737585816 | 0 |
| SLF_3_R_FD | SLF_3_R | 37 | 0.523025941 | 2.398302224 | 0.602448226 | 0 |
| SLF_3_R_FD | SLF_3_R | 38 | 0.974871532 | 2.391972411 | 0.333060719 | 0 |
| SLF_3_R_FD | SLF_3_R | 39 | 0.729221252 | 2.405204889 | 0.468157493 | 0 |
| SLF_3_R_FD | SLF_3_R | 40 | 1.02489173 | 2.321604202 | 0.308467185 | 0 |
| SLF_3_R_FD | SLF_3_R | 41 | 1.643754516 | 2.352609447 | 0.104128825 | 0 |
| SLF_3_R_FD | SLF_3_R | 42 | 0.312593891 | 2.450803924 | 0.755423709 | 0 |
| SLF_3_R_FD | SLF_3_R | 43 | 0.682598975 | 2.384648922 | 0.496928633 | 0 |
| SLF_3_R_FD | SLF_3_R | 44 | 0.147339264 | 2.40623412 | 0.883232512 | 0 |
| SLF_3_R_FD | SLF_3_R | 45 | -0.861937813 | 2.372786539 | 0.391276185 | 0 |

| | | | | | | |
|---|---|---|---|---|---|---|
| SLF_3_R_FD | SLF_3_R | 46 | -0.347561848 | 2.439725064 | 0.729092171 | 0 |
| SLF_3_R_FD | SLF_3_R | 47 | 0.493910918 | 2.411508602 | 0.622743766 | 0 |
| SLF_3_R_FD | SLF_3_R | 48 | -0.368231511 | 2.366645925 | 0.713663708 | 0 |
| SLF_3_R_FD | SLF_3_R | 49 | 0.378165686 | 2.392779476 | 0.706404828 | 0 |
| SLF_3_R_FD | SLF_3_R | 50 | 0.249134043 | 2.377129268 | 0.803981774 | 0 |
| UF_L_FD | UF_L | 1 | -1.658116693 | 2.410463717 | 0.101259609 | 0 |
| UF_L_FD | UF_L | 2 | -1.869173627 | 2.382438471 | 0.065404235 | 0 |
| UF_L_FD | UF_L | 3 | -1.682561077 | 2.407302043 | 0.096399875 | 0 |
| UF_L_FD | UF_L | 4 | -1.526875134 | 2.36749672 | 0.130724487 | 0 |
| UF_L_FD | UF_L | 5 | -1.566695346 | 2.381170382 | 0.121179772 | 0 |
| UF_L_FD | UF_L | 6 | -1.778246788 | 2.411993954 | 0.079273794 | 0 |
| UF_L_FD | UF_L | 7 | -1.795831273 | 2.439897521 | 0.076686359 | 0 |
| UF_L_FD | UF_L | 8 | -1.189179209 | 2.443194822 | 0.238127866 | 0 |
| UF_L_FD | UF_L | 9 | -0.760449649 | 2.464492771 | 0.449395758 | 0 |
| UF_L_FD | UF_L | 10 | -0.347689009 | 2.478902549 | 0.729078233 | 0 |
| UF_L_FD | UF_L | 11 | 0.174153737 | 2.4478783 | 0.862232382 | 0 |
| UF_L_FD | UF_L | 12 | 0.319909372 | 2.410840083 | 0.750010998 | 0 |
| UF_L_FD | UF_L | 13 | -0.033229649 | 2.36866385 | 0.973590225 | 0 |
| UF_L_FD | UF_L | 14 | 0.185724679 | 2.403565387 | 0.853236574 | 0 |
| UF_L_FD | UF_L | 15 | 1.084754369 | 2.411725525 | 0.281646812 | 0 |
| UF_L_FD | UF_L | 16 | 1.035751371 | 2.352076679 | 0.303681212 | 0 |
| UF_L_FD | UF_L | 17 | 1.508849609 | 2.407854169 | 0.135678038 | 0 |
| UF_L_FD | UF_L | 18 | 2.156157452 | 2.430441135 | 0.034652741 | 0 |
| UF_L_FD | UF_L | 19 | 1.708737596 | 2.410228719 | 0.092287388 | 0 |
| UF_L_FD | UF_L | 20 | 1.325389758 | 2.397526865 | 0.189549839 | 0 |
| UF_L_FD | UF_L | 21 | 1.433876759 | 2.351636587 | 0.155928106 | 0 |
| UF_L_FD | UF_L | 22 | 1.633904815 | 2.397706149 | 0.106400454 | 0 |
| UF_L_FD | UF_L | 23 | 1.711125669 | 2.39956453 | 0.091190673 | 0 |
| UF_L_FD | UF_L | 24 | 1.370552276 | 2.326973705 | 0.174722533 | 0 |
| UF_L_FD | UF_L | 25 | 0.664314376 | 2.400201127 | 0.508735234 | 0 |
| UF_L_FD | UF_L | 26 | 0.116613979 | 2.305268583 | 0.907510688 | 0 |
| UF_L_FD | UF_L | 27 | -0.211827207 | 2.422253849 | 0.832856781 | 0 |
| UF_L_FD | UF_L | 28 | -0.519725156 | 2.381474423 | 0.604767378 | 0 |
| UF_L_FD | UF_L | 29 | -0.880477461 | 2.404807794 | 0.381232897 | 0 |
| UF_L_FD | UF_L | 30 | -0.955266842 | 2.320924294 | 0.342285523 | 0 |
| UF_L_FD | UF_L | 31 | -0.900644067 | 2.41355551 | 0.370448308 | 0 |
| UF_L_FD | UF_L | 32 | -0.707380693 | 2.398845255 | 0.481360923 | 0 |
| UF_L_FD | UF_L | 33 | -0.277181116 | 2.335193589 | 0.782349454 | 0 |
| UF_L_FD | UF_L | 34 | 0.139293314 | 2.334707352 | 0.889582632 | 0 |
| UF_L_FD | UF_L | 35 | 0.853885993 | 2.393729559 | 0.395754439 | 0 |
| UF_L_FD | UF_L | 36 | 1.349666727 | 2.366905723 | 0.180905984 | 0 |
| UF_L_FD | UF_L | 37 | 1.350185842 | 2.382441278 | 0.180717161 | 0 |
| UF_L_FD | UF_L | 38 | 1.751763859 | 2.390840051 | 0.083600699 | 0 |

| | | | | | | |
|---|---|---|---|---|---|---|
| UF_L_FD | UF_L | 39 | 1.658226571 | 2.358743192 | 0.101147444 | 0 |
| UF_L_FD | UF_L | 40 | 1.143849806 | 2.345721163 | 0.256098089 | 0 |
| UF_L_FD | UF_L | 41 | 0.573407168 | 2.453996887 | 0.567990527 | 0 |
| UF_L_FD | UF_L | 42 | 0.117854597 | 2.404877894 | 0.906481928 | 0 |
| UF_L_FD | UF_L | 43 | -0.090323201 | 2.403206243 | 0.928255773 | 0 |
| UF_L_FD | UF_L | 44 | -0.457383094 | 2.326702023 | 0.648628189 | 0 |
| UF_L_FD | UF_L | 45 | -0.996096162 | 2.36611088 | 0.322239998 | 0 |
| UF_L_FD | UF_L | 46 | -0.859660545 | 2.31289115 | 0.392663182 | 0 |
| UF_L_FD | UF_L | 47 | -0.141902934 | 2.412717832 | 0.887570199 | 0 |
| UF_L_FD | UF_L | 48 | 0.818997685 | 2.350586641 | 0.415746287 | 0 |
| UF_L_FD | UF_L | 49 | 1.04124504 | 2.398820563 | 0.301571062 | 0 |
| UF_L_FD | UF_L | 50 | 1.331946013 | 2.404049407 | 0.186630827 | 0 |
| UF_R_FD | UF_R | 1 | 0.257390025 | 2.405303374 | 0.797631226 | 0 |
| UF_R_FD | UF_R | 2 | 0.754871308 | 2.381550293 | 0.452690745 | 0 |
| UF_R_FD | UF_R | 3 | 0.991406057 | 2.395608728 | 0.32474717 | 0 |
| UF_R_FD | UF_R | 4 | 0.710871503 | 2.352617335 | 0.479472692 | 0 |
| UF_R_FD | UF_R | 5 | 0.486741294 | 2.41284208 | 0.628045669 | 0 |
| UF_R_FD | UF_R | 6 | 0.290855415 | 2.426108302 | 0.772124264 | 0 |
| UF_R_FD | UF_R | 7 | 0.235029238 | 2.339864138 | 0.814962896 | 0 |
| UF_R_FD | UF_R | 8 | 0.330902953 | 2.354864327 | 0.741847526 | 0 |
| UF_R_FD | UF_R | 9 | 0.035971281 | 2.418347849 | 0.971420345 | 0 |
| UF_R_FD | UF_R | 10 | 0.071448917 | 2.374254199 | 0.943256174 | 0 |
| UF_R_FD | UF_R | 11 | 0.292930115 | 2.438665022 | 0.770438261 | 0 |
| UF_R_FD | UF_R | 12 | 0.366041084 | 2.464097384 | 0.715471863 | 0 |
| UF_R_FD | UF_R | 13 | 0.722280773 | 2.391384719 | 0.472662232 | 0 |
| UF_R_FD | UF_R | 14 | 0.979660206 | 2.398813143 | 0.330742473 | 0 |
| UF_R_FD | UF_R | 15 | 1.145683762 | 2.38667654 | 0.255983223 | 0 |
| UF_R_FD | UF_R | 16 | 1.625417489 | 2.42239952 | 0.108811933 | 0 |
| UF_R_FD | UF_R | 17 | 2.034807704 | 2.403024961 | 0.045765203 | 0 |
| UF_R_FD | UF_R | 18 | 1.969180839 | 2.377251285 | 0.052880419 | 0 |
| UF_R_FD | UF_R | 19 | 1.707522935 | 2.383722935 | 0.091671233 | 0 |
| UF_R_FD | UF_R | 20 | 1.937605714 | 2.395055805 | 0.056301764 | 0 |
| UF_R_FD | UF_R | 21 | 2.370273504 | 2.356109078 | 0.020354633 | 1 |
| UF_R_FD | UF_R | 22 | 2.360138328 | 2.347590881 | 0.021036398 | 1 |
| UF_R_FD | UF_R | 23 | 2.123353985 | 2.377286954 | 0.037201098 | 0 |
| UF_R_FD | UF_R | 24 | 1.940909525 | 2.393435635 | 0.056141832 | 0 |
| UF_R_FD | UF_R | 25 | 1.785585083 | 2.310468085 | 0.078312445 | 0 |
| UF_R_FD | UF_R | 26 | 1.509253262 | 2.380305487 | 0.135419266 | 0 |
| UF_R_FD | UF_R | 27 | 1.170216627 | 2.391435836 | 0.245511668 | 0 |
| UF_R_FD | UF_R | 28 | 0.765352423 | 2.363845029 | 0.446357906 | 0 |
| UF_R_FD | UF_R | 29 | 0.461038608 | 2.369463738 | 0.646028158 | 0 |
| UF_R_FD | UF_R | 30 | 0.431190041 | 2.404187005 | 0.667480177 | 0 |
| UF_R_FD | UF_R | 31 | 0.386289338 | 2.404585668 | 0.700304726 | 0 |

| | | | | | | |
|---|---|---|---|---|---|---|
| UF_R_FD | UF_R | 32 | 0.474304399 | 2.359063183 | 0.636576814 | 0 |
| UF_R_FD | UF_R | 33 | 0.79994777 | 2.401292305 | 0.426140179 | 0 |
| UF_R_FD | UF_R | 34 | 1.122784081 | 2.387402967 | 0.264880492 | 0 |
| UF_R_FD | UF_R | 35 | 1.749452219 | 2.436178901 | 0.084069496 | 0 |
| UF_R_FD | UF_R | 36 | 2.262614587 | 2.366761865 | 0.02639949 | 0 |
| UF_R_FD | UF_R | 37 | 2.290832581 | 2.379627003 | 0.024677551 | 0 |
| UF_R_FD | UF_R | 38 | 2.171945088 | 2.354004861 | 0.03298837 | 0 |
| UF_R_FD | UF_R | 39 | 2.083286726 | 2.399578018 | 0.040557561 | 0 |
| UF_R_FD | UF_R | 40 | 1.805468463 | 2.402398419 | 0.074856159 | 0 |
| UF_R_FD | UF_R | 41 | 1.314427897 | 2.381803674 | 0.192481768 | 0 |
| UF_R_FD | UF_R | 42 | 0.576622 | 2.344108719 | 0.565800443 | 0 |
| UF_R_FD | UF_R | 43 | -0.289967651 | 2.383617679 | 0.772602634 | 0 |
| UF_R_FD | UF_R | 44 | -0.818609924 | 2.368226645 | 0.415634137 | 0 |
| UF_R_FD | UF_R | 45 | -1.002376344 | 2.42418714 | 0.319511416 | 0 |
| UF_R_FD | UF_R | 46 | -0.910196448 | 2.34646588 | 0.365784153 | 0 |
| UF_R_FD | UF_R | 47 | -0.410946333 | 2.349718792 | 0.682350562 | 0 |
| UF_R_FD | UF_R | 48 | 0.57340136 | 2.359083317 | 0.568190662 | 0 |
| UF_R_FD | UF_R | 49 | 0.780746695 | 2.422443687 | 0.437577638 | 0 |
| UF_R_FD | UF_R | 50 | 0.895968221 | 2.413037441 | 0.372959782 | 0 |
| fornix_L_FD | fornix_L | 1 | 1.58384545 | 2.344629478 | 0.117555094 | 0 |
| fornix_L_FD | fornix_L | 2 | 2.513682613 | 2.403116258 | 0.013941967 | 1 |
| fornix_L_FD | fornix_L | 3 | 1.606978037 | 2.414329568 | 0.112056281 | 0 |
| fornix_L_FD | fornix_L | 4 | 1.123951872 | 2.332810919 | 0.264713833 | 0 |
| fornix_L_FD | fornix_L | 5 | 1.610616204 | 2.358338144 | 0.111880225 | 0 |
| fornix_L_FD | fornix_L | 6 | 1.12085561 | 2.364461643 | 0.266505092 | 0 |
| fornix_L_FD | fornix_L | 7 | 0.599691284 | 2.357468485 | 0.550823101 | 0 |
| fornix_L_FD | fornix_L | 8 | 0.511635698 | 2.407787964 | 0.610698503 | 0 |
| fornix_L_FD | fornix_L | 9 | 0.184674553 | 2.344876536 | 0.85411045 | 0 |
| fornix_L_FD | fornix_L | 10 | -0.375312688 | 2.38856557 | 0.708768175 | 0 |
| fornix_L_FD | fornix_L | 11 | -0.058904362 | 2.362603309 | 0.953235486 | 0 |
| fornix_L_FD | fornix_L | 12 | 0.246328709 | 2.313143272 | 0.806375771 | 0 |
| fornix_L_FD | fornix_L | 13 | 0.509283756 | 2.341083647 | 0.612510759 | 0 |
| fornix_L_FD | fornix_L | 14 | 0.068469937 | 2.34599315 | 0.945604449 | 0 |
| fornix_L_FD | fornix_L | 15 | -0.614441353 | 2.347275428 | 0.540832056 | 0 |
| fornix_L_FD | fornix_L | 16 | -0.584260166 | 2.390824988 | 0.56075551 | 0 |
| fornix_L_FD | fornix_L | 17 | -0.118583675 | 2.325349676 | 0.905898791 | 0 |
| fornix_L_FD | fornix_L | 18 | 0.406484016 | 2.384781884 | 0.68547413 | 0 |
| fornix_L_FD | fornix_L | 19 | 0.240870996 | 2.349193651 | 0.810271468 | 0 |
| fornix_L_FD | fornix_L | 20 | -0.460955754 | 2.390866244 | 0.646071722 | 0 |
| fornix_L_FD | fornix_L | 21 | -0.183654048 | 2.393152248 | 0.854743857 | 0 |
| fornix_L_FD | fornix_L | 22 | 0.129189211 | 2.360366111 | 0.89752931 | 0 |
| fornix_L_FD | fornix_L | 23 | -0.11895523 | 2.374365549 | 0.905617384 | 0 |
| fornix_L_FD | fornix_L | 24 | -0.533455388 | 2.380546879 | 0.595260659 | 0 |

| | | | | | | |
|---|---|---|---|---|---|---|
| fornix_L_FD | fornix_L | 25 | -0.782396921 | 2.448623385 | 0.436288805 | 0 |
| fornix_L_FD | fornix_L | 26 | -1.39521355 | 2.448299883 | 0.166815436 | 0 |
| fornix_L_FD | fornix_L | 27 | -1.966232027 | 2.36627556 | 0.052897386 | 0 |
| fornix_L_FD | fornix_L | 28 | -1.87553786 | 2.366864201 | 0.064640682 | 0 |
| fornix_L_FD | fornix_L | 29 | -1.400072605 | 2.344597729 | 0.16620902 | 0 |
| fornix_L_FD | fornix_L | 30 | -0.833300054 | 2.366231883 | 0.407646257 | 0 |
| fornix_L_FD | fornix_L | 31 | -0.845702197 | 2.364109527 | 0.40053286 | 0 |
| fornix_L_FD | fornix_L | 32 | -1.024097576 | 2.423767237 | 0.309344737 | 0 |
| fornix_L_FD | fornix_L | 33 | -1.108096562 | 2.40433906 | 0.271811243 | 0 |
| fornix_L_FD | fornix_L | 34 | -1.35592611 | 2.432862987 | 0.179627759 | 0 |
| fornix_L_FD | fornix_L | 35 | -1.355331383 | 2.380968818 | 0.179911198 | 0 |
| fornix_L_FD | fornix_L | 36 | -1.343942047 | 2.432670995 | 0.183605502 | 0 |
| fornix_L_FD | fornix_L | 37 | -1.145281371 | 2.415554532 | 0.25620442 | 0 |
| fornix_L_FD | fornix_L | 38 | -0.869272033 | 2.432845917 | 0.38785936 | 0 |
| fornix_L_FD | fornix_L | 39 | -0.962020945 | 2.34503377 | 0.339566527 | 0 |
| fornix_L_FD | fornix_L | 40 | -0.682153504 | 2.408690637 | 0.497357196 | 0 |
| fornix_L_FD | fornix_L | 41 | -0.201617587 | 2.373223347 | 0.840734742 | 0 |
| fornix_L_FD | fornix_L | 42 | -0.471638528 | 2.423109076 | 0.638471378 | 0 |
| fornix_L_FD | fornix_L | 43 | -0.426409423 | 2.332190911 | 0.670968877 | 0 |
| fornix_L_FD | fornix_L | 44 | 0.163302632 | 2.332613101 | 0.87069134 | 0 |
| fornix_L_FD | fornix_L | 45 | 0.825785227 | 2.403979079 | 0.411354261 | 0 |
| fornix_L_FD | fornix_L | 46 | 1.043102977 | 2.399033719 | 0.300036063 | 0 |
| fornix_L_FD | fornix_L | 47 | 1.018604217 | 2.42553302 | 0.311616546 | 0 |
| fornix_L_FD | fornix_L | 48 | 1.240495055 | 2.450628432 | 0.218649322 | 0 |
| fornix_L_FD | fornix_L | 49 | 1.065038727 | 2.387965194 | 0.290260144 | 0 |
| fornix_L_FD | fornix_L | 50 | 1.639766388 | 2.371450202 | 0.105243483 | 0 |
| fornix_R_FD | fornix_R | 1 | 1.530800102 | 2.475680638 | 0.130125189 | 0 |
| fornix_R_FD | fornix_R | 2 | 1.136697463 | 2.382575374 | 0.259029775 | 0 |
| fornix_R_FD | fornix_R | 3 | 0.918932365 | 2.387573156 | 0.360868967 | 0 |
| fornix_R_FD | fornix_R | 4 | 0.62910743 | 2.347271981 | 0.531078757 | 0 |
| fornix_R_FD | fornix_R | 5 | 0.739603319 | 2.392783995 | 0.461777658 | 0 |
| fornix_R_FD | fornix_R | 6 | 0.680849077 | 2.362456796 | 0.498060441 | 0 |
| fornix_R_FD | fornix_R | 7 | 0.54410591 | 2.405568894 | 0.588001793 | 0 |
| fornix_R_FD | fornix_R | 8 | 0.827004998 | 2.360479951 | 0.411005699 | 0 |
| fornix_R_FD | fornix_R | 9 | 0.582990535 | 2.354015342 | 0.561662082 | 0 |
| fornix_R_FD | fornix_R | 10 | 0.582496308 | 2.401711627 | 0.562079968 | 0 |
| fornix_R_FD | fornix_R | 11 | 0.248263145 | 2.415960537 | 0.804638568 | 0 |
| fornix_R_FD | fornix_R | 12 | -0.094007639 | 2.378740574 | 0.925350632 | 0 |
| fornix_R_FD | fornix_R | 13 | -0.372889212 | 2.374475406 | 0.710237536 | 0 |
| fornix_R_FD | fornix_R | 14 | -0.323943238 | 2.443715612 | 0.746860434 | 0 |
| fornix_R_FD | fornix_R | 15 | -0.244533449 | 2.413560439 | 0.807496979 | 0 |
| fornix_R_FD | fornix_R | 16 | -0.024387765 | 2.410392957 | 0.980606321 | 0 |
| fornix_R_FD | fornix_R | 17 | -0.600041801 | 2.409456168 | 0.550207268 | 0 |

| | | | | | | |
|---|---|---|---|---|---|---|
| fornix_R_FD | fornix_R | 18 | -1.05955858 | 2.384148052 | 0.29271948 | 0 |
| fornix_R_FD | fornix_R | 19 | -1.035824563 | 2.382521425 | 0.303551635 | 0 |
| fornix_R_FD | fornix_R | 20 | -0.207443247 | 2.342646384 | 0.836202652 | 0 |
| fornix_R_FD | fornix_R | 21 | -0.008618594 | 2.382001524 | 0.993146079 | 0 |
| fornix_R_FD | fornix_R | 22 | -0.385473379 | 2.410216817 | 0.701013609 | 0 |
| fornix_R_FD | fornix_R | 23 | -1.017656736 | 2.387775585 | 0.31277762 | 0 |
| fornix_R_FD | fornix_R | 24 | -1.056053558 | 2.380173679 | 0.295738765 | 0 |
| fornix_R_FD | fornix_R | 25 | -0.575027172 | 2.306744816 | 0.56749055 | 0 |
| fornix_R_FD | fornix_R | 26 | -0.722114884 | 2.365493817 | 0.472538549 | 0 |
| fornix_R_FD | fornix_R | 27 | -0.983427295 | 2.451740952 | 0.328349543 | 0 |
| fornix_R_FD | fornix_R | 28 | -1.362221509 | 2.398626779 | 0.176969882 | 0 |
| fornix_R_FD | fornix_R | 29 | -1.419647286 | 2.337571244 | 0.159823166 | 0 |
| fornix_R_FD | fornix_R | 30 | -1.573217355 | 2.399137833 | 0.119766329 | 0 |
| fornix_R_FD | fornix_R | 31 | -1.99333128 | 2.4159247 | 0.049822049 | 0 |
| fornix_R_FD | fornix_R | 32 | -1.5707452 | 2.36988882 | 0.120903442 | 0 |
| fornix_R_FD | fornix_R | 33 | -0.793952413 | 2.42169248 | 0.430218271 | 0 |
| fornix_R_FD | fornix_R | 34 | 0.294193581 | 2.413870882 | 0.769600614 | 0 |
| fornix_R_FD | fornix_R | 35 | 0.211607115 | 2.444822182 | 0.833039041 | 0 |
| fornix_R_FD | fornix_R | 36 | -0.059001152 | 2.381185761 | 0.953112029 | 0 |
| fornix_R_FD | fornix_R | 37 | -0.018547285 | 2.425940429 | 0.985253579 | 0 |
| fornix_R_FD | fornix_R | 38 | 0.47154779 | 2.398010001 | 0.638741059 | 0 |
| fornix_R_FD | fornix_R | 39 | 0.889980962 | 2.337888413 | 0.376637587 | 0 |
| fornix_R_FD | fornix_R | 40 | 0.746356401 | 2.359982014 | 0.458085145 | 0 |
| fornix_R_FD | fornix_R | 41 | 0.576321172 | 2.376411437 | 0.566400497 | 0 |
| fornix_R_FD | fornix_R | 42 | 0.499649155 | 2.387072786 | 0.618889757 | 0 |
| fornix_R_FD | fornix_R | 43 | 0.503373757 | 2.399633865 | 0.616115839 | 0 |
| fornix_R_FD | fornix_R | 44 | 0.527872609 | 2.39753491 | 0.599065328 | 0 |
| fornix_R_FD | fornix_R | 45 | -0.230190192 | 2.410244575 | 0.818532568 | 0 |
| fornix_R_FD | fornix_R | 46 | -0.085782537 | 2.416419556 | 0.931850999 | 0 |
| fornix_R_FD | fornix_R | 47 | 0.13726388 | 2.385474461 | 0.891162854 | 0 |
| fornix_R_FD | fornix_R | 48 | 0.048767373 | 2.396092008 | 0.961226037 | 0 |
| fornix_R_FD | fornix_R | 49 | -0.206576467 | 2.42367474 | 0.836881678 | 0 |
| fornix_R_FD | fornix_R | 50 | 0.818061973 | 2.382997366 | 0.416307149 | 0 |

Abbreviations: FD, fiber-specific apparent fiber density; AF, arcuate fasciculus; CC, corpus callosum (CC_1, rostrum; CC_2a, anterior genu; CC_2b, posterior genu; CC_3, rostral body; CC_4, anterior mid-body; CC_5, posterior mid-body; CC_6, isthmus; CC_7, splenium); CG, cingulum; IFOF, inferior frontal occipital fasciculus; ILF, inferior longitudinal fasciculus; SLF, superior longitudinal fasciculus; UF, uncinate fasciculus; CR, corona radiata; CST, corticospinal tract; OR, optic radiation; SCP, superior cerebellar peduncle; MCP, middle cerebellar peduncle; L, left; R, right; sig, significant.
